# Multi-ancestry genome-wide study in >2.5 million individuals reveals heterogeneity in mechanistic pathways of type 2 diabetes and complications

**DOI:** 10.1101/2023.03.31.23287839

**Authors:** Ken Suzuki, Konstantinos Hatzikotoulas, Lorraine Southam, Henry J. Taylor, Xianyong Yin, Kim M. Lorenz, Ravi Mandla, Alicia Huerta-Chagoya, Nigel W. Rayner, Ozvan Bocher, S. V. Arruda Ana Luiza de, Kyuto Sonehara, Shinichi Namba, Simon S. K. Lee, Michael H. Preuss, Lauren E. Petty, Philip Schroeder, Brett Vanderwerff, Mart Kals, Fiona Bragg, Kuang Lin, Xiuqing Guo, Weihua Zhang, Jie Yao, Young Jin Kim, Mariaelisa Graff, Fumihiko Takeuchi, Jana Nano, Amel Lamri, Masahiro Nakatochi, Sanghoon Moon, Robert A. Scott, James P. Cook, Jung-Jin Lee, Ian Pan, Daniel Taliun, Esteban J. Parra, Jin-Fang Chai, Lawrence F. Bielak, Yasuharu Tabara, Yang Hai, Gudmar Thorleifsson, Niels Grarup, Tamar Sofer, Matthias Wuttke, Chloé Sarnowski, Christian Gieger, Darryl Nousome, Stella Trompet, Soo-Heon Kwak, Jirong Long, Meng Sun, Lin Tong, Wei-Min Chen, Suraj S. Nongmaithem, Raymond Noordam, Victor J. Y. Lim, Claudia H. T. Tam, Yoonjung Yoonie Joo, Chien-Hsiun Chen, Laura M. Raffield, Bram Peter Prins, Aude Nicolas, Lisa R. Yanek, Guanjie Chen, Jennifer A. Brody, Edmond Kabagambe, Ping An, Anny H. Xiang, Hyeok Sun Choi, Brian E. Cade, Jingyi Tan, K. Alaine Broadaway, Alice Williamson, Zoha Kamali, Jinrui Cui, Linda S. Adair, Adebowale Adeyemo, Carlos A. Aguilar-Salinas, Tarunveer S. Ahluwalia, Sonia S. Anand, Alain Bertoni, Jette Bork-Jensen, Ivan Brandslund, Thomas A. Buchanan, Charles F. Burant, Adam S. Butterworth, Mickaël Canouil, Juliana C. N. Chan, Li-Ching Chang, Miao-Li Chee, Ji Chen, Shyh-Huei Chen, Yuan-Tsong Chen, Zhengming Chen, Lee-Ming Chuang, Mary Cushman, John Danesh, Swapan K. Das, H. Janaka de Silva, George Dedoussis, Latchezar Dimitrov, Ayo P. Doumatey, Shufa Du, Qing Duan, Kai-Uwe Eckardt, Leslie S. Emery, Daniel S. Evans, Michele K. Evans, Krista Fischer, James S. Floyd, Ian Ford, Oscar H. Franco, Timothy M. Frayling, Barry I. Freedman, Pauline Genter, Hertzel C. Gerstein, Vilmantas Giedraitis, Clicerio González-Villalpando, Maria Elena González-Villalpando, Penny Gordon-Larsen, Myron Gross, Lindsay A. Guare, Sophie Hackinger, Sohee Han, Andrew T. Hattersley, Christian Herder, Momoko Horikoshi, Annie-Green Howard, Willa Hsueh, Mengna Huang, Wei Huang, Yi-Jen Hung, Mi Yeong Hwang, Chii-Min Hwu, Sahoko Ichihara, Mohammad Arfan Ikram, Martin Ingelsson, Md. Tariqul Islam, Masato Isono, Hye-Mi Jang, Farzana Jasmine, Guozhi Jiang, Jost B. Jonas, Torben Jørgensen, Fouad R. Kandeel, Anuradhani Kasturiratne, Tomohiro Katsuya, Varinderpal Kaur, Takahisa Kawaguchi, Jacob M. Keaton, Abel N. Kho, Chiea-Chuen Khor, Muhammad G. Kibriya, Duk-Hwan Kim, Florian Kronenberg, Johanna Kuusisto, Kristi Läll, Leslie A. Lange, Kyung Min Lee, Myung-Shik Lee, Nanette R. Lee, Aaron Leong, Liming Li, Yun Li, Ruifang Li-Gao, Symen Lithgart, Cecilia M. Lindgren, Allan Linneberg, Ching-Ti Liu, Jianjun Liu, Adam E. Locke, Tin Louie, Jian’an Luan, Andrea O. Luk, Xi Luo, Jun Lv, Julie A. Lynch, Valeriya Lyssenko, Shiro Maeda, Vasiliki Mamakou, Sohail Rafik Mansuri, Koichi Matsuda, Thomas Meitinger, Andres Metspalu, Huan Mo, Andrew D. Morris, Jerry L. Nadler, Michael A. Nalls, Uma Nayak, Ioanna Ntalla, Yukinori Okada, Lorena Orozco, Sanjay R. Patel, Snehal Patil, Pei Pei, Mark A Pereira, Annette Peters, Fraser J. Pirie, Hannah G. Polikowsky, Bianca Porneala, Gauri Prasad, Laura J. Rasmussen-Torvik, Alexander P. Reiner, Michael Roden, Rebecca Rohde, Katheryn Roll, Charumathi Sabanayagam, Kevin Sandow, Alagu Sankareswaran, Naveed Sattar, Sebastian Schönherr, Mohammad Shahriar, Botong Shen, Jinxiu Shi, Dong Mun Shin, Nobuhiro Shojima, Jennifer A. Smith, Wing Yee So, Alena Stančáková, Valgerdur Steinthorsdottir, Adrienne M. Stilp, Konstantin Strauch, Kent D. Taylor, Barbara Thorand, Unnur Thorsteinsdottir, Brian Tomlinson, Tam C. Tran, Fuu-Jen Tsai, Jaakko Tuomilehto, Teresa Tusie-Luna, Miriam S. Udler, Adan Valladares-Salgado, Rob M. van Dam, Jan B. van Klinken, Rohit Varma, Niels Wacher-Rodarte, Eleanor Wheeler, Ananda R. Wickremasinghe, Ko Willems van Dijk, Daniel R. Witte, Chittaranjan S. Yajnik, Ken Yamamoto, Kenichi Yamamoto, Kyungheon Yoon, Canqing Yu, Jian-Min Yuan, Salim Yusuf, Matthew Zawistowski, Liang Zhang, Wei Zheng, VA Million Veteran Program, AMED GRIFIN Diabetes Initiative Japan, Biobank Japan Project, Penn Medicine BioBank, Regeneron Genetics Center, eMERGE Consortium, International Consortium for Blood Pressure (ICBP), Meta-Analyses of Glucose and Insulin-Related Traits Consortium (MAGIC), Leslie J Raffel, Michiya Igase, Eli Ipp, Susan Redline, Yoon Shin Cho, Lars Lind, Michael A. Province, Myriam Fornage, Craig L. Hanis, Erik Ingelsson, Alan B. Zonderman, Bruce M. Psaty, Ya-Xing Wang, Charles N. Rotimi, Diane M. Becker, Fumihiko Matsuda, Yongmei Liu, Mitsuhiro Yokota, Sharon L. R. Kardia, Patricia A. Peyser, James S. Pankow, James C. Engert, Amélie Bonnefond, Philippe Froguel, James G. Wilson, Wayne H. H. Sheu, Jer-Yuarn Wu, M. Geoffrey Hayes, Ronald C. W. Ma, Tien-Yin Wong, Dennis O. Mook-Kanamori, Tiinamaija Tuomi, Giriraj R. Chandak, Francis S. Collins, Dwaipayan Bharadwaj, Guillaume Paré, Michèle M. Sale, Habibul Ahsan, Ayesha A. Motala, Xiao-Ou Shu, Kyong-Soo Park, J Wouter Jukema, Miguel Cruz, Yii-Der Ida Chen, Stephen S. Rich, Roberta McKean-Cowdin, Harald Grallert, Ching-Yu Cheng, Mohsen Ghanbari, E-Shyong Tai, Josee Dupuis, Norihiro Kato, Markku Laakso, Anna Köttgen, Woon-Puay Koh, Donald W. Bowden, Colin N. A. Palmer, Jaspal S. Kooner, Charles Kooperberg, Simin Liu, Kari E. North, Danish Saleheen, Torben Hansen, Oluf Pedersen, Nicholas J. Wareham, Juyoung Lee, Bong-Jo Kim, Iona Y. Millwood, Robin G. Walters, Kari Stefansson, Mark O. Goodarzi, Karen L. Mohlke, Claudia Langenberg, Christopher A. Haiman, Ruth J. F. Loos, Jose C. Florez, Daniel J. Rader, Marylyn D. Ritchie, Sebastian Zöllner, Reedik Mägi, Joshua C. Denny, Toshimasa Yamauchi, Takashi Kadowaki, John C. Chambers, Maggie C. Y. Ng, Xueling Sim, Jennifer E. Below, Philip S. Tsao, Kyong-Mi Chang, Mark I. McCarthy, James B. Meigs, Anubha Mahajan, Cassandra N. Spracklen, Josep M. Mercader, Michael Boehnke, Jerome I. Rotter, Marijana Vujkovic, Benjamin F. Voight, Andrew P. Morris, Eleftheria Zeggini

**Affiliations:** Centre for Genetics and Genomics Versus Arthritis, Centre for Musculoskeletal Research, Division of Musculoskeletal and Dermatological Sciences, The University of Manchester, Manchester, UK; Department of Diabetes and Metabolic Diseases, Graduate School of Medicine, The University of Tokyo, Tokyo, Japan; Department of Statistical Genetics, Osaka University Graduate School of Medicine, Suita, Japan; Institute of Translational Genomics, Helmholtz Zentrum München, German Research Center for Environmental Health, Neuherberg, Germany.; Center for Precision Health Research, National Human Genome Research Institute; National Institutes of Health, Bethesda, MD, USA. British Heart Foundation Cardiovascular Epidemiology Unit, Department of Public Health and Primary Care, University of Cambridge, Cambridge, UK; Heart and Lung Research Institute, University of Cambridge, Cambridge, UK; Department of Biostatistics and Center for Statistical Genetics, University of Michigan, Ann Arbor, MI, USA; Department of Epidemiology, School of Public Health, Nanjing Medical University, Nanjing City, China; Corporal Michael J Crescenz VA Medical Center, Philadelphia, PA, USA; Department of Systems Pharmacology and Translational Therapeutics, University of Pennsylvania Perelman School of Medicine, Philadelphia, PA, USA; Department of Genetics, University of Pennsylvania Perelman School of Medicine, Philadelphia, PA, USA; Programs in Metabolism and Medical and Population Genetics, Broad Institute of Harvard and MIT, Cambridge, MA, USA; Diabetes Unit and Center for Genomic Medicine, Massachusetts General Hospital, Boston, MA, USA; Consejo Nacional de Ciencia y Tecnología (CONACYT), Instituto Nacional de Ciencias Médicas y Nutrición Salvador Zubirán, Mexico City, Mexico; Department of Genome Informatics, Graduate School of Medicine, The University of Tokyo, Tokyo, Japan; Integrated Frontier Research for Medical Science Division, Institute for Open and Transdisciplinary Research Initiatives, Osaka University, Suita, Japan; Laboratory for Systems Genetics, RIKEN Center for Integrative Medical Sciences, Kanagawa, Japan; The Charles Bronfman Institute for Personalized Medicine, Icahn School of Medicine at Mount Sinai, New York, NY, USA; Department of Medicine, Vanderbilt University Medical Center, Nashville, TN, USA; Estonian Genome Centre, Institute of Genomics, University of Tartu, Tartu, Estonia; Nuffield Department of Population Health, University of Oxford, Oxford, UK; Medical Research Council Population Health Research Unit, University of Oxford, Oxford, UK; The Institute for Translational Genomics and Population Sciences, Department of Pediatrics, The Lundquist Institute for Biomedical Innovation (formerly Los Angeles Biomedical Research Institute) at Harbor-UCLA Medical Center, Torrance, CA, USA; Department of Epidemiology and Biostatistics, Imperial College London, London, UK; Department of Cardiology, Ealing Hosptial, London NorthWest Healthcare NHS Trust, Middlesex, UK; Division of Genome Science, Department of Precision Medicine, National Institute of Health, Cheongju-si, South Korea; Department of Epidemiology, Gillings School of Global Public Health, University of North Carolina at Chapel Hill, Chapel Hill, NC, USA; Department of Gene Diagnostics and Therapeutics, Research Institute, National Center for Global Health and Medicine, Tokyo, Japan; Institute of Epidemiology, Helmholtz Zentrum Munchen, German Research Center for Environmental Health, Neuherberg, Germany; Department of Medicine, McMaster University, Hamilton, ON, Canada; Population Health Research Institute, Hamilton Health Sciences and McMaster University, Hamilton, ON, Canada; Public Health Informatics Unit, Department of Integrated Health Sciences, Nagoya University Graduate School of Medicine, Nagoya, Japan; MRC Epidemiology Unit, Institute of Metabolic Science, University of Cambridge School of Clinical Medicine, Cambridge, UK; Department of Health Data Science, University of Liverpool, Liverpool, UK; Division of Translational Medicine and Human Genetics, University of Pennsylvania, Philadelphia, PA, USA; Department of Epidemiology, Brown University School of Public Health, Providence, RI, USA; Department of Anthropology, University of Toronto at Mississsauga, Mississauga, ON, Canada; Saw Swee Hock School of Public Health, National University of Singapore and National University Health System, Singapore, Singapore; Department of Epidemiology, School of Public Health, University of Michigan, Ann Arbor, MI, USA; Center for Genomic Medicine, Kyoto University Graduate School of Medicine, Kyoto, Japan; deCODE Genetics, Amgen Inc., Reykjavik, Iceland; Novo Nordisk Foundation Center for Basic Metabolic Research, Faculty of Health and Medical Sciences, University of Copenhagen, Copenhagen, Denmark; Department of Biostatistics, Harvard University, Boston, MA, USA; Division of Sleep and Circadian Disorders, Brigham and Women’s Hospital, Boston, MA, USA; Department of Medicine, Harvard University, Boston, MA, USA; Institute of Genetic Epidemiology, Department of Data Driven Medicine, Faculty of Medicine and Medical Center, University of Freiburg, Freiburg, Germany; Department of Epidemiology, Human Genetics, and Environmental Sciences, The University of Texas Health Science Center at Houston School of Public Health, Houston, TX, USA; German Center for Diabetes Research (DZD), Neuherberg, Germany; Research Unit of Molecular Epidemiology, Helmholtz Zentrum München, German Research Center for Environmental Health, Neuherberg, Germany; Department of Population and Public Health Sciences, Keck School of Medicine of USC, Los Angeles, CA, USA; Department of Cardiology, Leiden University Medical Center, Leiden, The Netherlands; Section of Gerontology and Geriatrics, Department of Internal Medicine, Leiden University Medical Center, Leiden, The Netherlands.; Department of Internal Medicine, Seoul National University Hospital, Seoul, South Korea; Division of Epidemiology, Department of Medicine, Institute for Medicine and Public Health, Vanderbilt Genetics Institute, Vanderbilt University Medical Center, Nashville, TN, USA; Nuffield Department of Surgical Sciences, University of Oxford, Oxford, UK; Institute for Population and Precision Health (IPPH), Biological Sciences Division, The University of Chicago, Chicago, IL, USA; Department of Public Health Sciences and Center for Public Health Genomics, University of Virginia School of Medicine, Charlottesville, VA, USA; Genomic Research on Complex Diseases (GRC-Group), CSIR-Centre for Cellular and Molecular Biology (CSIR-CCMB), Hyderabad, India; Department of Medicine and Therapeutics, The Chinese University of Hong Kong, Hong Kong, China; Chinese University of Hong Kong-Shanghai Jiao Tong University Joint Research Centre in Diabetes Genomics and Precision Medicine, The Chinese University of Hong Kong, Hong Kong, China; Institute of Data Science, Korea University, Seoul, South Korea; Division of Endocrinology, Metabolism, and Molecular Medicine, Department of Medicine, Northwestern University Feinberg School of Medicine, Chicago, IL, USA; Department of Health and Biomedical Informatics, Northwestern University Feinberg School of Medicine, Chicago, IL, USA; Institute of Biomedical Sciences, Academia Sinica, Taipei, Taiwan; Department of Genetics, University of North Carolina at Chapel Hill, Chapel Hill, NC, USA; Department of Human Genetics, Wellcome Sanger Institute, Wellcome Genome Campus, Hinxton, UK; Laboratory of Neurogenetics, National Institute on Aging, National Institutes of Health, Bethesda, MD, USA; Department of Medicine, Johns Hopkins University School of Medicine, Baltimore, MD, USA; Center for Research on Genomics and Global Health, National Human Genome Research Institute, National Institutes of Health, Bethesda, MD, USA; Cardiovascular Health Research Unit, Department of Medicine, University of Washington, Seattle, WA, USA; Division of Academics, Ochsner Health, New Orleans, LA, USA; Division of Statistical Genomics, Washington University School of Medicine, St. Louis, MO, USA; Department of Research and Evaluation, Division of Biostatistics Research, Kaiser Permanente of Southern California, Pasadena, CA, USA; Department of Biomedical Science, Hallym University, Chuncheon, South Korea; Harvard Medical School, Boston, MA, USA; Metabolic Research Laboratories, Wellcome Trust-Medical Research Council Institute of Metabolic Science, Department of Clinical Biochemistry, University of Cambridge, Cambridge, UK; Department of Epidemiology, University of Groningen, University Medical Centre Groningen, Groningen, The Netherlands; Department of Bioinformatics, Isfahan University of Medical Sciences, Isfahan, Iran; Department of Medicine, Division of Endocrinology, Diabetes and Metabolism, Cedars-Sinai Medical Center, Los Angeles, CA, USA; Department of Nutrition, Gillings School of Global Public Health, University of North Carolina at Chapel Hill, Chapel Hill, NC, USA; Unidad de Investigación en Enfermedades Metabólicas and Departamento de Endocrinología y Metabolismo, Instituto Nacional de Ciencias Médicas y Nutrición Salvador Zubirán, Mexico City, Mexico; Steno Diabetes Center Copenhagen, Herlev, Denmark; The Bioinformatics Center, Department of Biology, University of Copenhagen, Copenhagen, Denmark; Department of Health Research Methods, Evidence, and Impact, McMaster University, Hamilton, ON, Canada; Department of Epidemiology and Prevention, Division of Public Health Sciences, Wake Forest School of Medicine, Winston-Salem, NC, USA; Institute of Regional Health Research, University of Southern Denmark, Odense, Denmark; Department of Clinical Biochemistry, Vejle Hospital, Vejle, Denmark; Department of Medicine, Division of Endocrinology and Diabetes, Keck School of Medicine of USC, Los Angeles, CA, USA; Department of Internal Medicine, University of Michigan, Ann Arbor, MI, USA; British Heart Foundation Centre of Research Excellence, School of Clinical Medicine, Addenbrooke’s Hospital, University of Cambridge, Cambridge, UK; Health Data Research UK Cambridge, Wellcome Genome Campus and University of Cambridge, Hinxton, UK; National Institute for Health and Care Research (NIHR) Blood and Transplant Unit (BTRU) in Donor Health and Behaviour, Heart and Lung Research Institute, University of Cambridge, Cambridge, UK; Inserm U1283, CNRS UMR 8199, European Genomic Institute for Diabetes (EGID), Institut Pasteur de Lille, Lille University Hospital, Lille, France; University of Lille, Lille, France; Li Ka Shing Institute of Health Sciences, The Chinese University of Hong Kong, Hong Kong, China; Hong Kong Institute of Diabetes and Obesity, The Chinese University of Hong Kong, Hong Kong, China; Singapore Eye Research Institute, Singapore National Eye Centre, Singapore, Singapore; Exeter Centre of Excellence in Diabetes (ExCEeD), Exeter Medical School, University of Exeter, Exeter, UK; Wellcome Sanger Institute, Wellcome Genome Campus, Hinxton, UK; Department of Biostatistics and Data Science, Wake Forest School of Medicine, Winston-Salem, NC, USA; Division of Endocrinology and Metabolism, Department of Internal Medicine, National Taiwan University Hospital, Taipei, Taiwan; Institute of Epidemiology and Preventive Medicine, National Taiwan University, Taipei, Taiwan; Department of Medicine, University of Vermont, Colchester, VT, USA; Section on Endocrinology and Metabolism, Department of Internal Medicine, Wake Forest School of Medicine, Winston-Salem, NC, USA; Department of Medicine, Faculty of Medicine, University of Kelaniya, Ragama, Sri Lanka; Department of Nutrition and Dietetics, Harokopio University of Athens, Athens, Greece; Center for Genomics and Personalized Medicine Research, Wake Forest School of Medicine, Winston-Salem, NC, USA; Carolina Population Center, University of North Carolina at Chapel Hill, Chapel Hill, NC, USA; Department of Nephrology and Medical Intensive Care Medicine, Charité Universitätsmedizin Berlin, Berlin, Germany; Department of Nephrology and Hypertension, Friedrich-Alexander-Universität Erlangen-Nürnberg, Erlangen, Germany; Department of Biostatistics, University of Washington, Seattle, WA, USA; California Pacific Medical Center Research Institute, San Francisco, CA, USA; Laboratory of Epidemiology and Population Sciences, National Institute on Aging, National Institutes of Health, Baltimore, MD, USA; Institute of Mathematics and Statistics, University of Tartu, Tartu, Estonia; Robertson Centre for Biostatistics, University of Glasgow, Glasgow, UK; Department of Epidemiology, Erasmus MC University Medical Center, Rotterdam, The Netherlands; Genetics of Complex Traits, University of Exeter Medical School, University of Exeter, Exeter, UK; Department of Internal Medicine, Wake Forest School of Medicine, Winston-Salem, NC, USA; Department of Medicine, Division of Endocrinology and Metabolism, Lundquist Research Institute at Harbor-UCLA Medical Center, Torrance, CA, USA; Department of Public Health and Caring Sciences, Uppsala University, Uppsala, Sweden; Centro de Estudios en Diabetes, Unidad de Investigacion en Diabetes y Riesgo Cardiovascular, Centro de Investigacion en Salud Poblacional, Instituto Nacional de Salud Publica, Mexico City, Mexico; Department of Laboratory Medicine and Pathology, University of Minnesota, Minneapolis, MN, USA; Genomics and Computational Biology Graduate Group, University of Pennsylvania Perelman School of Medicine, Philadelphia, PA, USA; University of Exeter Medical School, University of Exeter, Exeter, UK; Institute for Clinical Diabetology, German Diabetes Center, Leibniz Center for Diabetes Research at Heinrich Heine University Düsseldorf, Düsseldorf, Germany; Department of Endocrinology and Diabetology, Medical Faculty and University Hospital Dusseldorf, Heinrich Heine University Düsseldorf, Düsseldorf, Germany; Laboratory for Genomics of Diabetes and Metabolism, RIKEN Center for Integrative Medical Sciences, Kanagawa, Japan; Department of Biostatistics, Gillings School of Global Public Health, University of North Carolina at Chapel Hill, Chapel Hill, NC, USA; Department of Internal Medicine, Diabetes and Metabolism Research Center, The Ohio State University Wexner Medical Center, Columbus, OH, USA; Center for Global Cardiometabolic Health, Brown University, Providence, RI, USA; Shanghai-MOST Key Laboratory of Health and Disease Genomics, Chinese National Human Genome Center at Shanghai (CHGC) and Shanghai Institute for Biomedical and Pharmaceutical Technologies (SIBPT), Shanghai, China; Division of Endocrine and Metabolism, Tri-Service General Hospital Songshan Branch, Taipei, Taiwan; School of Medicine, National Defense Medical Center, Taipei, Taiwan; Division of Genome Science, Department of Precision Medicine, National Institute of Health, Cheongju-si, Korea; Section of Endocrinology and Metabolism, Department of Medicine, Taipei Veterans General Hospital, Taipei, Taiwan; School of Medicine, National Yang Ming Chiao Tung University, Taipei, Taiwan; Department of Environmental and Preventive Medicine, Jichi Medical University School of Medicine, Shimotsuke, Japan; University of Chicago Research Bangladesh, Dhaka, Bangladesh; Institute of Molecular and Clinical Ophthalmology Basel, Basel, Switzerland; Center for Clinical Research and Prevention, Bispebjerg and Frederiksberg Hospital, Frederiksberg, Denmark; Faculty of Health and Medical Sciences, University of Copenhagen, Copenhagen, Denmark; Faculty of Medicine, Aalborg University, Aalborg, Denmark; Department of Clinical Diabetes, Endocrinology and Metabolism, Department of Translational Research and Cellular Therapeutics, City of Hope, Duarte, CA, USA; Department of Public Health, Faculty of Medicine, University of Kelaniya, Ragama, Sri Lanka; Department of Clinical Gene Therapy, Osaka University Graduate School of Medicine, Osaka, Japan; Department of Geriatric and General Medicine, Graduate School of Medicine, Osaka University, Osaka, Japan; Division of General Internal Medicine and Geriatrics, Department of Medicine, Northwestern University Feinberg School of Medicine, Chicago, IL, USA; Center for Health Information Partnerships, Institute for Public Health and Medicine, Northwestern University Feinberg School of Medicine, Chicago, IL, USA; Genome Institute of Singapore, Agency for Science, Technology and Research, Singapore, Singapore; Department of Molecular Cell Biology, Sungkyunkwan University School of Medicine, Suwon, South Korea; Institute of Genetic Epidemiology, Medical University of Innsbruck, Innsbruck, Austria; Institute of Clinical Medicine, Internal Medicine, University of Eastern Finland and Kuopio University Hospital, Kuopio, Finland; Department of Medicine, University of Colorado Denver, Anschutz Medical Campus, Aurora, CO, USA; VA Salt Lake City Health Care System, Salt Lake City, UT, USA; Department of Internal Medicine, University of Utah School of Medicine, Salt Lake City, UT, USA; Severance Biomedical Science Institute and Department of Internal Medicine, Yonsei University College of Medicine, Seoul, South Korea; Department of Medicine, Samsung Medical Center, Sungkyunkwan University School of Medicine, Seoul, South Korea; USC-Office of Population Studies Foundation Inc., University of San Carlos, Cebu City, Philippines; Department of Medicine, Harvard Medical School, Boston, MA, USA; Division of General Internal Medicine, Massachusetts General Hospital, Boston, MA, USA; Department of Epidemiology and Biostatistics, School of Public Health, Peking University, Beijing, China; Peking University Center for Public Health and Epidemic Preparedness and Response, Beijing, China; Department of Clinical Epidemiology, Leiden University Medical Center, Leiden, The Netherlands; Wellcome Centre for Human Genetics, Nuffield Department of Medicine, University of Oxford, Oxford, UK; Program in Medical and Population Genetics, Broad Institute, Cambridge, MA, USA; Big Data Institute, Li Ka Shing Centre For Health Information and Discovery, University of Oxford, Oxford, UK; Department of Clinical Medicine, Faculty of Health and Medical Sciences, University of Copenhagen, Copenhagen, Denmark; Department of Biostatistics, Boston University School of Public Health, Boston, MA, USA; Department of Medicine, Yong Loo Lin School of Medicine, National University of Singapore and National University Health System, Singapore, Singapore; McDonnell Genome Institute, Washington University School of Medicine, St Louis, MO, USA; Department of Medicine, Division of Genomics and Bioinformatics, Washington University School of Medicine, St Louis, MO, USA; Department of Biostatistics and Data Science, The University of Texas Health Science Center at Houston School of Public Health, Houston,, USA; Department of Clinical Sciences, Diabetes and Endocrinology, Lund University Diabetes Centre, Malmö, Sweden; Department of Clinical Science, Center for Diabetes Research, University of Bergen, Bergen, Norway; Department of Advanced Genomic and Laboratory Medicine, Graduate School of Medicine, University of the Ryukyus, Okinawa, Japan; Division of Clinical Laboratory and Blood Transfusion, University of the Ryukyus Hospital, Okinawa, Japan; Dromokaiteio Psychiatric Hospital, National and Kapodistrian University of Athens, Athens, Greece; Computational Biology and Medical Sciences, Graduate School of Frontier Sciences, The University of Tokyo, Tokyo, Japan; Institute of Human Genetics, Helmholtz Zentrum München, German Research Center for Environmental Health, Neuherberg, Germany; Institute of Human Genetics, Technical University Munich, Munich, Germany; German Centre for Cardiovascular Research (DZHK), Partner Site Munich Heart Alliance, Munich, Germany; The Usher Institute to the Population Health Sciences and Informatics, University of Edinburgh, Edinburgh, UK; Department of Medicine and Pharmacology, New York Medical College, Valhalla, NY, USA; Data Tecnica International LLC, Glen Echo, MD, USA; Center for Alzheimer’s and Related Dementias, National Institutes of Health, Bethesda, MD, USA; William Harvey Research Institute, Barts and The London School of Medicine and Dentistry, Queen Mary University of London, London, UK; Laboratory of Statistical Immunology, Immunology Frontier Research Center (WPI-IFReC), Osaka University, Suita, Japan; Instituto Nacional de Medicina Genómica, Mexico City, Mexico; Division of Pulmonary, Allergy, and Critical Care Medicine, Department of Medicine, University of Pittsburgh, Pittsburgh, PA, USA; Division of Epidemiology and Community Health, School of Public Health, University of Minnesota, Minneapolis, MN, USA; Institute for Medical Information Processing, Biometry and Epidemiology, Ludwig Maximilians Universität München, Munich, Germany; Department of Diabetes and Endocrinology, Nelson R Mandela School of Medicine, College of Health Sciences, University of KwaZulu-Natal, Durban, South Africa; Academy of Scientific and Innovative Research, CSIR-Human Resource Development Campus, Ghaziabad, India; Genomics and Molecular Medicine Unit, CSIR-Institute of Genomics and Integrative Biology, New Delhi, India; Department of Preventive Medicine, Northwestern University Feinberg School of Medicine, Chicago, IL, USA; Fred Hutchinson Cancer Research Center, Seattle, WA, USA; Ophthalmology and Visual Sciences Academic Clinical Program (Eye ACP), Duke-NUS Medical School, Singapore, Singapore; Department of Ophthalmology, Yong Loo Lin School of Medicine, National University of Singapore and National University Health System, Singapore, Singapore; Institute of Cardiovascular and Medical Sciences, University of Glasgow, Glasgow, UK; Survey Research Center, Institute for Social Research, University of Michigan, Ann Arbor, MI, USA; Institute of Genetic Epidemiology, Helmholtz Zentrum Munchen, German Research Center for Environmental Health, Neuherberg, Germany; Institute for Medical Biostatistics, Epidemiology and Informatics (IMBEI), University Medical Center, Johannes Gutenberg University, Mainz, Germany; Chair of Genetic Epidemiology, Institute of Medical Information Processing, Biometry, and Epidemiology, Faculty of Medicine, Ludwig Maximilians Universität München, Munich, Germany; Faculty of Medicine, University of Iceland, Reykjavik, Iceland; Faculty of Medicine, Macau University of Science and Technology, Macau, China; Department of Medical Genetics and Medical Research, China Medical University Hospital, Taichung, Taiwan; Population Health Unit, Finnish Institute for Health and Welfare, Helsinki, Finland, Finnish Institute for Health and Welfare, Helsinki, Finland; National School of Public Health, Madrid, Spain; Department of Public Health, University of Helsinki, Helsinki, Finland; Diabetes Research Group, King Abdulaziz University, Jeddah, Saudi Arabia; Unidad de Biología Molecular y Medicina Genómica, Instituto Nacional de Ciencias Médicas y Nutrición Salvador Zubirán, Mexico City, Mexico; Departamento de Medicina Genómica y Toxiología Ambiental, Instituto de Investigaciones Biomédicas, UNAM, Mexico City, Mexico; Unidad de Investigacion Medica en Bioquimica, Hospital de Especialidades, Centro Medico Nacional Siglo XXI, Instituto Mexicano del Seguro Social, Mexico City, Mexico; Einthoven Laboratory for Experimental Vascular Medicine, Leiden University Medical Center, Leiden, The Netherlands; Department of Human Genetics, Leiden University Medical Center, Leiden, The Netherlands; Department of Clinical Chemistry, Laboratory of Genetic Metabolic Disease, Amsterdam University Medical Center, Amsterdam, The Netherlands; Southern California Eye Institute, CHA Hollywood Presbyterian Hospital, Los Angeles, CA, USA; Unidad de Investigación Médica en Epidemiologia Clinica, Hospital de Especialidades, Centro Medico Nacional Siglo XXI, Instituto Mexicano del Seguro Social, Mexico City, Mexico; Department of Internal Medicine, Division of Endocrinology, Leiden University Medical Center, Leiden, The Netherlands; Department of Public Health, Aarhus University, Aarhus, Denmark; Danish Diabetes Academy, Odense, Denmark; Diabetology Research Centre, King Edward Memorial Hospital and Research Centre, Pune, India; Department of Medical Biochemistry, Kurume University School of Medicine, Kurume, Japan; Department of Pediatrics, Osaka University Graduate School of Medicine, Suita, Japan; Division of Cancer Control and Population Sciences, UPMC Hillman Cancer Center, University of Pittsburgh, Pittsburgh, PA, USA; Department of Epidemiology, Graduate School of Public Health, University of Pittsburgh, Pittsburgh, PA, USA; Department of Pediatrics, Division of Genetic and Genomic Medicine, UCI Irvine School of Medicine, Irvine, CA, USA; Department of Anti-Aging Medicine, Ehime University Graduate School of Medicine, Ehime, Japan; Division of Pulmonary, Critical Care, and Sleep Medicine, Beth Israel Deaconess Medical Center, Boston, MA, USA; Department of Medical Sciences, Uppsala University, Uppsala, Sweden; Institute of Molecular Medicine, The University of Texas Health Science Center at Houston School of Public Health, Houston, TX, USA; Human Genetics Center, University of Texas Health Science Center at Houston, Houston, TX, US; Department of Medicine, Stanford University School of Medicine, Stanford, CA, USA; Department of Medical Sciences, Molecular Epidemiology and Science for Life Laboratory, Uppsala University, Uppsala, Sweden; Department of Epidemiology, University of Washington, Seattle, WA, USA; Department of Health Systems and Population Health, University of Washington, Seattle, WA, USA; Beijing Institute of Ophthalmology, Ophthalmology and Visual Sciences Key Laboratory, Beijing Tongren Hospital, Capital Medical University, Beijing, China; Department of Medicine, Division of Cardiology, Duke University School of Medicine, Durham, NC, USA; Kurume University School of Medicine, Kurume, Japan; Department of Medicine, McGill University, Montreal, QC, Canada; Department of Human Genetics, McGill University, Montreal, QC, Canada; Department of Metabolism, Digestion and Reproduction, Imperial College London, London, UK; Department of Physiology and Biophysics, University of Mississippi Medical Center, Jackson, MS, USA; Division of Endocrinology and Metabolism, Department of Medicine, Taichung Veterans General Hospital, Taichung, Taiwan; Center for Genetic Medicine, Northwestern University Feinberg School of Medicine, Chicago, IL, USA; Department of Anthropology, Northwestern University, Evanston, IL, USA; Department of Endocrinology, Helsinki University Hospital, Helsinki, Finland; Institute for Molecular Medicine Finland (FIMM), University of Helsinki, Helsinki, Finland; Folkhalsan Research Center, Helsinki, Finland; Lund University Diabetes Centre, Malmö, Sweden; Systems Genomics Laboratory, School of Biotechnology, Jawaharlal Nehru University, New Delhi, India; Department of Pathology and Molecular Medicine, McMaster University, Hamilton, ON, Canada; Department of Molecular Medicine and Biopharmaceutical Sciences, Graduate School of Convergence Science and Technology, Seoul National University, Seoul, South Korea; Netherlands Heart Institute, Utrecht, The Netherlands; Center for Public Health Genomics, University of Virginia School of Medicine, Charlottesville, VA, USA; Research Unit of Molecular Epidemiology, Helmholtz Zentrum München, German Research Center for Environmental Health, Munich, Germany; Duke-NUS Medical School, Singapore, Singapore; Department of Epidemiology, Biostatistics and Occupational Health, McGill University, Montreal, QC, Canada; Singapore Institute for Clinical Sciences, Agency for Science Technology and Research (A*STAR), Singapore, Singapore; Healthy Longevity Translational Research Programme, Yong Loo Lin School of Medicine, National University of Singapore, Singapore, Singapore; Center for Diabetes Research, Wake Forest School of Medicine, Winston-Salem, NC, USA; Department of Biochemistry, Wake Forest School of Medicine, Winston-Salem, NC, USA; Pat Macpherson Centre for Pharmacogenetics and Pharmacogenomics, University of Dundee, Dundee, UK; Imperial College Healthcare NHS Trust, Imperial College London, London, UK; MRC-PHE Centre for Environment and Health, Imperial College London, London, UK; National Heart and Lung Institute, Imperial College London, London, UK; Department of Medicine, Brown University Alpert School of Medicine, Providence, RI, USA; Department of Medicine, Columbia University Irving Medical Center, New York, NY, USA; Department of Cardiology, Columbia University Irving Medical Center, New York, NY, USA; Center for Non-Communicable Diseases, Karachi, Pakistan; Computational Medicine, Berlin Institute of Health at Charité Universitätsmedizin, Berlin, Germany; Precision Healthcare University Research Institute, Queen Mary University of London, London, UK; Department of Preventive Medicine, Keck School of Medicine of USC, Los Angeles, CA, USA; The Mindich Child Health and Development Institute, Ichan School of Medicine at Mount Sinai, New York, NY, USA; Division of Translational Medicine and Therapeutics, Department of Medicine, University of Pennsylvania Perelman School of Medicine, Philadelphia, PA, USA; Institute for Translational Medicine and Therapeutics, University of Pennsylvania Perelman School of Medicine, Philadelphia,, USA; Department of Pediatrics, University of Pennsylvania Perelman School of Medicine, Philadelphia, PA, USA; Center for Precision Medicine, University of Pennsylvania Perelman School of Medicine, Philadelphia, PA, USA; Institute for Biomedical Informatics, University of Pennsylvania Perelman School of Medicine, Philadelphia, PA, USA; Department of Psychiatry, University of Michigan, Ann Arbor, MI, USA; All of Us Research Program, National Institutes of Health, Bethesda, MD, USA; Toranomon Hospital, Tokyo, Japan; Lee Kong Chian School of Medicine, Nanyang Technological University, Singapore, Singapore; Vanderbilt Genetics Institute, Division of Genetic Medicine, Vanderbilt University Medical Center, Nashville, TN, USA; VA Palo Alto Health Care System, Palo Alto, CA, USA; Stanford Cardiovascular Institute, Stanford University School of edicine, Stanford, CA, USA; Department of Medicine, University of Pennsylvania Perelman School of Medicine, Philadelphia, PA, USA; Oxford Centre for Diabetes, Endocrinology and Metabolism, Radcliffe Department of Medicine, University of Oxford, Oxford, UK; Oxford NIHR Biomedical Research Centre, Churchill Hosptial, Oxford University Hospitals NHS Foundation Trust, Oxford, UK; Department of Biostatistics and Epidemiology, University of Massachusetts Amherst, Amherst, MA, USA; Division of Epidemiology, Department of Biostatistics, Epidemiology and Informatics, University of Pennsylvania Perelman School of Medicine, Philadelphia, PA, USA; TUM School of Medicine, Technical University of Munich and Klinikum Rechts der Isar, Munich, Germany

## Abstract

Type 2 diabetes (T2D) is a heterogeneous disease that develops through diverse pathophysiological processes. To characterise the genetic contribution to these processes across ancestry groups, we aggregate genome-wide association study (GWAS) data from 2,535,601 individuals (39.7% non-European ancestry), including 428,452 T2D cases. We identify 1,289 independent association signals at genome-wide significance (P<5×10^−8^) that map to 611 loci, of which 145 loci are previously unreported. We define eight non-overlapping clusters of T2D signals characterised by distinct profiles of cardiometabolic trait associations. These clusters are differentially enriched for cell-type specific regions of open chromatin, including pancreatic islets, adipocytes, endothelial, and enteroendocrine cells. We build cluster-specific partitioned genetic risk scores (GRS) in an additional 137,559 individuals of diverse ancestry, including 10,159 T2D cases, and test their association with T2D-related vascular outcomes. Cluster-specific partitioned GRS are more strongly associated with coronary artery disease and end-stage diabetic nephropathy than an overall T2D GRS across ancestry groups, highlighting the importance of obesity-related processes in the development of vascular outcomes. Our findings demonstrate the value of integrating multi-ancestry GWAS with single-cell epigenomics to disentangle the aetiological heterogeneity driving the development and progression of T2D, which may offer a route to optimise global access to genetically-informed diabetes care.

---

Diabetes mellitus is a huge public health burden, with an estimated prevalence of 537 million adults worldwide in 2021, of whom >90% are affected by type 2 diabetes (T2D)^1^. The biological processes through which T2D and its primary disabling complications of vascular and renal disease develop are diverse, and include insulin secretion and insulin resistance. This aetiological heterogeneity leads to substantial variability in patient phenotypes, including age of disease onset, manifestation of disease complications, and response to management strategies^2, 3^. Whilst environment and lifestyle are well-established risk factors for T2D, the disease heritability has been estimated to be 69% amongst individuals in the age range of 35 to 60 years^4^. The largest previous genome-wide association studies (GWAS) of T2D have identified >500 risk loci^5, 6^, which demonstrated variable patterns of association with clinical features mediated via effector genes acting through distinct molecular mechanisms that are often cell-type specific^7, 8^. Through the newly-established Type 2 Diabetes Global Genomics Initiative, we present findings from the largest T2D GWAS meta-analysis to date, comprising >2.5 million individuals of diverse ancestry and a near three-fold increase in effective sample size over previous efforts^5, 6^. We leverage the power afforded by this increased sample size with emerging single-cell functional genomics data derived from disease-relevant tissues to disentangle the aetiological heterogeneity of T2D. For the first time across multiple ancestry groups, we construct partitioned genetic risk scores (GRS)^9^ and assess their association with T2D-related macrovascular outcomes and progression to microvascular complications in individuals with T2D. Taken together, our findings showcase the value of analyses conducted in multiple ancestries to understand the pathophysiological processes driving development of T2D and risk of vascular complications, and advance global opportunities for clinical translation of T2D GWAS findings.

## Study overview

We assembled GWAS including 428,452 T2D cases and 2,107,149 controls (effective sample size of 1,246,658) (**Supplementary** Tables 1 and 2). We organised these GWAS into five groups representing individuals with similar genetic background and refer to these as “ancestry groups”. Specifically, we considered: a European ancestry group (EUR, 60.3% of the effective sample size); an East Asian ancestry group (EAS, 19.8%); an African ancestry group, including admixed African Americans (AFR, 10.7%); a Hispanic group, who share genetic similarity with populations from the Americas, Africa, and Europe (HIS, 5.9%); and a South Asian ancestry group (SAS, 3.3%). The GWAS in each ancestry group were imputed to reference panels from the Trans-Omics for Precision Medicine Program^10^ (42.1% of the effective sample size), Haplotype Reference Consortium^11^ (14.7%), 1000 Genomes Project^12, 13^ (11.8%), or whole-genome sequence data specific for and genetically similar to individuals in the study (31.4%). Subsequent association analyses accounted for study-level population stratification and relatedness, and adjusted for age and sex, where appropriate, and additional study-specific covariates (Supplementary Table 3**, Methods**). We analysed bi-allelic autosomal single nucleotide variants (SNVs) with minor allele frequency (MAF) ≥0.5% in at least one of the five ancestry groups from the 1000 Genomes Project^13^.

**Table 1.**
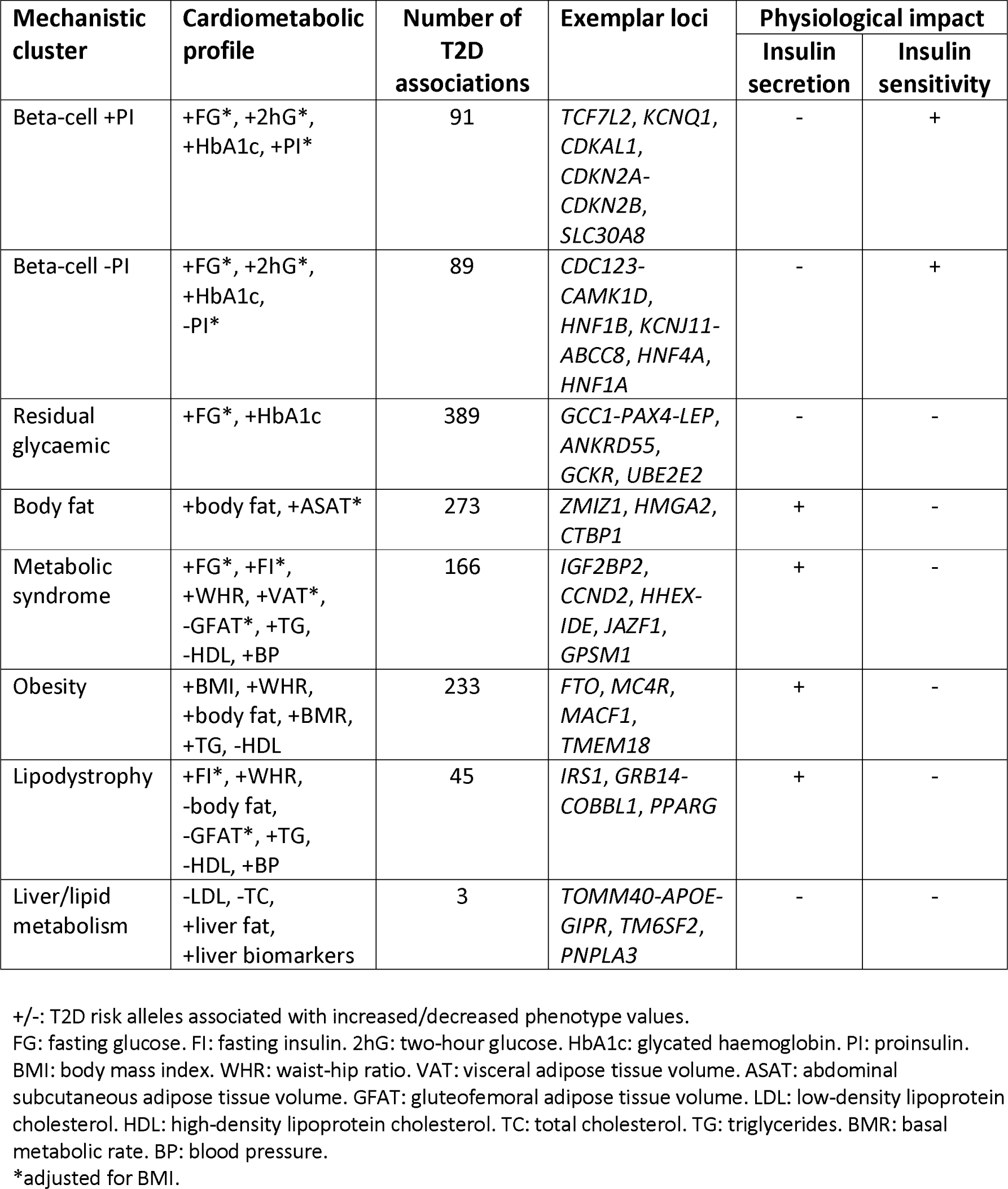
Cardiometabolic profile, exemplar loci, and physiological impact of index SNVs at T2D association signals allocated to eight mechanistic clusters.

## Discovery of 145 previously-unreported T2D loci

We aggregated association summary statistics across GWAS via multi-ancestry meta-regression, implemented in MR-MEGA^14^, which allows for allelic effect heterogeneity that is correlated with ancestry. MR-MEGA is not restricted to broad continental ancestry categories but represents ancestry as multidimensional and continuous axes of genetic variation, which better reflect the continuum of human genetic diversity and demographic history^15^. We included three axes of genetic variation as covariates that separated GWAS from different ancestry groups, but which also revealed finer-scale population differences within ancestry groups (Supplementary Figure 1**, Methods**). Accounting for ancestry-correlated allelic effect heterogeneity among GWAS in the meta-regression provided better control for residual structure than a fixed-effects meta-analysis, as measured by the genomic control inflation factor (**λ_GC_**=1.120 and **λ_GC_**=1.396, respectively). At the conventional genome-wide significance threshold (**P**<5×10^-8^), we identified 1,289 distinct T2D association signals that were represented by independent index SNVs that were not in linkage disequilibrium (LD), defined as **r**^2^<0.05 in all ancestry groups from the 1000 Genomes Project^13^ (Supplementary Figure 2, Supplementary Table 4**, Methods**). The 1,289 association signals mapped to 611 loci, of which 145 loci (23.7%) have not been previously reported in T2D GWAS. We observed multiple distinct association signals at 235 (38.5%) loci, of which 34 loci were represented by more than five index SNVs. At association signals mapping to loci not previously reported for T2D, index SNVs were predominantly common (MAF >5% in at least one ancestry group) with odds-ratios (ORs) <1.1 (Supplementary Figure 3).

## T2D association signals are enriched for ancestry-correlated heterogeneity

Previous multi-ancestry GWAS have demonstrated widespread heterogeneity in allelic effects at T2D association signals across ancestry groups^6, 16^. Such heterogeneity is not driven by variation in allele frequency between ancestry groups, but may reflect differences in LD structure, or interactions with environment or polygenic background. We took advantage of the meta-regression model to partition heterogeneity into an ancestry-correlated component explained by three axes of genetic variation, and a residual component reflecting differences in environmental exposures (that are not correlated with ancestry) and/or study design (Supplementary Table 5). We observed 127 (9.9%) independent T2D association signals with significant evidence for ancestry-correlated heterogeneity (**P_HET_**<3.9×10^-5^, Bonferroni correction for 1,289 signals). We would expect <1 signal to meet this threshold of significance, highlighting that ancestry-correlated heterogeneity is strongly enriched at T2D associations (one-sided binomial test **P**<2.2×10^-16^). In contrast, we observed significant evidence of residual heterogeneity at only 4 (0.3%) association signals (one-side binomial test **P**=0.031). These results therefore suggest that differences in allelic effects at index SNVs are more likely correlated with genetic ancestry than due to other factors that vary between GWAS.

We next sought to understand better the drivers of differences in allelic effects between GWAS at the 127 association signals with significant evidence of ancestry-correlated heterogeneity. To do this, we assessed the contribution of each of the three axes of genetic variation to heterogeneity (**Methods**). For 118 (92.9%) signals, allelic effect sizes were most strongly associated with the first two axes of genetic variation, which separate AFR, EAS, and EUR GWAS (Supplementary Figure 1, Supplementary Table 6). This may simply reflect power to detect heterogeneity because these three ancestry groups make the largest contributions to the effective sample size of the multi-ancestry meta-analysis. The magnitude and direction of the association of index SNVs with these two axes reflected differences in allelic effect size between AFR and EAS GWAS on the AFR-EAS axis, and AFR and EUR GWAS on the AFR-EUR axis (Supplementary Figure 4). The most significant evidence of ancestry-correlated heterogeneity was observed for the T2D association signal at the **HNF1A** locus indexed by rs1169299 (**P_HET_**=4.8×10^-35^). This index SNV was negatively associated with the AFR-EAS axis (**P_HET_**=2.7×10^-11^), and positively associated with the AFR-EUR axis (**P_HET_**=4.6×10^-9^), corresponding to an AFR allelic effect (OR=1.02) that was intermediate between the EAS and EUR allelic effects (OR=0.95 and OR=1.05, respectively). In contrast, the association signal indexed by rs2237884, at the locus encompassing **INS**, **IGF2**, and **KCNQ1**, was not associated with either the AFR-EAS axis (**P_HET_**=0.61) or AFR-EUR axis (**P_HET_**=0.56), indicating no difference in allelic effects between AFR, EAS, and EUR GWAS (OR=1.03 for all three ancestry groups). Instead, the heterogeneity for this signal was driven by association with the third axis of genetic variation (**P_HET_**=2.8×10^-8^), which separates HIS and SAS GWAS (OR=1.01 and OR=0.97, respectively).

Ancestry-correlated heterogeneity can occur because of interaction between index SNVs and environmental/lifestyle factors that vary between ancestry groups, if not accounted for in the association analysis^17^. We therefore investigated whether ancestry-correlated heterogeneity could be explained by differences in the distribution of body mass index (BMI) across ancestry groups (Supplementary Figure 5). To do this, we extended the MR-MEGA meta-regression model to allow for allelic effect heterogeneity at index SNVs due to mean BMI in controls, in addition to variation by ancestry (**Methods**). Across all index SNVs, there was a negative correlation between T2D risk and BMI in controls (Supplementary Table 7), where the mean reduction in log-OR per unit of BMI was 0.0016 (**P**=9.6×10^-53^). After adjustment for BMI, the number of independent T2D associations with significant evidence for ancestry-correlated heterogeneity decreased from 127 (9.9%) to 49 (3.8%). These results indicate that ancestry-correlated heterogeneity at T2D index SNVs can be partly but not fully explained by differences in the distribution of BMI between ancestry groups.

## The contribution of under-represented populations to evidence for locus discovery

Despite the global burden of common complex diseases, including T2D, GWAS are biased towards EUR and EAS ancestry groups^18^. Failure to detect T2D association signals that contribute to disease risk outside of these ancestry groups limits opportunities for clinical translation of GWAS into under-represented ancestries and has the potential to exacerbate health disparities^19^. This multi-ancestry meta-analysis includes 98,060 T2D cases and 198,020 controls from AFR, HIS, and SAS GWAS (19.9% of the effective sample size), which substantially increases sample sizes in these under-represented groups over previous efforts^5, 6, 20^. To assess their contribution to the discovery of T2D association signals, we repeated the meta-regression using only EAS and EUR GWAS, including just one axis of genetic variation needed to separate these two ancestry groups (Supplementary Figure 1**, Methods**). Of the 1,289 association signals identified in the multi-ancestry meta-analysis, 589 did not retain genome-wide significance after exclusion of AFR, HIS, and SAS GWAS (Supplementary Figure 6, Supplementary Table 8), emphasizing the power of population diversity for locus discovery.

## Eight mechanistic clusters of T2D index SNVs with distinct cardiometabolic profiles

To understand the genetic contribution to phenotypic heterogeneity in T2D, we classified the 1,289 index SNVs according to their profile of associations (aligned to the T2D risk allele) with 37 cardiometabolic phenotypes from the largest available GWAS. These included glycaemic traits, anthropometric measures, body fat and adipose tissue volume, blood pressure, circulating plasma lipid levels, and biomarkers of liver function and lipid metabolism^21–29^ (Supplementary Table 9). We applied an unsupervised “hard clustering” approach with imputation of missing phenotype associations, which defines non-overlapping but exhaustive subsets of index SNVs with similar cardiometabolic profiles (**Methods**). According to the majority rule across 27 indices of cluster performance^30^, eight clusters of index SNVs was considered “optimal” (Figure 1, Table 1, Supplementary Figure 7**, Supplementary** Tables 10 and 11). Despite differences in the methods and phenotypes used for clustering compared to previous efforts^7, 8, 31^ (**Methods**), there is clear overlap in the cardiometabolic features and loci of five of the identified clusters with those previously reported, representing beta-cell dysfunction with positive or negative association with proinsulin (PI), and insulin resistance mediated via obesity, lipodystrophy, and liver/lipid metabolism. However, by increasing the number of index SNVs in the clustering by nearly four-fold over previous efforts, we provide a more granular view of the biological processes through which T2D associations impact disease, and highlight previously-unreported clusters of signals with cardiometabolic profiles that are representative of the metabolic syndrome, body fat, and residual glycaemic effects. We observed significant differences in allelic effects on T2D between the eight clusters (**P**<2.2×10^-16^), with weaker associations observed at index SNVs assigned to the three previously-unreported residual glycaemic, body fat, and metabolic syndrome clusters (Supplementary Figure 8, Supplementary Table 12).

**Figure 1.**
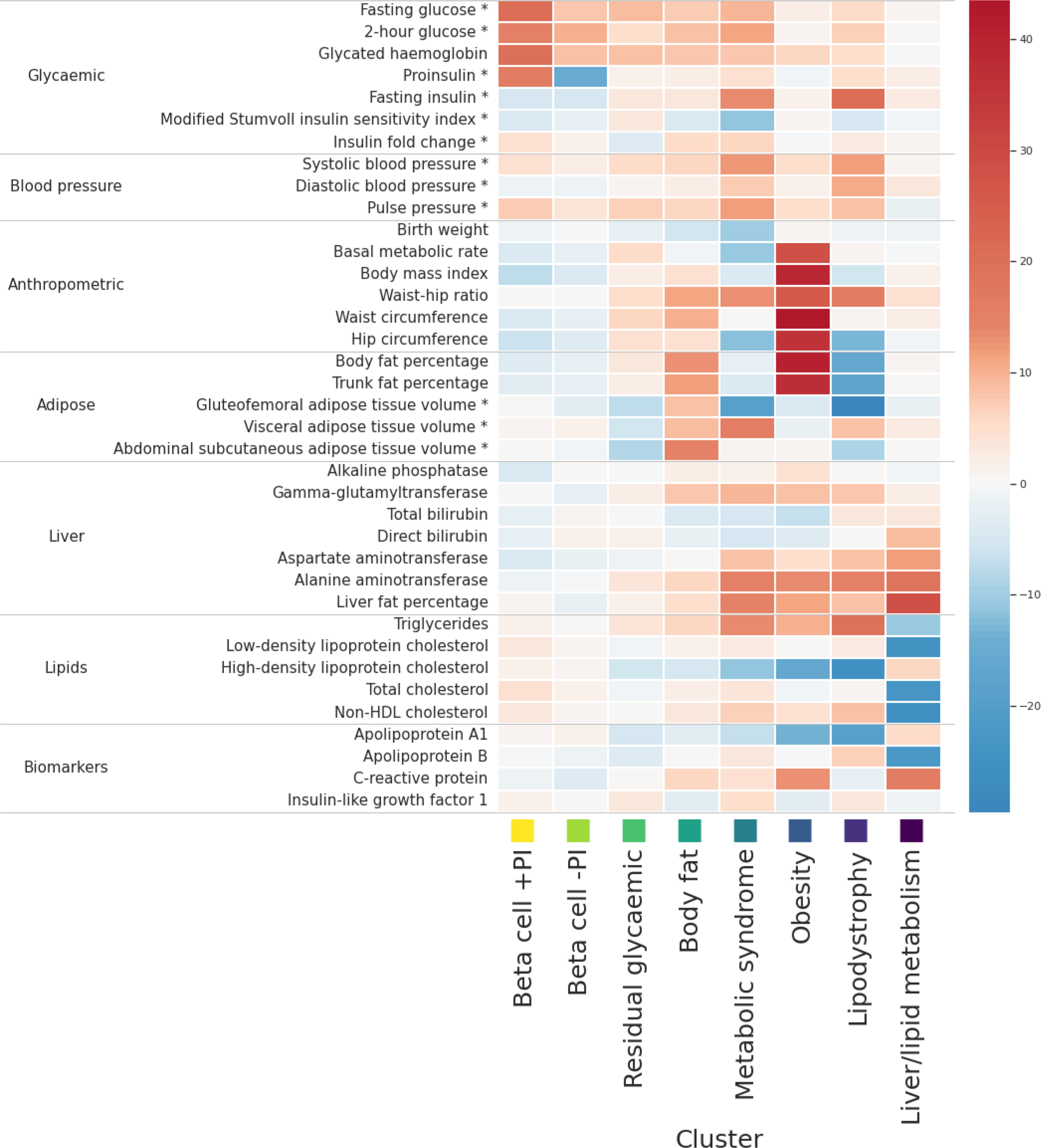
Heatmap of associations of 37 cardiometabolic phenotypes with eight mechanistic clusters of index SNVs for T2D association signals. Each column corresponds to a cluster. Each row corresponds to a cardiometabolic phenotype. The “temperature” of each cell represents the Z-score (aligned to the T2D risk allele) of association of the phenotype with index SNVs assigned to the cluster. *Phenotype is adjusted for body mass index.

T2D risk alleles at index SNVs in the two beta-cell dysfunction clusters are associated with increased fasting glucose (FG), two-hour glucose, and glycated haemoglobin (HbA1c), and with decreased fasting insulin (FI). Index SNVs in both clusters are also associated with PI, but with opposite directions of effect for the T2D risk allele. These findings are consistent with observed differences in directions of effect of PI-associated SNVs with FG^21^, which differentiate signals that reflect defects in the insulin synthesis pathway that occur upstream (decreased PI) or downstream (increased PI) of the first enzymatic step of the conversion of PI to insulin and C-peptide^32^. The beta-cell +PI cluster, where the T2D risk allele is associated with increased PI, comprises 91 index SNVs, which include T2D associations that map to loci encompassing genes with established roles in beta-cell function (such as **TCF7L2** and **CDKAL1**). The beta-cell -PI cluster, where the T2D risk allele is associated with decreased PI, comprises 89 index SNVs, which include T2D associations that map to monogenic diabetes genes that play a role in beta-cell development (such as **HNF1A**, **HNF4A**, **HNF1B**, **KCNJ11**, and **ABCC8**).

The clusters reflecting mechanisms of insulin resistance mediated via obesity, lipodystrophy, and liver/lipid metabolism, include index SNVs that are associated with anthropometric measures and circulating plasma lipid levels. T2D risk alleles at the 233 index SNVs in the obesity cluster are associated with increased BMI, waist-hip ratio (WHR), body fat percentage and basal metabolic rate, and with decreased high-density lipoprotein (HDL) cholesterol. The index SNVs assigned to this cluster include T2D associations that map to loci previously reported for BMI (such as **FTO** and **MC4R**). The lipodystrophy cluster comprises 45 index SNVs for which T2D risk alleles are associated with increased FI, WHR, blood pressure, and triglycerides, and with decreased body fat percentage, gluteofemoral adipose tissue (GFAT) volume, and HDL cholesterol. Index SNVs in this cluster include T2D associations that map to loci previously reported for regional adiposity (such as **IRS1**, **GRB14**, and **PPARG**). T2D risk alleles at the three index SNVs assigned to the liver/lipid metabolism cluster are associated with increased liver fat and liver-related biomarkers, and with decreased low-density lipoprotein cholesterol and total cholesterol. These index SNVs map to loci that have been previously implicated in non-alcoholic fatty liver disease (such as **TM6SF2** and **PNPLA3**).

The cardiometabolic profiles of the three remaining clusters are less well-defined, likely reflecting the weaker T2D associations of index SNVs assigned to them (Supplementary Figure 8). T2D risk alleles at the 166 index SNVs assigned to the metabolic syndrome cluster are associated with increased FG, WHR, triglycerides, and blood pressure, and with decreased HDL cholesterol, which together are used to define the syndrome. T2D risk alleles in this cluster are also associated with increased FI, and with accumulations of unhealthy fat depots: increased visceral adipose tissue (VAT) volume and liver fat, and with decreased GFAT volume. Observationally, individuals with the metabolic syndrome are at increased risk of T2D^33^, although Mendelian randomisation studies indicate that a causal effect is driven by increased waist circumference and increased FG^34^. T2D risk alleles at the 273 index SNVs assigned to the body fat cluster are associated with increased abdominal subcutaneous adipose tissue volume, VAT volume, and body fat percentage. Whilst the profile of associations with cardiometabolic phenotypes share these features in common with obesity-mediated insulin resistance, index SNVs in the body fat cluster are not strongly associated with BMI, lipid levels, or basal metabolic rate. Previous investigations have highlighted that body fat percentage is predictive of abnormal blood glucose in individuals with a healthy BMI^35^. Finally, T2D risk alleles at the 389 index SNVs assigned to the residual glycaemic cluster are most strongly associated with increased FG and HbA1c, but unlike the two beta-cell clusters, are not associated with PI or decreased FI.

## Clusters are differentially associated with insulin-related endophenotypes

The clustering provides a framework to better understand the diverse physiological processes through which T2D develops. We assessed the association of index SNVs with insulin-related endophenotypes that were not used for clustering and derived from hyperinsulinemic-euglycemic clamp assessments and oral glucose tolerance tests (OGTT)^36^, and homeostatic model assessment measures of beta-cell function (HOMA-B) and insulin resistance (HOMA-IR)^37^ (**Methods**). We observed significant heterogeneity in the effects of T2D risk alleles at index SNVs between clusters on HOMA-B (**P** HET <2.2×10^-16^), HOMA-IR (**P** =4.1×10^-15^), insulin secretion (OGTT-derived area under the curve for insulin normalised for glucose from baseline to 30 minutes, **P_HET_**=0.0026), and insulin sensitivity (clamp-derived glucose infusion rate, **P_HET_**=0.026). T2D risk alleles at index SNVs showed a gradient of effects on these correlated measures across clusters (Supplementary Figure 9**, Supplementary** Tables 13 and 14), representing a cline from insulin production and processing in the two beta-cell clusters (increased insulin sensitivity; decreased insulin secretion, HOMA-B, and HOMA-IR) through to insulin resistance (decreased insulin sensitivity; increased insulin secretion, HOMA-B, and HOMA-IR) that was most extreme in the lipodystrophy cluster.

## Clusters are differentially associated with insulin resistance-related disorders

T2D is genetically correlated with other insulin resistance-related disorders, including gestational diabetes mellitus (GDM) and polycystic ovary syndrome (PCOS). To understand the shared biological pathways driving these genetic correlations, we extracted association summary statistics for each T2D index SNV from the largest available published GWAS for both disorders^38, 39^ (**Methods**). We observed significant heterogeneity in the effects of T2D risk alleles at index SNVs between clusters for both disorders (Supplementary Figure 10, Supplementary Table 15): GDM (**P_HET_**=7.0×10^-16^) and PCOS (**P_HET_**=0.00022). Index SNVs in the beta-cell +PI cluster demonstrated the strongest associations with GDM. This cluster includes T2D index SNVs that overlap with association signals previously reported for GDM, mapping to/near **MTNR1B**, **CDKAL1**, **TCF7L2**, and **CDKN2A-CDKN2B**, consistent with hyperglycaemia due to beta-cell dysfunction on a background of pregnancy-induced physiologic insulin resistance^40^. In contrast, PCOS is most strongly associated with index SNVs in the obesity cluster, consistent with previous reports that the genetic correlation with T2D is primarily driven by higher BMI^41^.

## Clusters are differentially enriched for cell-type specific regions of open chromatin

To gain insight into tissue-specific regulatory processes underpinning mechanistic clusters, we integrated T2D association signals with Assay for Transposase-Accessible Chromatin using sequencing (ATAC-seq) peaks for 222 cell types derived from adult and foetal tissues via single-cell atlases of chromatin accessibility (CATLAS and DESCARTES)^42, 43^. This expands the range of cell types considered over previous efforts to understand cluster-specific enrichment for regulatory processes that were conducted only in bulk tissue or single-cell data in pancreatic islets^7, 8, 31^. Index SNVs assigned to six of the eight clusters were significantly enriched (**P**<0.00023, Bonferroni correction for 222 cell types) for regions of open chromatin that were specific to 20 cell types when compared to “null” SNVs that were not in LD with index SNVs, defined as **r**^2^<0.05, in any ancestry group (Figure 2, Supplementary Table 16**, Methods**).

**Figure 2.**
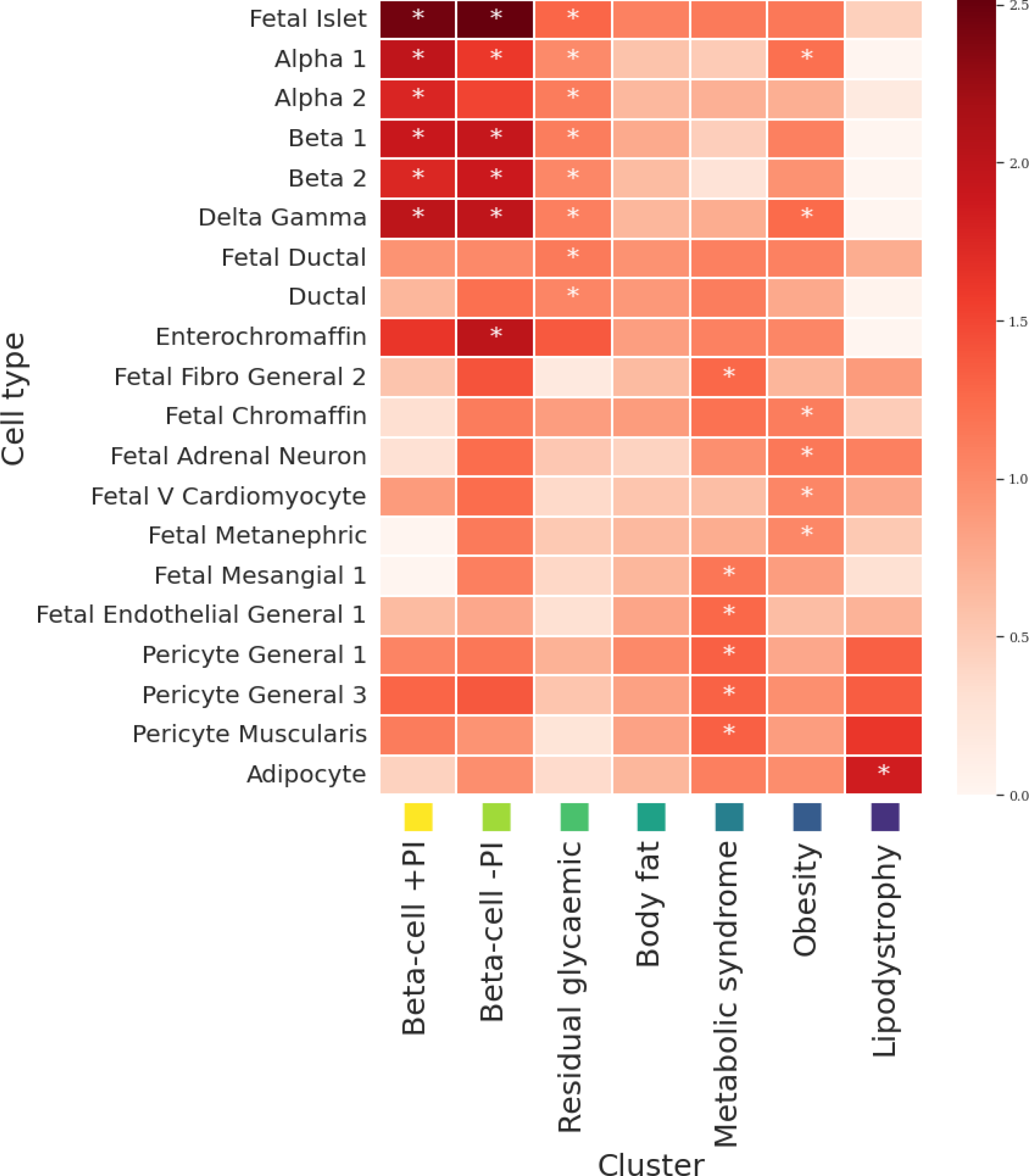
Heatmap of cluster-specific enrichments of T2D associations for cell type-specific regions of open chromatin derived from single-cell ATAC-seq peaks in adult and foetal tissue. Each column represents a mechanistic cluster. Each row represents a cell type that was significantly enriched (P<0.00023, Bonferroni correction for 222 cell types) for T2D associations in at least one cluster (indicated by an asterisk). The temperature of each cell defines the magnitude of the log-fold enrichment. The liver/lipid metabolism cluster is not presented because it includes only three T2D association signals and the regression model did not converge.

We observed significant enrichment for regions of open chromatin in foetal islets and adult neuroendocrine cells in pancreatic islets (alpha, beta, gamma, and delta) in the beta-cell +PI, beta-cell -PI, and residual glycaemic clusters. The beta-cell -PI cluster was also enriched in adult enterochromaffin cells, a type of enteroendocrine cell that plays an essential role in regulating intestinal motility and secretion in the gastrointestinal tract^44^. Enterochromaffin cells are a major target for GLP-1 and highly express GLP-1 receptor, whose agonists are widely used as medications for T2D^45^. The residual glycaemic cluster was also enriched in foetal and adult pancreatic ductal cells.

Some cell types important in the pathogenesis of obesity, such as the hypothalamus in the central nervous system, are not sufficiently covered by CATLAS/DESCARTES, and so were not available for this analysis. However, the obesity cluster was enriched for regions of open chromatin in adult pancreatic islets, although not as strongly as the beta-cell clusters. Enrichment was observed only for alpha, gamma, and delta cells, suggesting potential alternative pathways through which islets impact the development of T2D than through insulin secretion from beta cells. The obesity cluster was further enriched in foetal adrenal gland (chromaffin cells and adrenal neurons), foetal heart (ventricular cardiomyocytes), and foetal kidney (metanephric cells). Previous studies have reported enrichment of BMI loci/heritability for epigenomic annotations in pancreatic islets and adrenal gland^46, 47^, consistent with our findings.

The remaining four clusters (lipodystrophy, metabolic syndrome, body fat, and liver/lipid metabolism) were not significantly enriched for regions of open chromatin in pancreatic islets. The lipodystrophy cluster was enriched only in adult adipocytes, which confirms previous reports in bulk adipose tissue^8, 31^. Consistent with these results, association signals for WHR, triglycerides, and HDL cholesterol, which are strongly impacted by index SNVs in the lipodystrophy cluster, have been shown to be enriched in candidate cis-regulatory elements in adipocytes^43^. The metabolic syndrome cluster was enriched in cells that reside in the walls of blood vessels (adult pericytes and foetal endothelial cells), foetal kidney (mesangial cells), and foetal fibroblasts. Association signals for systolic and diastolic blood pressure, a key component of the metabolic syndrome, have been shown to be enriched in candidate cis-regulatory elements in these cell types^43^. Endothelial dysfunction is not only a consequence of insulin resistance, but impairs insulin signalling to further reduce insulin sensitivity, thereby providing a pathophysiological mechanism that links metabolic and cardiovascular components of metabolic syndrome^48^. We observed no significant enrichments in the body fat cluster or liver/lipid metabolism cluster.

## Ancestry-correlated heterogeneity differs between mechanistic clusters

We next sought to investigate whether the observed ancestry-correlated differences in allelic effects on T2D between ancestry groups varied across mechanistic clusters. To do this, we compared the magnitude and direction of association of index SNVs in each cluster with the first two axes of genetic variation that drive ancestry-correlated heterogeneity at T2D association signals (**Methods**). We observed significant differences in mean Z-scores for association between clusters for both the AFR-EAS axis (**P**=5.0×10^-6^) and the AFR-EUR axis (**P**=1.5×10^-6^). Index SNVs in the two beta-cell clusters were most positively associated with the AFR-EAS axis, indicating allelic effects on T2D that were greater in EAS than in AFR and EUR GWAS (Supplementary Figure 11, Supplementary Table 17). In contrast, index SNVs in the lipodystrophy and obesity clusters were most positively associated with the AFR-EUR axis, indicating allelic effects on T2D that were greater in EUR than in EAS or AFR GWAS. These results indicate that ancestry-correlated heterogeneity varies between mechanistic clusters, which may have clinical implications for T2D management across ancestry groups.

## Cluster-specific partitioned GRS are associated with vascular outcomes across ancestry groups

The major complications in individuals with T2D are macrovascular outcomes including coronary artery disease (CAD), ischemic stroke, and peripheral artery disease, and progression to severe microvascular complications, including end-stage diabetic nephropathy (ESDN) and proliferative diabetic retinopathy. We tested for association of a cluster-specific partitioned GRS with these clinical outcomes in up to 137,559 individuals (including 10,159 T2D cases) across four ancestry groups (AFR, EAS, EUR, and HIS) from the All of Us Research Program and Biobank Japan (**Methods**). These individuals were not included in the multi-ancestry meta-analysis, and so allowed us to avoid potential inflated type I error rates due to overlap between the discovery and testing datasets. To maximise sample size, we tested macrovascular outcomes in all individuals, adjusted for T2D status, and we tested microvascular complications only in individuals with T2D (**Methods,** Supplementary Table 18). We first conducted GRS association analyses within ancestry groups, and subsequently aggregated results across ancestry groups via fixed-effects meta-analysis (**Methods,** Supplementary Table 19). We also tested the association of an overall GRS, agnostic to cluster membership, to assess the evidence that mechanistic clusters in the partitioned GRS are differentially associated with T2D-related clinical outcomes. Figure 3 provides an overview of the associations of the overall GRS and each cluster-specific component of the partitioned GRS with the five T2D-related clinical outcomes across ancestry groups.

**Figure 3.**
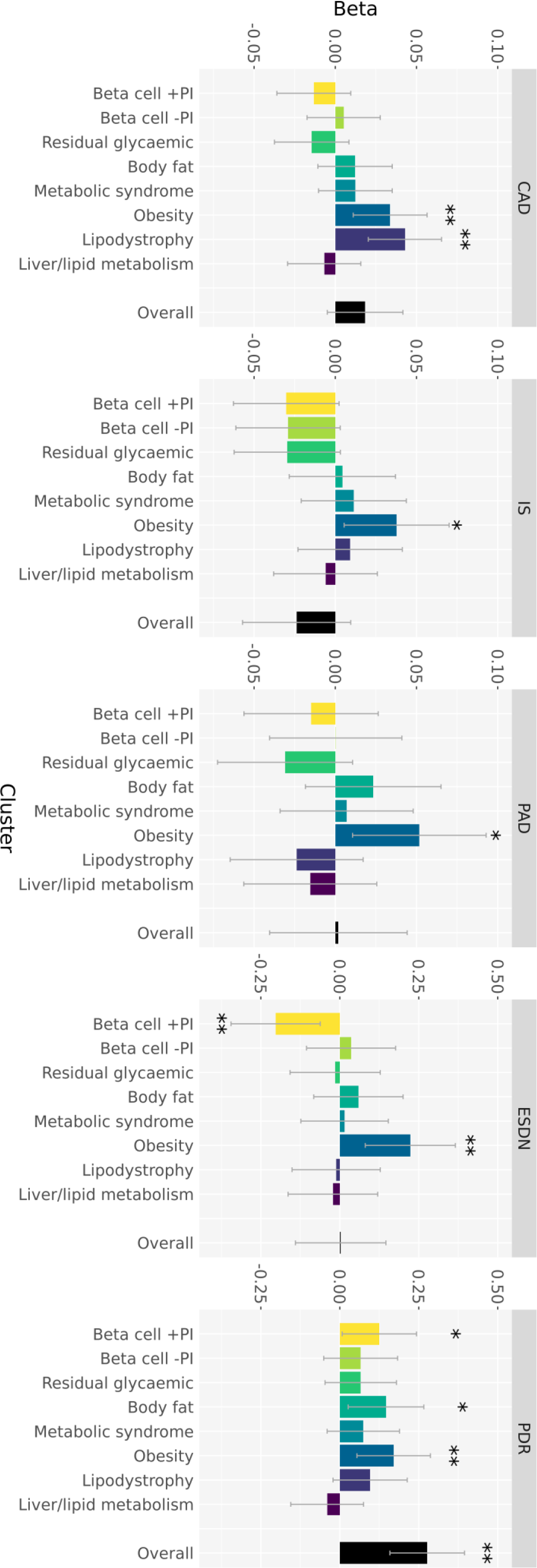
Associations of the overall GRS and cluster-specific partitioned GRS with five T2D-related vascular outcomes in up to 137,559 individuals from multiple ancestry groups. Each of the panels summarise the associations of the overall GRS and each cluster-specific component of the partitioned GRS with coronary artery disease (CAD), peripheral artery disease (PAD), ischemic stroke (IS), end-stage diabetic nephropathy (ESDN), and proliferative diabetic retinopathy (PDR). The height of each bar corresponds to the log-odds ratio (beta) per standard deviation of the GRS, and the grey bar shows the 95% confidence interval. Analyses of T2D-related macrovascular complications (CAD, PAD, and IS) were undertaken in all individuals, with adjustment for T2D status. Analysis of microvascular complications were undertaken in individuals with T2D only. *P<0.05, nominal association. **P<0.0063, Bonferroni correction for eight clusters.

We observed significant positive association (**P**<0.0063, Bonferroni correction for eight clusters) of two components of the partitioned GRS with CAD: the lipodystrophy cluster (OR=1.04 per standard deviation of GRS, **P**=0.00020) and the obesity cluster (OR=1.03, **P**=0.0036). There was no evidence of heterogeneity in the effects of these two clusters on CAD across ancestry groups (Supplementary Figure 12, Supplementary Table 19). Importantly, after adjustment for a CAD GRS derived from a recently-published multi-ancestry meta-analysis of CAD GWAS^49^, the positive CAD association with both components of the GRS remained nominally significant (Supplementary Figure 13, Supplementary Table 20): lipodystrophy cluster (OR=1.03, **P**=0.029) and obesity cluster (OR=1.04, **P**=0.011). These results demonstrate that additional information is provided by the partitioned GRS for association with CAD in individuals with T2D beyond a more general CAD GRS. CAD association signals have been reported to be enriched for annotations (from transcriptomics and epigenomics) in bulk tissues including aorta/arteries, heart, adrenal gland, and adipose^49–51^. This is in line with the observed enrichment of T2D index SNVs assigned to the obesity and lipodystrophy clusters in regions of open chromatin in foetal ventricular cardiomyocytes, foetal adrenal neuron and adult chromaffin cells in the adrenal gland, and adult adipocytes, providing a clear link to shared biological mechanisms driving development of both diseases. Whilst no components of the partitioned GRS were significantly associated with the other macrovascular outcomes, the obesity cluster had the strongest effect on both ischemic stroke (OR=1.04, **P**=0.022) and peripheral artery disease (OR=1.05, **P**=0.014). T2D risk alleles at index SNVs in the obesity cluster were strongly associated with increased C-Reactive Protein, indicating a potentially more pronounced role of inflammation in the development of ischemic stroke and peripheral artery disease than in CAD. We observed no significant evidence for association of the overall GRS with CAD (**P**=0.12), ischemic stroke (**P**=0.16), or peripheral artery disease (**P**=0.93). These results therefore highlight the advantages of the partitioned GRS over an overall GRS for detecting association with T2D-related macrovascular outcomes and providing insight into the biological processes that lead to these disease complications.

We observed significant positive association of the obesity cluster from the partitioned GRS with ESDN (OR=1.25, **P**=0.0022). We also observed a significant negative association of the beta-cell +PI cluster with ESDN (OR=0.82, **P**=0.0048). There was no evidence of heterogeneity in the effects of these two clusters on ESDN across ancestry groups, (Supplementary Figure 14, Supplementary Table 19), and the overall GRS was not associated with ESDN (**P**=0.97). T2D index SNVs assigned to the obesity cluster are enriched for regions of open chromatin in foetal metanephric cells, which is consistent with the observation that association signals for estimated glomerular filtration rate, a measure of renal function used to define chronic kidney disease, are enriched for kidney tissue-specific regulatory annotations^52^. The obesity cluster from the partitioned GRS was also significantly positively associated with proliferative diabetic retinopathy (OR=1.19, **P**=0.0041), with no evidence of heterogeneity in effects across ancestry groups (Supplementary Figure 15, Supplementary Table 19). However, in contrast to ESDN, the overall GRS was more significantly associated with proliferative diabetic nephropathy (OR=1.32, **P**=4.3×10^-6^) than any component of the partitioned GRS. Taken together, these results suggest that ESDN is disproportionately associated with obesity and confirm previous reports that proliferative diabetic retinopathy is driven by hyperglycaemia^53^ and therefore more strongly associated with the overall burden of T2D risk variants.

### DISCUSSION

T2D is a heterogeneous polygenic disease, which is typically diagnosed based solely on elevated glucose levels and treated without regard to the diverse pathophysiological processes that lead to the development and progression of hyperglycaemia. Instead, management strategies involve trial and error following treatment algorithms that are not informed by personalised pathways to disease. The complications of T2D have substantial morbidity and mortality and thus pose a huge public health burden. A more detailed understanding of the biological mechanisms through which the disease develops and progresses to T2D complications are therefore urgently needed and eagerly anticipated.

To better understand the aetiological heterogeneity of T2D across diverse populations, we assembled the largest collection of T2D GWAS for five ancestry groups through the Type 2 Diabetes Global Genomics Initiative. By increasing the effective sample size by almost three-fold compared to previous efforts, we identified a total of 611 loci attaining genome-wide association significance, 145 (23.7%) of which have not been previously reported. Of the 1,289 signals mapping to these loci, 589 (45.7%) would not have been identified without inclusion of GWAS from under-represented ancestry groups. We observed a highly significant enrichment of T2D association signals with evidence of ancestry-correlated heterogeneity. BMI could not fully explain this enrichment, indicating that interactions with other environmental/lifestyle factors that vary with ancestry are also likely to contribute to the observed differences in allelic effects between GWAS.

Within the landscape of the genetic architecture of T2D, we identified eight clusters of index SNVs with distinct profiles of associations with 37 cardiometabolic phenotypes, which defined pathophysiologic-relevant groupings. The addition of previously-unreported T2D signals identified through the multi-ancestry meta-analysis helped define three clusters that were not detected in previous clustering efforts^7, 8, 31^. These three clusters have cardiometabolic profiles that are consistent with T2D-related biology: residual glycaemic effects, accumulations of body fat, and the metabolic syndrome. Integration of cluster-specific T2D association signals with single-cell chromatin accessibility data across diverse cell types provided new insights into distinct tissue-specific regulatory processes and biological pathways through which these mechanistic clusters are likely to lead to disease. Finally, we leveraged the information captured by these mechanistic clusters to develop a partitioned GRS that was more strongly associated with T2D-related macrovascular outcomes and ESDN than an overall T2D GRS. Whilst the effect sizes of the cluster-specific components of the partitioned GRS were small, they enhance understanding of key biological processes driving heterogeneity in these clinical features of T2D phenotypes, and motivate future work to strengthen these effects through identification of additional, statistically-compelling T2D associations in larger sample sizes. Previous efforts have highlighted associations of cluster-specific components of a partitioned GRS with CAD and renal function in EUR populations^54^. For the first time across multiple ancestry groups, we demonstrated more significant associations of clinical outcomes with cluster-specific components of the partitioned GRS than with the overall GRS, suggesting that development of complications may be associated with genetic burden in particular biological pathways that is consistent between populations.

Our findings demonstrate the value of integrating multi-ancestry GWAS of T2D and cardiometabolic traits with single-cell epigenomics across diverse tissues to disentangle the aetiological heterogeneity driving the development and progression of T2D across population groups. Improved understanding of the varied pathophysiological processes that link T2D to vascular outcomes may offer a route to genetically-informed diabetes care and global opportunities for the clinical translation of T2D GWAS findings.

## CONSORTIA

**VA Million Veteran Program.** Kim M. Lorenz^10–12^, Kyung Min Lee^155, 156^, Julie A. Lynch^155, 156^, Philip S. Tsao^234, 286, 287^, Kyong-Mi Chang^10, 288^, Marijana Vujkovic^10, 288, 292^, and Benjamin F. Voight^10–12, 277^. Contributors to the VA Million Veteran Program are listed in the **Supplementary Note**.

**AMED GRIFIN Diabetes Initiative Japan.** Ken Suzuki^1–3^, Kyuto Sonehara^3, 16–18^, Shinichi Namba^3^, Momoko Horikoshi^128^, Shiro Maeda^128, 176, 177^, Koichi Matsuda^179^, Yukinori Okada^3, 16, 18, 188^, Nobuhiro Shojima^2^, Kenichi Yamamoto^3, 188, 225^, Toshimasa Yamauchi^2^, and Takashi Kadowaki^2, 283^. Contributors to AMED GRIFIN Diabetes Initiative Japan are listed in the **Supplementary Note**.

**Biobank Japan Project.** Koichi Matsuda^179^. Contributors to the Biobank Japan Project are listed in the **Supplementary Note**.

**Penn Medicine BioBank.** Lindsay A. Guare^124^, Daniel J. Rader^12, 276–278^, Marylyn D. Ritchie^12, 279, 280^, Marijana Vujkovic^10, 288, 292^, and Benjamin F. Voight^10–12, 277^. Contributors to the Penn Medicine BioBank are listed in the **Supplementary Note**.

**Regeneron Genetics Center.** Contributors to the Regeneron Genetics Center are listed in the **Supplementary Note**.

**eMERGE Consortium.** Yoonjung Yoonie Joo^62–64^, Abel N. Kho^148, 149^, and M. Geoffrey Hayes^63, 246, 247^. Contributors to the eMERGE Consortium are listed in the **Supplementary Note**.

International Consortium of Blood Pressure (ICBP). Zoha Kamali^78, 79^. Contributors to ICBP are listed in the Supplementary Note.

**Meta-Analyses of Glucose and Insulin-Related Traits Consortium (MAGIC).** K Alaine Broadaway^66^, Alice Williamson^34, 77^, Karen L. Mohlke^66^, and Claudia Langenberg^34, 272, 273^. Contributors to MAGIC are listed in the **Supplementary Note.**

## Data Availability

Genome-wide association summary statistics from the multi-ancestry meta-analysis reported in this study will be made available through the DIAGRAM Consortium website (http://www.diagram-consortium.org/downloads.html) an acceptance of the manuscript.

## ACKNOWLEDGEMENTS

Central analyses were supported by Japan Agency for Medical Research and Development (JP21km0405213, JP20km0405202, and JP21tm0424218), NHGRI (HG011723), American Diabetes Association Innovative and Clinical Translational Award (1-19-ICTS-068), American Heart Association Postdoctoral Fellowship (15POST24470131 and 17POST33650016), American Diabetes Association (11-22-JDFPM-06), Corporal Michael J Crescenz VA Medical Center Research, NIDDK (DK126194 and DK105535), Versus Arthritis (21754), NIHR Manchester Biomedical Research Centre (NIHR203308). A complete list of acknowledgments and funding appears in the **Supplementary Note**.

We thank the International Consortium of Blood Pressure (ICBP) and the Meta-Analyses of Glucose and Insulin-related traits Consortium (MAGIC) for providing pre-publication access to GWAS summary statistics for blood pressure, proinsulin, and post-challenge insulin resistance measures.

The views expressed in this article are those of the authors and do not necessarily represent those of: the UK National Health Service, the UK National Institute for Health Research, or the UK Department of Health and Social Care; the US National Heart, Lung, and Blood Institute, the US National Institute of Neurological Disorders and Stroke, the US National Institute on Aging, the US National Institutes of Health, the US Department of Health and Human Services, the US Department of Veterans Affairs, the US Food and Drug Administration, or the US Government.

## AUTHOR CONTRIBUTIONS

**Central analysis group:** K.Suzuki, K.H., L.S., H.J.T., X.Y., K.M.Lorenz, R.Mandla, A.H.-C., N.W.R., O.B., A.L.d.S.V.A., A.Mahajan, C.N.S., J.M.M., M.B., J.I.R., M.V., B.F.V., A.P.M., E.Z. **Study-level primary analysis:** K.Suzuki, H.J.T., X.Y., K.M.Lorenz, R.Mandla, A.H.-C., K.Sonehara, S.N., S.S.K.L., M.H.P., L.E.P., P.S., B.V., M.K., F.B., K.Lin, X.G., W.Zhang, J.Y., Y.J.K., M.Graff, F.T., J.N., A.Lamri, M.N., S.Moon, R.A.S., J.P.C., J.-J.L., I.P., D.T., E.J.P., J.-F.C., L.F.B., Y.T., Y.H., G.T., N.G., T.S., M.W., C.Sarnowski, C.G., D.N., S.T., S.-H.K., J.Long, M.Sun, L.T., W.-M.C., S.S.N., R.N., V.J.Y.L., C.H.T.T., Y.Y.J., C.-H.C., L.M.R., B.P.P., A.N., L.R.Y., G.C., J.A.B., E.K., P.A., A.H.X., H.S.C., B.E.C., J.Tan, M.C.Y.N., X.S., A.Mahajan, C.N.S., A.P.M. **Study-level sample collection, phenotyping, genotyping, and additional analysis:** L.S.A., A.A., C.A.A.-S., T.S.A., S.S.A., A.Bertoni, J.B.-J., I.B., T.A.B., C.F.B., A.S.B., M.Canouil, J.C.N.C., L.-C.C., M.-L.C., J.Chen, S.-H.C., Y.-T.C., Z.C., L.-M.C., M.Cushman, J.Danesh, S.K.D., H.J.d.S., G.D., L.D., A.P.D., S.D., Q.D., K.-U.E., L.S.E., D.S.E., M.K.E., K.F., J.S.F., I.F., O.H.F., T.M.F., B.I.F., P.G., H.C.G., V.G., C.G.-V., M.E.G.-V., P.G.-L., M.Gross, L.A.G., S.Hackinger, S.Han, A.T.H., C.H., M.Horikoshi, A.-G.H., W.Hsueh, M.Huang, W.Huang, Y.-J.H., M.Y.H., C.-M.H., S.I., M.A.I., M.Ingelsson, M.T.I., M.Isono, H.-M.J., F.J., G.J., J.B.J., T.J., F.R.K., A.Kasturiratne, T.Katsuya, V.K., T.Kawaguchi, J.M.K., A.N.K., C.-C.K., M.G.K., D.-H.K., F.K., J.K., K.Läll, L.A.L., K.M.Lee, M.-S.L., N.R.L., A.Leong, L.Li, Y.Liu, R.L.-G., S.Lithgart, C.M.L., A.Linneberg, C.-T.L., J.Liu, A.E.L., T.L., J.Luan, A.O.L., X.L., J.Lv, J.A.L., V.L., S.Maeda, V.M., S.R.M., K.M., T.M., A.Metspalu, H.M., A.D.M., J.L.N., M.A.N., U.N., I.N., Y.O., L.O., S.R.P., S.P., P.P., M.A.Pereira, A.P., F.J.P., H.G.P., B.P., G.Prasad, L.J.R.-T., A.P.R., M.R., R.R., K.R., C.Sabanayagam, K.Sandow, A.Sankareswaran, N.Sattar, S.S., M.Shahriar, B.S., J.S., D.M.S., N.Shojima, J.A.S., W.Y.S., A.Stančáková, V.S., A.M.S., K.Strauch, K.D.T., B.Thorand, U.T., B.Tomlinson, T.C.T., F.-J.T., J.Tuomilehto, T.T.-L., M.S.U., A.V., R.M.v.D., J.B.v.K., R.V., N.W.-R., E.W., A.R.W., K.W.v.D., D.R.W., C.S.Y., Ken.Yamamoto, Kenichi.Yamamoto, K.Yoon, C.Y., J.-M.Y., S.Y., M.Z., L.Z., W.Zheng, J.S.P., T.-Y.W., F.S.C., M.O.G., K.L.M., J.C.F., J.E.B., M.I.M., M.V. **Study-level principal investigator:** L.J.R., M.Igase, E.Ipp, S.R., Y.S.C., L.Lind, M.A.Province, M.F., C.L.H., E.Ingelsson, A.B.Z., B.M.P., Y.-X.W., C.N.R., D.M.B., F.M., Y.Liu, M.Y., S.L.R.K., P.A.P., J.S.P., J.C.E., A.Bonnefond, P.F., J.G.W., W.H.H.S., J.-Y.W., M.G.H., R.C.W.M., T.-Y.W., D.O.M.-K., T.T., G.R.C., F.S.C., D.B., G.Paré, M.M.S., H.A., A.A.M., X.-O.S., K.-S.P., J.W.J., M.Cruz, Y.-D.I.C., S.S.R., R.M.-C., H.G., C.-Y.C., M.G., E.-S.T., J.Dupuis, N.K., M.L., A.Köttgen, W.-P.K., D.W.B., C.N.A.P., J.S.K., C.K., S.Liu, K.E.N., D.S., T.H., O.P., N.J.W., J.Lee, B.-J.K., I.Y.M., R.G.W., K.Stefansson, K.L.M., C.L., C.A.H., R.J.F.L., J.C.F., D.J.R., M.D.R., S.Z., R.Mägi, J.C.D., T.Y., T.Kadowaki, J.C.C., J.E.B., P.S.T., K.-M.C., M.I.M., J.B.M., J.M.M., M.B., J.I.R., M.V., B.F.V., A.P.M., E.Z. **MAGIC analysis:** K.A.B., A.W., K.L.M., C.L. **ICBP analysis:** Z.K. **GUARDIAN Consortium analysis:** J.Cui, M.O.G. **DIAMANTE Consortium ancestry-specific data co-ordination:** J.C.C., M.C.Y.N., X.S., J.E.B., A.Mahajan. **Manuscript preparation:** K.Suzuki, K.H., L.S., H.J.T., X.Y., K.M.Lorenz, R.Mandla, M.I.M., J.B.M., A.Mahajan, C.N.S., J.M.M., M.B., J.I.R., M.V., B.F.V., A.P.M., E.Z. **T2DGGI steering group:** M.I.M., J.B.M., A.Mahajan, C.N.S., J.M.M., M.B., J.I.R., M.V., B.F.V., A.P.M., E.Z.

## FIGURE LEGENDS

## METHODS

### Ethics statement

Study-level ethics statements are provided in the Supplementary Note.

#### Study-level analyses

Within each study, we assigned individuals to one of five ancestry groups using self-report and genetic background (Supplementary Tables 1 and 2). Any individuals that could not be assigned to a single ancestry group were excluded as population outliers. Within each ancestry group-specific GWAS, we conducted quality control of genotype data. Subsequently, we pre-phased individuals and imputed up to reference panels from the Trans-Omics for Precision Medicine Program^10^, Haplotype Reference Consortium^11^, 1000 Genomes Project (phase 1, March 2012 release; phase 3, October 2014 release)^12, 13^, or population-specific whole-genome sequencing^55–61^ (Supplementary Table 3). Studies imputed to reference panels mapped to GRCh38/hg38 were lifted back to hg19 using the UCSC liftOver tool (https://genome.ucsc.edu/cgi-bin/hgLiftOver). We excluded SNVs with poor imputation quality and/or minor allele count (MAC) <5 from downstream association analyses (Supplementary Table 3).

Within each ancestry group-specific GWAS, we tested for association of each SNV with T2D via generalised linear regression, under an additive model in the dosage of the minor allele, with adjustment for age and sex (where appropriate), and additional study-specific covariates (Supplementary Table 3). We employed different strategies to account for population stratification and/or kinship: (i) exclude closely related individuals and adjust for principal components derived from a genetic relatedness matrix (GRM) as additional covariates in the regression model; or (ii) incorporate a random effect for the GRM in a mixed model (Supplementary Table 3). Allelic effects and corresponding standard errors that were estimated from a linear mixed model were converted to the log-odds scale^62^. We corrected study-level association summary statistics for residual structure by the LD-score regression intercept^63^ (Supplementary Table 3), which was calculated using an LD reference that we derived from ancestry-matched haplotypes from the 1000 Genomes Project (phase 3, October 2014 release)^13^.

#### Multi-ancestry meta-analyses

We analysed autosomal bi-allelic SNVs that overlap reference panels from the 1000 Genomes Project reference panel (phase 3, October 2014 release)^13^ and the Haplotype Reference Consortium^11^. We considered only those SNVs with MAF > 0.5% in haplotypes in at least one of the five major ancestry groups in the 1000 Genomes Project (phase 3, October 2014 release)^13^. We excluded SNVs that differed in allele frequency by >20% when comparing reference panels in the same subsets of haplotypes.

We used meta-regression, implemented in MR-MEGA^14^, to aggregate association summary statistics across GWAS. MR-MEGA models allelic effect heterogeneity that is correlated with genetic ancestry by including axes of genetic variation as covariates in the meta-regression model to capture population diversity between GWAS. We used SNVs that were reported in all studies to construct a distance matrix of mean effect allele frequency differences between each pair of GWAS. We implemented multi-dimensional scaling of the distance matrix to obtain three principal components that represent axes of genetic variation to separate GWAS from the five major ancestry groups (Supplementary Figure 1).

For each SNV, we aggregated inverse-variance weighted allelic effects across GWAS via linear regression, including three axes of genetic variation as covariates. We tested for: (i) association with T2D allowing for ancestry-correlated allelic effect heterogeneity between GWAS; (ii) ancestry-correlated allelic effect heterogeneity between GWAS (defined by the axes of genetic variation); and (iii) residual allelic effect heterogeneity between GWAS (due to differences in environmental exposures and/or study design). We also aggregated association summary statistics across GWAS via fixed-effects meta-analysis (inverse-variance weighting of allelic effects) using METAL^64^. To assess the extent of residual structure between GWAS, we calculated the genomic control inflation factor^65^ for the multi-ancestry meta-regression and fixed-effects meta-analysis.

#### Defining T2D signals and loci

We began by identifying all SNVs attaining the traditional genome-wide significance threshold (**P**<5×10^-8^) for association with T2D from the multi-ancestry meta-regression. Clumps were formed around index variants, which were selected using a greedy algorithm in PLINKv1.9^66^, after ranking SNVs by ascending **P**-value. SNVs <5Mb from an index SNV were assigned to the clump if **r**^2^>0.05 in at least one of the five ancestry groups using haplotypes from the 1000 Genomes Project (phase 3, October 2014 release)^13^. Index SNVs separated by <1Mb were assigned to the same locus. Each locus was then defined as mapping 500kb up- and down-stream of index SNVs contained within it. We considered the locus to have been previously reported if it contained variants discovered in published large-scale T2D GWAS at genome-wide significance.

#### Ancestry group-specific meta-analyses

We aggregated association summary statistics across GWAS from the same ancestry group via fixed-effects meta-analysis based on inverse-variance weighting of allelic log-OR to obtain effect size estimates using METAL^64^. We estimated the mean effect allele frequency across GWAS from each ancestry group, weighted by the effective sample size of the study. We generated forest plots of association summary statistics of index SNVs across ancestry groups using the R package meta (https://cran.r-project.org/package=meta/).

Contribution of each of axes of genetic variation to ancestry-correlated heterogeneity. For each index SNV, we calculated a Z-score (beta/SE) for association with each axis of variation by aligning the effect from the meta-regression model to the T2D-risk allele. For each index SNV, we identified the axis of genetic variation with the strongest association (greatest magnitude Z-score).

#### Impact of BMI on ancestry-correlated heterogeneity in allelic effects

For each index SNV, we aggregated inverse-variance weighted allelic effects across GWAS via linear regression, implemented in MR-MEGA^14^, including as covariates: (i) three axes of genetic variation; and (ii) mean BMI in controls. We tested for: (i) ancestry-correlated allelic effect heterogeneity between GWAS, after adjustment for BMI; and (ii) allelic effect heterogeneity due to BMI, after adjustment for ancestry. For the 1th index SNV, we estimated the meta-regression coefficient and corresponding variance, per unit of BMI, denoted by y_}_ and v_}_, respectively. Across all index SNVs, the mean effect on log-OR per unit of BMI was given by ∑_}_ y_}_⁄v_}_, with corresponding variance ∑ 1⁄v ]^-1^.

#### European and East Asian ancestry group meta-regression

For each SNV, we aggregated inverse-variance weighted allelic effects across EAS and EUR GWAS via linear regression, including only the first axis of genetic variation as a covariate, since this was sufficient to separate the two ancestry groups (Supplementary Figure 1). We tested for association with T2D allowing for ancestry-correlated allelic effect heterogeneity between GWAS under this model.

#### Defining clusters of T2Dindex SNVs with distinct cardiometabolic profiles

We considered cardiometabolic-related quantitative phenotypes that are used to define T2D status and/or are associated with risk of T2D or complications. We excluded phenotypes for which GWAS summary statistics were available only after imputation to reference panels from the International HapMap Project^67^ because they did not provide sufficient coverage of SNVs included in the multi-ancestry meta-analysis. We considered the largest available GWAS meta-analysis (ancestry-specific or multi-ancestry) that provided the following association summary statistics for each SNV: effect allele, other allele, effect estimate aligned to the effect allele, and standard error of the effect estimate. A summary of phenotypes, GWAS resources, and references are available in Supplementary Table 9. We re-aligned the effect estimate to the T2D risk allele from the fixed-effects multi-ancestry meta-analysis, denoted {_i}_ for the 1th index SNV and the ith phenotype. We then calculated a sample size corrected Z-score, given by zi} = {i}⁄(JNi si}), where si} is the standard error of the effect estimate of the 1th index SNV and the ith phenotype, and N_i_ is the maximum sample size reported for the ith phenotype. Standardisation by sample size enabled a more uniform weighting of cardiometabolic phenotypes in the subsequent clustering of index SNVs. Where association summary statistics were not reported for the 1th index SNV and the ith phenotype, the Z-score was set as “missing”.

Previous efforts to construct partitioned GRS for T2D have implemented “soft clustering” approaches, such as Bayesian non-negative matrix factorisation, that generate weights for cluster membership for each index SNV^8^. Assignment of index SNVs to clusters is then determined given a threshold weight for cluster membership, allowing for the possibility that a T2D association signal impacts on disease through multiple pathophysiological pathways. In practice, however, the choice of threshold for cluster membership is subjective, and some index SNVs remain unassigned. Bayesian non-negative matrix factorisation also considers positive and negative associations with the same phenotype as independent variables in the clustering, and cannot allow for missing phenotype associations, relying instead on the use of proxy SNVs, which can be challenging to identify in a multi-ancestry context.

To address these potential limitations, we conducted K-means clustering of index SNVs with imputation of missing Z-scores using the R package ClustImpute (https://cran.r-project.org/package=ClustImpute). For a pre-defined number of clusters, ClustImpute replaces missing Z-scores at random from the marginal distribution for the phenotype in the first iteration and performs K-means clustering. In subsequent iterations, missing Z-scores are updated, conditional on the current cluster assignment, so that correlations between phenotypes are considered. At each iteration, penalizing weights are imposed on imputed values and successively decreased (to zero) as the missing data imputation improves. In this “hard clustering” approach, each index SNV is assigned to exactly one cluster. Finally, we determined the “optimal” number of clusters according to the majority rule across 27 indices of cluster performance^30^, implemented in the R package NbClust (https://cran.r-project.org/package=NbClust).

We tested for association of the ith phenotype with index SNVs across clusters in a linear regression model, given by E(z_i}_) = ∑_k_ y_ik_c_}k_, where c_}k_ is an indicator variable that takes the value “1” if the 1th index SNV was assigned to the kth cluster and “0” otherwise. The strength/direction of the association of each phenotype with each cluster was then presented in a heatmap, where the “temperature” was defined by the direction of the regression coefficient y_ik_ and the corresponding -log_10_ P-value. Regression models were fitted using the glm function in R.

#### Cluster-specific associations of index SNVs with T2D

We tested for association of T2D with index SNVs across clusters in a linear regression model, given by E({_}_) = ∑_k_ y_k_c_}k_, where c_}k_ is an indicator variable that takes the value “1” if the 1th index SNV was assigned to the kth cluster and “0” otherwise, and weighted by the inverse of the variance of the allelic effect. We tested for heterogeneity in cluster effects on each endophenotype by comparing the deviance of this model with that of E({_}_) = y_0_, again weighted by the inverse of the variance of the allelic effect. Regression models were fitted using the glm function in R.

#### Cluster-specific associations of index SNVs with insulin-related endophenotypes and insulin resistance-related disorders

We extracted association summary statistics for measures of glucose homeostasis derived from hyperinsulinemic-euglycemic clamp assessments and oral glucose tolerance tests (OGTT) performed by the GUARDIAN Consortium^36^, which were obtained from GWAS undertaken in up to 1,316 Mexican American participants without diabetes from the Mexican American Coronary Artery Disease (MACAD) study^68^ and the Hypertension and Insulin Resistance (HTN-IR) study^69^. The measures used were: insulin sensitivity (clamp-derived glucose infusion rate in 1,316 participants from MACAD and HTN-IR); insulin clearance (clamp-derived metabolic clearance rate of insulin in 1,261 participants from MACAD and HTN-IR); and insulin secretion (OGTT-derived area under the curve for insulin normalised for glucose from baseline to 30 minutes in 513 participants from MACAD). We also extracted association summary statistics for homeostatic model assessment measures of beta-cell function (HOMA-B) and insulin resistance (HOMA-IR) from published GWAS meta-analyses of up to 36,466 non-diabetic individuals of European ancestry from MAGIC^37^. Note that we did not include HOMA measures as part of the cardiometabolic phenotype clustering because they were imputed only up to reference panels from the International HapMap Consortium^67^, and thus included a high proportion of missing Z-scores across index SNVs that we anticipated would introduce too much noise with ClustImpute. We also extracted association summary statistics for insulin resistance-related disorders from published GWAS meta-analyses of: (i) 5,485 GDM cases and 347,856 female controls of diverse ancestry from the GenDIP Consortium^39^; and (ii) 10,074 PCOS cases and 103,164 female controls of European ancestry^38^.

For each endophenotype/disorder, we aligned the effect estimate to the T2D risk allele from the fixed-effects multi-ancestry meta-analysis, denoted {_}_ for the 1th index SNV. We then calculated the Z-score, given by z_}_ = {_}_⁄s_}_ , where s_}_ is the standard error of the effect estimate of the 1th index SNV. We tested for association of each endophenotype with index SNVs across clusters in a linear regression model, given by E(z_}_) = ∑_k_ y_k_c_}k_, where c_}k_ is an indicator variable that takes the value “1” if the 1th index SNV was assigned to the kth cluster and “0” otherwise. We tested for heterogeneity in cluster effects on each endophenotype by comparing the deviance of this model with that of E(z_}_) = y_0_. Regression models were fitted using the glm function in R.

#### Enrichment of T2D associations for cell type-specific regions of open chromatin within clusters

For each T2D association signal, we defined “null” SNVs that mapped within 50kb of the index SNV and were not in LD (**r**^2^>0.05) with the index SNV in any of the five major ancestry groups using haplotypes from the 1000 Genomes Project (phase 3, October 2014 release)^13^. We defined an indicator variable, Y , taking the value “1” if the 1th SNV is an index SNV and “0” if the 1th SNV is a null SNV. We mapped index SNVs and null SNVs to genic regions, as defined by the Ensembl Project (release 104)^70^, including protein-coding exons, and 3’ UTRs and 5’ UTRs. We defined indicator variables, c^EXON^, c^3UTR^, and c^SUTR^, that each take the value “1” if the 1th SNV mapped to the respective genic annotation and “0” otherwise. We also mapped index SNVs and null SNVs to ATAC-seq peaks for 222 cell types derived from adult and foetal tissues via single-cell atlases of chromatin accessibility (CATLAS and DESCARTES)^42, 43^. We defined an indicator variable, x , that takes the value “1” if the 1th SNV mapped to an ATAC-seq peak for the ith cell type and “0” otherwise. Within each cluster, we modelled enrichment of T2D associations for ATAC-seq peaks in the ith cell type, after accounting for genic annotations, in a Firth bias-reduced logistic regression, given by

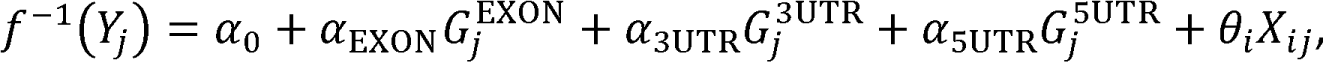

where J is the logit link function. In this expression, a_0_ is an intercept, a_EXON_ , a_3UTR_, and a_SUTR_ are log-fold enrichments of genic annotations, and e_i_ is the log-fold enrichment of ATAC-seq peaks in the ith cell type. We conducted a test of enrichment of the ith cell type by comparing the deviances of models in which e_i_ = 0 and e_i_ is unconstrained. We identified cell types with significant evidence of enrichment (**P**<0.00023, Bonferroni correction for 222 cell types). All models were fitted using the R package logistf (https://cran.r-project.org/package=logistf).

#### Difference in ancestry-correlated heterogeneity between mechanistic clusters

We tested for differences in Z-scores (beta/SE) for association of index SNVs in each cluster with the ith axis of genetic variation by comparing two linear models via ANOVA: (i) J^-1^(z_i}_) = ,_0i_ ; and (ii) J^-1^(z_i}_) = ∑_k_ ,_ki_ c_}k_. In these expressions: J is the identity link function; z_i}_ is Z-score for the 1th index SNV; c_}k_ is an indicator variable that takes the value “1” if the 1th index SNV was assigned to the kth cluster and “0” otherwise; and ,_0i_ and ,_ki_ are regression coefficients. Regression models were fitted using the glm function in R.

#### Cluster-specific partitioned GRS analyses

We tested for association of a cluster-specific partitioned GRS and an overall GRS with T2D-related macrovascular outcomes (CAD, ischemic stroke, and peripheral artery disease) and microvascular complications (ESDN and proliferative diabetic retinopathy) in participants from the All of Us Research Program (AoURP, multiple ancestry groups) and Biobank Japan (BBJ, East Asian ancestry).

##### AoURP cohort description, sequencing, quality control, and phenotype derivation

We considered 78,260 participants with whole-genome sequencing (WGS) and electronic health record (EHR) data from the AoURP Controlled Tier Dataset v5^71, 72^. Details of the generation and quality control of the genomic data can be found in the AoURP Genomic Quality Report release C2021Q3R5 (https://support.researchallofus.org/hc/en-us/article_attachments/7002034709652/All_Of_Us_Beta_Release_Genomic_Quality_Report__4_.pdf). We used genetic ancestries defined in the AoURP Genomic Quality Report release C2021Q3R5. We removed related individuals in the maximal independent set (kinship score >0.1) as described in the AoURP Genomic Quality Report release C2021Q3R5. To reduce the computational burden of the WGS dataset, we considered only bi-allelic SNVs with MAF ≥0.1% across all participants. To correct for population structure, within each ancestry, we derived principal components using Hail v0.2.107 (https://hail.is/docs/0.2/index.html). In these calculations, we excluded SNVs that were not present in the 1000 Genomes Project (phase 3, October 2014 release) reference panel^13^. We also excluded SNVs with MAF <1% and deviation from Hardy-Weinberg equilibrium (**P**<10^-6^), and subsequently extracted autosomal LD-pruned SNVs (**r**^2^<0.05). Cases of T2D, T2D-related macrovascular outcomes, and microvascular complications were derived from the combination of diagnosis codes (ICD-9-CM and ICD-10-CM), drug exposures, and LOINC codes for laboratory test results, extracted from EHR data. Full details are provided in the **Supplementary Methods**.

##### BBJ cohort description, genotyping, quality control, and phenotype derivation

BBJ is a multi-institutional hospital-based registry that comprises DNA and medical records from individuals of Japanese ancestry^73, 74^. The first BBJ cohort comprises approximately 200,000 participants with at least one of 47 common diseases collected between 2003 and 2007. The second BBJ cohort comprises approximately 67,000 participants with at least one of 38 common diseases collected between 2013 and 2017. Physicians of 66 cooperating hospitals determined the eligibility of cases. Only those individuals who were not included in the multi-ancestry meta-analysis were considered for testing of the partitioned GRS.

Genomic DNA was prepared following standard protocols from peripheral blood samples and genotyped using the Illumina Asian Screening Array, following the manufacturer’s instructions. We excluded individuals with call rate <98% and outliers from the cluster of East Asian populations based on principal component analysis with reference individuals from Phase II HapMap^67^. We excluded SNVs with call rate <99%, MAC <5, exact Hardy-Weinberg equilibrium **P**<10^-10^, and >5% difference in MAF when compared with Japanese whole-genome sequence data^58, 75^ and the Tohoku Medical Megabank Project^76^. After quality control, we performed pre-phasing using SHAPEIT4^77^. Phased haplotypes were imputed to the combined reference panel of 1000 Genomes Project Phase 3 and Japanese whole-genome sequencing data from 1,037 individuals^58, 75^ using Minimac4^78^. We subsequently excluded individuals with a mismatch between inferred genetic sex and sex registered in clinical information, who were not in a set of unrelated individuals defined by using PLINK with king-cutoff <0.09375, or were outliers of heterozygosity rates (more than 5 SD from the mean). To correct for population structure, we derived principal components using PLINKv2.0^79^, calculated from a set of autosomal LD-pruned SNVs (**r**^2^<0.1) with MAF ≥0.5% after excluding the major histocompatibility complex region.

We selected participants of at least 18 years of age for GRS analyses. We defined T2D cases as participants with a diagnosis of T2D, made by physicians at participating hospitals, but not type 1 diabetes, mitochondrial diabetes, maturity-onset diabetes of the young, or any other type of diabetes^80^. We extracted cases of microvascular complications from medical records in which diagnosis was made by physicians at participating hospitals. We defined controls for microvascular complications as T2D cases without any diagnosis of diabetic nephropathy or diabetic retinopathy. We defined CAD as a composite of stable angina, unstable angina, and myocardial infarction. These conditions, in addition to ischemic stroke and peripheral artery disease, were diagnosed by physicians at collaborating hospitals based on general medical practices following relevant guidelines.

##### GRS association analyses in AoURP and BBJ

We conducted analyses separately for each ancestry group in AoURP (African, American, and European) and BBJ (East Asian). For each ancestry, we performed GRS analyses for T2D-related macrovascular outcomes using all individuals, irrespective of T2D status, and for microvascular complications in individuals with T2D only. For each analysis, we calculated overall and cluster-specific GRS for each individual, with each index SNV weighted by the allelic log-OR from the ancestry-specific meta-analyses. We did not include index SNVs with MAF <1% in the GRS. We also excluded index SNVs with poor imputation quality (**r**^2^<0.7) in BBJ, and those with extreme deviation from Hardy-Weinberg equilibrium (**P**<10^-6^) in AoURP. We standardised overall and cluster-specific GRS to have mean zero and unit variance. We tested for association with each outcome/complication via generalised linear regression, under a joint model including all eight cluster-specific GRS, and a model including only the overall GRS. We adjusted association analyses with macrovascular outcomes, conducted in all individuals, for T2D status. We adjusted association analyses with microvascular complications for duration of T2D. In AoURP and BBJ, we adjusted analyses for age, sex, and the first 20 principal components. In AoURP, we defined age as age at last hospital visit. In BBJ, we defined age as age at first record. In BBJ, we also adjusted for recruitment phase and status of the registered common diseases (other than T2D) to account for ascertainment. For CAD, we also conducted sensitivity analyses by including, as an additional covariate, a CAD GRS from the largest published multi-ancestry CAD GWAS^49^. The GRS was constructed from index SNVs for 241 conditionally independent CAD associations, weighted by the multi-ancestry allelic log-OR (ancestry-specific effects were not available), and standardised to have mean zero and unit variance. All association analyses were conducted using the glm function in R.

##### GRS multi-ancestry meta-analyses

For each outcome, we aggregated association summary statistics from the partitioned GRS model and the overall GRS model across ancestries via multivariate fixed-effects meta-analysis (weighting by the inverse covariance matrix)^81^. We also tested for heterogeneity using the multivariate Cochran Q statistic^81^. All meta-analyses were conducted using the R package mvmeta (https://cran.r-project.org/package=mvmeta).

**Data availability.** Genome-wide association summary statistics from the multi-ancestry meta-analysis reported in this study will be made available through the DIAGRAM Consortium website (http://www.diagram-consortium.org/downloads.html) an acceptance of the manuscript.

**Code availability.** Analyses were conducted using publicly available software: UCSC liftOver tool (https://genome.ucsc.edu/cgi-bin/hgLiftOver), MR-MEGA (https://genomics.ut.ee/en/tools), METAL (https://genome.sph.umich.edu/wiki/METAL), PLINKv1.9 (https://www.cog-genomics.org/plink/1.9/), Hail v0.2.107 (https://hail.is/docs/0.2/index.html), SHAPEIT4 (https://odelaneau.github.io/shapeit4/), Minimac4 (https://genome.sph.umich.edu/wiki/Minimac4), PLINKv2.0 (https://www.cog-genomics.org/plink/2.0/). Analyses were also conducted using the following R packages: meta (https://cran.r-project.org/package=meta), ClustImpute (https://cran.r-project.org/package=ClustImpute), NbClust (https://cran.r-project.org/package=NbClust), logistf (https://cran.r-project.org/package=logistf), and mvmeta (https://cran.r-project.org/package=mvmeta).

