## Supplementary Information for "Multi-ancestry genome-wide study in >2.5 million individuals reveals heterogeneity in mechanistic pathways of type 2 diabetes and complications"

**Supplementary Note**

- Supplementary Methods
- Acknowledgements and Funding
- Ethics Statements
- VA Million Veteran Program: Core Acknowledgement for Publications
- Contributors to AMED GRIFIN Diabetes Initiative Japan
- Contributors to Biobank Japan Project
- Penn Medicine BioBank Banner Author List and Contribution Statements
- Regeneron Genetics Center Banner Author List and Contribution Statements
- Contributors to eMERGE Consortium
- Membership of the International Consortium of Blood Pressure
- Membership of the Meta-Analyses of Glucose and Insulin-Related Traits Consortium

**Supplementary Figures**

### Supplementary Methods

**Phenotype derivation in AoURP.** Using EHR data, we filtered downstream analyses to a subset of participants that were classified as T2D cases or controls using a combination of diagnosis codes (ICD-9-CM and ICD-10-CM), drug exposures, and LOINC codes for laboratory test results.

*Derivation of T2D cases and controls.* For T2D cases, we used a previously developed method (https://phekb.org/phenotype/type-2-diabetes-mellitus). Briefly, we considered participants as T2D cases if they fit the following criteria: (a) at least one T2D diagnosis code and at least one drug exposure for T2D medications, unless at least one type 1 diabetes (T1D) diagnosis code; (b) at least one T2D diagnosis code, at least two drug exposures for T1D and T2D medications with a T2D drug exposure occurring at least one day before T1D drug exposure, unless at least one T1D diagnosis code; (c) at least two T2D diagnosis codes and at least one drug exposure for T1D medication, unless at least one T1D diagnosis code; or (d) at least one drug exposures for T2D medications and at least one abnormal laboratory test result (random glucose, fasting glucose, or HbA1c), unless at least one T1D diagnosis codes. For controls, we considered those participants that were free of all diabetes diagnosis codes, including T2D, T1D, and other forms of diabetes. Additionally, we excluded participants that matched criteria (d) from the T2D definition.

For T2D, we used diagnosis codes 250.00, 250.02, 250.20, 250.22, 250.30, 250.32, 250.40, 250.42, 250.50, 250.52, 250.60, 250.62, 250.70, 250.72, 250.80, 250.82, 250.90, 250.92 from ICD-9-CM and E11.00, E11.01, E11.21, E11.29, E11.311, E11.319, E11.36, E11.39, E11.40, E11.51, E11.618, E11.620, E11.621, E11.622, E11.628, E11.630, E11.638, E11.641, E11.649, E11.65, E11.69, E11.8, E11.9 from ICD-10-CM. For T2D drug exposures, we used the following medications: acarbose, acetohexamide, albiglutide, alogliptin, canagliflozin, chlorpropamide, colesevelam, dapagliflozin, dulaglutide, empagliflozin, exenatide, glimepiride, glipizide, glyburide, linagliptin, liraglutide, lixisenatide, metformin, miglitol, nateglinide, pioglitazone, repaglinide, rosiglitazone, saxagliptin, semaglutide, sitagliptin, tolazamide, and troglitazone. Finally, we considered the following abnormal lab results: random glucose (LOINC codes: 2339-0, 2345-7) > 200mg/dl, fasting glucose (LOINC code: 1558-6) ≥ 125mg/dl, and HbA1c (LOINC codes: 4548-4, 17856-6,4549-2, 17855-8) ≥ 6.5%. For T1D, we used diagnosis codes 250.01, 250.03, 250.11, 250.13, 250.21, 250.23, 250.31, 250.33, 250.41, 250.43, 250.51, 250.53, 250.61, 250.63, 250.71, 250.73, 250.81, 250.83, 250.91, 250.93 from ICD-9-CM and E10.10, E10.11, E10.21, E10.29, E10.311, E10.319, E10.36, E10.39, E10.40, E10.51, E10.618, E10.620, E10.621, E10.622, E10.628, E10.630, E10.638, E10.641, E10.649, E10.65, E10.69, E10.8, E10.9 from ICD-10-CM. For T1D drug exposures, we used the following medications: insulin, insulin NPH, insulin aspart, insulin degludec, insulin detemir, insulin glargine, insulin glulisine, insulin lispro, pramlintide. For other forms of diabetes, we used diagnosis codes 249*, 648.0*, 648.8* in ICD-9-CM and E08*, E09*, E13*, O24* in ICD-10-CM.

*Derivation of cases and controls for T2D-related clinical outcomes.* For each T2D-related clinical outcome, we used previously-defined ICD-9-CM and ICD-10-CM diagnosis codes from EHR data to identify cases and controls[1-4]. For macrovascular outcomes (CAD, ischemic stroke, and peripheral artery disease), we defined cases and controls as participants with and without, respectively, the relevant diagnosis codes, irrespective of T2D status. For CAD, we used 410*, 411*, 412*, 413* in ICD-9-CM and I20*, I21*, I22*, I23*, I24*, I25* in ICD-10-CM. For ischemic stroke, we used 433*, 434* in ICD-9-CM and I63* in

ICD-10-CM. For peripheral artery disease, we used 4400, 4402, 4438, 4439 in ICD-9-CM and I70.0, I70.00, I70.01, I70.2, I70.20, I70.21, I70.8, I70.80, I70.9, I70.90, I73.8, I73.9 in ICD-10-CM. For microvascular complications (ESDN and proliferative diabetic retinopathy), we considered only T2D cases. ESDN cases were defined with relevant diagnosis codes for both diabetic nephropathy and end-stage kidney disease (ESKD), and ESDN controls were defined as being free of any diagnosis code for diabetic nephropathy, defined using the AoURP cohort builder. For ESKD, we used 403.01, 403.11, 403.91, 404.02, 404.03, 404.12, 404.13, 404.92, 404.93, 585.6 in ICD-9-CM and I12.0, I13.11, I13.2, N18.6 in ICD-10-CM. For DN, we used E11.21 in ICD-10-CM. Proliferative diabetic retinopathy cases were defined with relevant diagnosis codes. Proliferative diabetic retinopathy controls were defined as being free of any diagnosis code for diabetic retinopathy. For proliferative diabetic retinopathy, we used 362.02 in ICD-9-CM and E08.35*, E09.35*, E10.35*, E11.35*, E13.35* in ICD-10-CM. For diabetic retinopathy, we used 362.0* in ICD-9-CM and E08.31*, E08.32*, E08.33*, E08.34*, E08.35*, E09.31*, E09.32*, E09.33*, E09.34*, E09.35*, E10.31*, E10.32*, E10.33*, E10.34*, E10.35*, E11.31*, E11.32*, E11.33*, E11.34*, E11.35*, E13.31*, E13.32*, E13.33*, E13.34*, E13.35* in ICD-10-CM.


1. Fall, T., Gustafsson, S., Orho-Melander, M. & Ingelsson, E. Genome-wide association study of coronary artery disease among individuals with diabetes: the UK Biobank. *Diabetologia* **61**, 2174-2179 (2018).
2. Malik, R. et al. Multiancestry genome-wide association study of 520,000 subjects identifies 32 loci associated with stroke and stroke subtypes. *Nat. Genet.* **50**, 524-537 (2018).
3. Klarin, D. et al. Genome-wide association study of peripheral artery disease in the Million Veteran Program. *Nat. Med.* **25**, 1274-1279 (2019).
4. Wang, R. et al. Derivation and validation of essential predictors and risk index for early detection of diabetic retinopathy using electronic health records. *J. Clin. Med.* **10**, 1473 (2021).

**Acknowledgements and Funding**

**Anti-aging Study Cohort (AASC)** is supported by the Grant-in-Aid for Scientific Research (20018020, 19659163, 20390185, 23659382, 24390084, 23659352, 25293141, 26670313, 17H04123) from the Ministry of Education, Culture, Sports, Science and Technology of Japan, research grant from the Japan Atherosclerosis Prevention Found, National Cardiovascular Research Grants, and Research Promotion Award from Ehime University.

**All Of Us Research Program (AOURP)** is supported by the National Institutes of Health, Office of the Director: Regional Medical Centers: 1 OT2 OD026549; 1 OT2 OD026554; 1 OT2 OD026557; 1 OT2 OD026556; 1 OT2 OD026550; 1 OT2 OD 026552; 1 OT2 OD026553; 1 OT2 OD026548; 1 OT2 OD026551; 1 OT2 OD026555; IAA #: AOD 16037; Federally Qualified Health Centers: HHSN 263201600085U; Data and Research Center: 5 U2C OD023196; Biobank: 1 U24 OD023121; The Participant Center: U24 OD023176; Participant Technology Systems Center: 1 U24 OD023163; Communications and Engagement: 3 OT2 OD023205; 3 OT2 OD023206; and Community Partners: 1 OT2 OD025277; 3 OT2 OD025315; 1 OT2 OD025337; 1 OT2 OD025276. In addition, the All of Us Research Program would not be possible without the partnership of its participants.

**Atherosclerosis Risk in Communities (ARIC)** study has been funded in whole or in part with Federal funds from the National Heart, Lung, and Blood Institute, National Institutes of Health, Department of Health and Human Services (contract numbers HHSN268201700001I, HHSN268201700002I, HHSN268201700003I, HHSN268201700004I and HHSN268201700005I), R01HL087641, R01HL059367 and R01HL086694; National Human Genome Research Institute contract U01HG004402; and National Institutes of Health contract HHSN268200625226C. The authors thank the staff and participants of the ARIC study for their important contributions. Infrastructure was partly supported by Grant Number UL1RR025005, a component of the National Institutes of Health and NIH Roadmap for Medical Research.

**BioBank Japan (BBJ).** This study was funded by the BioBank Japan project, which is supported by the Ministry of Education, Culture, Sports, Sciences and Technology (MEXT) of Japanese government and the Japan Agency for Medical Research and Development (AMED, grant ID JP21km0605001). AMED GRIFIN Diabetes Initiative Japan was supported by Japan Agency for Medical Research and Development (JP20km0405202, JP21tm0424218). Scarda was supported by AMED under Grant Number 223fa627011.

**Beijing Eye Study (BES)** was supported by National Natural Science Foundation of China (grant 81570835).

**BioMe Biobank (BIOME)** is supported by The Andrea and Charles Bronfman Philanthropies and in part by funding of the NIH (U01HG007417; R56HG010297; X01HL134588). BIOME thanks all participants in the Mount Sinai Biobank, and also thanks all the recruiters who have assisted and continue to assist in data collection and management. BIOME is grateful for the computational resources and staff expertise provided by Scientific Computing at the Icahn School of Medicine at Mount Sinai.

**Vanderbilt University Medical Center's BioVU (BIOVU)** projects are supported by numerous sources: institutional funding, private agencies, and federal grants. These include NIH funded Shared Instrumentation Grant S10OD017985, S10RR025141, and S10OD025092; CTSA grants UL1TR002243, UL1TR000445, and UL1RR024975. Genomic data are also supported by investigator-led projects that include U01HG004798, R01NS032830, RC2GM092618, P50GM115305, U01HG006378, U19HL065962, and R01HD074711. This work was conducted in part using the resources of the Advanced Computing Center for Research and Education at Vanderbilt University, Nashville, TN, supported in part by an S10 instrumentation award (1S10OD023680-01).

**Bangladesh Population Cohort (BPC)** was supported by US National Institute of Environmental Health Sciences Grants P42 ES10349 and P30 ES09089.

**Cardiometabolic Genome Epidemiology (CAGE-AMAGASKI and CAGE-GWAS)** was supported by grants for the Core Research for Evolutional Science and Technology (CREST) from the Japan Science Technology Agency; KAKENHI (Grant-in-Aid for Scientific Research) from the Ministry of Education, Culture, Sports, Science and Technology of Japan; and the Grant and research budget of National Center for Global Health and Medicine (NCGM). CAGE-AMAGASKI thanks Drs. Toshio Ogihara, Yukio Yamori, Akihiro Fujioka, Chikanori Makibayashi, Sekiharu Katsuya, Ken Sugimoto, Kei Kamide, and Ryuichi Morishita and the many physicians of the participating hospitals and medical institutions in Amagasaki Medical Association for their assistance in collecting the DNA samples and accompanying clinical information.

**Cardiometabolic Genome Epidemiology Kita-Nagoya Genomic Epidemiology (CAGE-KING)** was supported in part by Grants-in-Aid from MEXT (nos. 24390169, 16H05250, 15K19242, 16H06277) as well as by a grant from the Funding Program for Next-Generation World-Leading Researchers (NEXT Program, no. LS056).

**Coronary Artery Risk Development in Young Adults (CARDIA)** was conducted and supported by the National Heart, Lung, and Blood Institute (NHLBI) in collaboration with the University of Alabama at Birmingham (HHSN268201800005I & HHSN268201800007I), Northwestern University (HHSN268201800003I), University of Minnesota (HHSN268201800006I), and Kaiser Foundation Research Institute (HHSN268201800004I). CARDIA was also partially supported by the Intramural Research Program of the National Institute on Aging (NIA) and an intra-agency agreement between NIA and NHLBI (AG0005). Genotyping was funded as part of the NHLBI Candidate-gene Association Resource (N01-HC-65226) and the NHGRI Gene Environment Association Studies (GENEVA) (U01-HG004729, U01-HG04424, and U01-HG004446).

**Cleveland Family Study (CFS)** is supported by grants to Case Western Reserve University (NIH HL 46380, M01RR00080) and Brigham and Women's Hospital (K01-HL135405-01, R01-HL113338-04, R35-HL135818-01, 5-R01-HL046380-15 and 5-KL2-RR024990-05).

**China Health and Nutrition Survey (CHNS)** was supported by: the National Institute for Nutrition and Health, the Chinese Center for Disease Control and Prevention; the National Institutes of Health (R01AG065357, R01HD30880, R01HL108427 and R01DK104371); the

Fogarty International Center of the National Institutes of Health (TW009077); the China-Japan Friendship Hospital, the Beijing Municipal Center for Disease Prevention and Control, the China National Health Commission (formerly the Chinese Ministry of Health); the Chinese National Human Genome Center at Shanghai; and the Carolina Population Center (P2CHD050924), The University of North Carolina at Chapel Hill.

**Cardiovascular Health Study (CHS)** was supported by NHLBI contracts HHSN268201200036C, HHSN268200800007C, HHSN268201800001C, N01HC55222, N01HC85079, N01HC85080, N01HC85081, N01HC85082, N01HC85083, N01HC85086, 75N92021D00006; and NHLBI grants U01HL080295, R01HL085251, R01HL087652, R01HL105756, R01HL103612, R01HL120393, and R01HL130114 with additional contribution from the National Institute of Neurological Disorders and Stroke (NINDS). Additional support was provided through R01AG023629 from the National Institute on Aging (NIA). A full list of principal CHS investigators and institutions can be found at CHS-NHLBI.org. The provision of genotyping data was supported in part by the National Center for Advancing Translational Sciences, CTSI grant UL1-TR-001881, and the National Institute of Diabetes and Digestive and Kidney Disease Diabetes Research Center (DRC) grant DK063491 to the Southern California Diabetes Endocrinology Research Center. The content is solely the responsibility of the authors and does not necessarily represent the official views of the National Institutes of Health.

**China Kadoorie Biobank (CKB)** chiefly acknowledges the participants, project staff, and the China National Centre for Disease Control and Prevention (CDC) and its regional offices. China's National Health Insurance provides electronic linkage to all hospital treatment. Funding sources: Baseline survey and first re-survey - Kadoorie Charitable Foundation, Hong Kong; long-term follow-up - UK Wellcome Trust (212946/Z/18/Z, 202922/Z/16/Z, 104085/Z/14/Z, 088158/Z/09/Z), National Natural Science Foundation of China (82192901, 82192904, 82192900), and National Key Research and Development Program of China (2016YFC 0900500, 0900501, 0900504, 1303904); DNA extraction and genotyping – GlaxoSmithKline, and the UK Medical Research Council (MC-PC-13049, MC-PC-14135); core funding for the project to the Clinical Trial Service Unit and Epidemiological Studies Unit at Oxford University - British Heart Foundation (CH/1996001/9454), UK MRC (MC-UU-00017/1, MC-UU-12026/2, MC_U137686851), and Cancer Research UK (C16077/A29186, C500/A16896).

**Cebu Longitudinal Health and Nutrition Survey (CLHNS)** was supported by: US National Institutes of Health grants DK078150, TW005596 and HL085144; pilot funds from RR020649, ES010126, and DK056350; and the Office of Population Studies Foundation.

**Diabetic Cohort and Singapore Prospective Study Program (DC/SP2)** was supported by the individual research grant and clinician scientist award schemes from the National Medical Research Council (NMRC) and the Biomedical Research Council (BMRC) of Singapore, Ministry of Health, Singapore, National University of Singapore and National University Health System, Singapore.

**Durban Diabetes Study and Durban Diabetes Case Control (DDS/DCC)** was supported by: the Wellcome Trust (grant number 098051); the African Partnership for Chronic Disease

Research (Medical Research Council UK partnership grant number MR/K013491/1); the National Institute for Health Research Cambridge Biomedical Research Centre (UK); Novo-Nordisk (South Africa); Sanofi-Aventis (South Africa); MSD Pharmaceuticals (Pty) Ltd (Southern Africa); Servier Laboratories (South Africa); South African Sugar Association; and the Victor Daitz Foundation.

**deCODE genetics (DECODE)** thank the participants in the deCODE study, the staff at deCODE genetics core facilities and the staff at the Research Service Center for their contribution to this work.

**Diabetes Gene Discovery Group (DGDG)** was supported by Genome Canada, Génome Québec, the Canada Foundation for Innovation, the French Government ("Agence Nationale de la Recherche"), the French Region of "Nord Pas De Calais" ("Contrat de Projets État-Région"), and the charities: "Association Française des Diabétiques", "Programme National de Recherche sur le Diabète" and "Association de Langue Française pour l'Etude du Diabète et des Maladies Métaboliques". This study was also supported in part by a grant from the European Union (Integrated Project EuroDia LSHM-CT-2006-518153in the Framework Programme 6 [FP6] of the European Community). This work was supported by grants from the French National Research Agency (ANR-10-LABX-46 [European Genomics Institute for Diabetes] and ANR-10-EQPX-07-01 [LIGAN-PM]). Case and control recruitment was supported by the Fédération Française des Diabetiques, INSERM, CNAMTS, Centre Hospitalier Universitaire Poitiers, La Fondation de France, and the Endocrinology-Diabetology department of the Corbeil-Essonnes Hospital. C. Petit, J.-P. Riveline, and S. Franc were instrumental in recruitment and S. Brunet, F. Bacot, R. Frechette, V. Catudal, M. Deweirder, F. Allegaert, P. Laflamme, P. Lepage, W. Astle, M. Leboeuf, and S. Leroux provided technical assistance. K. Shazand and N. Foisset provided organizational guidance. The D.E.S.I.R. study, which mostly contributed controls, was supported by CNAMTS, Lilly, Novartis Pharma and Sanofi-Aventis, by INSERM ("Réseaux en Santé Publique, Interactions entre les déterminants de la santé"), by "Association Diabète Risque Vasculaire", "Fédération Française de Cardiologie", "Fondation de France", ALFEDIAM, ONIVINS, Ardix Medical, Bayer Diagnostics, Becton Dickinson, Cardionics, Merck Santé, Novo Nordisk, Pierre Fabre, Roche, Topcon. The D.E.S.I.R. Study Group: INSERM U780: B. Balkau, P. Ducimetière, E. Eschwège; INSERM U367: F. Alhenc-Gelas; CHU D'Angers: Y. Gallois, A. Girault; Bichat Hospital: F. Fumeron, M. Marre; Medical Examination Services: Alençon, Angers, Caen, Chateauroux, Cholet, Le Mans, and Tours; Research Institute for General Medicine: J. Cogneau; General practitioners of the region; Cross-Regional Institute for Health: C. Born, E. Caces, M. Cailleau, J. G. Moreau, F. Rakotozafy, J. Tichet, S. Vol. DGDG thank M. Deweider and F. Allegaert for the DNA bank management and are sincerely indebted to all study participants.

**Diabetes Genetics Initiative (DGI)** was supported by the Novartis Institute for BioMedical Research with additional support from The Richard and Susan Smith Family Foundation and American Diabetes Association Pinnacle Program Project Award. The Botnia Study (study subject cohort) was financially supported by the Folkhalsan Research Foundation, the Sigrid Juselius Foundation, Nordic Center of Excellence in Disease Genetics, EU (EXGENESIS), The Academy of Finland, University of Helsinki, Finnish Diabetes Research Foundation, Foundation for Life and Health in Finland, Finnish Medical Society, Helsinki University

Central Hospital Research Foundation, Perklén Foundation, Ollqvist Foundation, Närpes Health Care Foundation, Municipal Heath Care Center and Hospital in Jakobstad and Health Care Centers in Vasa, Närpes and Korsholm. The work in Malmö, Sweden, was also funded by a Linné grant from the Swedish Research Council (349-2006-237). The contribution of the Botnia and Skara research teams is gratefully acknowledged.

**Electronic Medical Records and Genomics Network (EMERGE)** was initiated and funded by NHGRI through the following grants: U01HG006828 (Cincinnati Children's Hospital Medical Center/Boston Children's Hospital); U01HG006830 (Children's Hospital of Philadelphia); U01HG006389 (Essentia Institute of Rural Health, Marshfield Clinic Research Foundation and Pennsylvania State University); U01HG006382 (Geisinger Clinic); U01HG006375 (Group Health Cooperative/University of Washington); U01HG006379 (Mayo Clinic); U01HG006380 (Icahn School of Medicine at Mount Sinai); U01HG006388 (Northwestern University); U01HG006378 (Vanderbilt University Medical Center); and U01HG006385 (Vanderbilt University Medical Center serving as the Coordinating Center). The Northwestern University Enterprise Data Warehouse was funded in part by a grant from the National Center for Research Resources, UL1RR025741. Part of the dataset(s) used for the analyses described were obtained from Vanderbilt University Medical Center's BioVU which is supported by institutional funding and by the Vanderbilt CTSA grant UL1 TR000445 from NCATS/NIH. The eMERGE imputed merged Phase I and Phase II dataset was generated by genotyping centers CIDR (U01HG004438) and the Broad Institute (U01HG004424).

**European Prospective Investigation into Cancer and Nutrition (EPIC-INTERACT)** project (LSHM-CT-2006-037197) is a European-Community funded project under Framework Programme 6. EPIC-INTERACT thank all EPIC participants and staff for their contribution to the study. EPIC-INTERACT thank Nicola Kerrison (MRC Epidemiology Unit, Cambridge) for managing the data for the InterAct Project and staff from the Laboratory Team, Field Epidemiology Team, and Data Functional Group of the MRC Epidemiology Unit in Cambridge, UK, for carrying out sample preparation, DNA provision and quality control, genotyping, and data-handling work. The funders had no role in study design, data collection and analysis, decision to publish, or preparation of the manuscript. GWAS summary statistics from the EPIC-InterAct study are available to download from the Dryad Digital Repository (https://doi.org/10.5061/dryad.qnk98sfcg).

**Epidemiologic Study of the Screenees for Diabetes Reduction Assessment with Ramipril and Rosiglitazone Medication (EPIDREAM)** was funded by a grant from the Canadian Institutes of Health Research University Industry competition with partner funding from the GlaxoSmithKline and Sanofi Aventis Global, Sanofi Aventis Canada, Genome Quebec Innovation Centre, Heart and Stroke Foundation of Canada.

**Estonian Biobank (ESTBB)** was funded by the Estonian Research Council Grant IUT20-60, IUT24-6, PRG687, and the European Union through the European Regional Development Fund Project No. 2014-2020.4.01.15-0012 GENTRANSMED.

**Family Heart Study (FAMHS)** was supported by NIH grants R01-HL-087700 and R01-HL-088215 from NHLBI, and R01-DK-089256 and R01-DK-075681 from NIDDK.

**Framingham Heart Study (FHS)** was conducted and supported by the National Heart, Lung and Blood Institute (NHLBI) in collaboration with Boston University (contracts 75N92019D00031, HHSN268201500001I and N01-HC-25195), and its contract with Affymetrix, Inc for genotyping services (contract number N02-HL-6-4278). The analyses reflect intellectual input and resource development from the Framingham Heart Study investigators participating in the SNP Health Association Resource (SHARe) project. FHS was also supported by: NHLBI R01 HL105756, National Institute for Diabetes and Digestive and Kidney Diseases (NIDDK) R01 DK078616, U01 DK078616, NIDDK K24 DK080140 and American Diabetes Association Mentor-Based Postdoctoral Fellowship Award #7-09-MN-32 (to J.B.M.); and NIDDK K24 DK110550 (to J.C.F.).

**Finland-United States Investigation of NIDDM Genetics (FUSION)** was supported by DK093757, DK072193, DK062370, and ZIA-HG000024.

**German Chronic Kidney Disease (GCKD)** was funded by the German Ministry of Research and Education (Bundesminsterium für Bildung und Forschung, BMBF) and by the Foundation KfH Stiftung Präventivmedizin. Unregistered grants to support the study were provided by Bayer, Fresenius Medical Care and Amgen. Genotyping was supported by Bayer AG.

**Genetic Study of Atherosclerosis Risk (GENESTAR)** was supported by NIH grants through the National Heart, Lung, and Blood Institute (HL49762, HL58625, HL59684, HL071025, U01HL72518, and HL087698) and the National Institute of Nursing Research (NR0224103) and by M01-RR000052 to the Johns Hopkins General Clinical Research Center.

**Genetic Epidemiology Network of Arteriosclerosis (GENOA)** was supported by the National Institutes of Health grant numbers HL054457, HL054464, HL054481, HL087660 and HL119443 from the National Heart, Lung, and Blood Institute. Genotyping was performed at the Mayo Clinic by Stephen Turner, Mariza de Andrade, and Julie Cunningham. GENOA thanks Eric Boerwinkle and Megan Grove from the Human Genetics Center and Institute of Molecular Medicine and Division of Epidemiology, University of Texas Health Science Center, Houston, Texas, USA for their help with genotyping. GENOA also thanks the families that participated in the study.

**Resource for Genetic Epidemiology on Adult Heath and Aging (GERA)** was supported by a grant (RC2 AG033067; PIs Schaefer and Risch) awarded to the Kaiser Permanente Research Program on Genes, Environment, and Health (RPGEH) and the UCSF Institute for Human Genetics. The RPGEH was supported by grants from the Robert Wood Johnson Foundation, the Wayne and Gladys Valley Foundation, the Ellison Medical Foundation, Kaiser Permanente Northern California, and the Kaiser Permanente National and Northern California Community Benefit Programs.

**Genetics of Diabetes and Audit Research in Tayside Scotland (GODARTS)** was funded by The Wellcome Trust Study Cohort Functional Genomics Grant (2004-2008, 072960/Z/03/Z) and The Wellcome Trust Scottish Health Informatics Programme (SHIP, 2009-2012, 086113/Z/08/Z).

**Genetics of Latinos Diabetic Retinopathy (GOLDR)** was supported by grants EY14684 and UL1TR000124.

**Genetic Overlap Between Metabolic and Psychiatric Traits and Teens of Attica: Genes and Environment (GOMAP-TEENAGE)** was funded by the Wellcome Trust (098051) and was also co-financed by the European Union (European Social Fund - ESF) and Greek national funds through the Operational Program "Education and Lifelong Learning" of the National Strategic Reference Framework (NSRF) - Research Funding Program: Heracleitus II. GOMAP-TEENAGE thanks all study participants and their families, as well as all volunteers for their contribution in this study. GOMAP-TEENAGE is grateful to: Georgia Markou, Laiko General Hospital Diabetes Centre; Maria Emetsidou and Panagiota Fotinopoulou, Hippokratio General Hospital Diabetes Centre; Athina Karabela, Dafni Psychiatric Hospital; Eirini Glezou and Marios Mangioros, Dromokaiteio Psychiatric Hospital; Angela Rentari, Harokopio University of Athens; and Danielle Walker, Wellcome Trust Sanger Institute. GOMAP-TEENAGE thanks the Sample Management and Genotyping Facilities staff at the Wellcome Trust Sanger Institute for sample preparation, quality control and genotyping.

**Genomic Research Cohort for CCMB Diabetes Study (GRCCDS)** comprises of various cohorts that are supported by: Council of Scientific Industrial Research (CSIR); Ministry of Science and Technology, Govt. of India, India; and Wellcome Trust, London, UK. GRCCDS is grateful to the patients and subjects who voluntarily participated in the study, and thankfully acknowledge other researchers who have supported the study.

**Health, Aging and Body Composition Study (HABC)** was supported by NIA contracts N01AG62101, N01AG62103, and N01AG62106. The genome-wide association study was funded by NIA grant 1R01AG032098-01A1 to Wake Forest University Health Sciences and genotyping services were provided by the Center for Inherited Disease Research (CIDR). CIDR is fully funded through a federal contract from the National Institutes of Health to The Johns Hopkins University, contract number HHSN268200782096C. This research was supported in part by the Intramural Research Program of the NIH, National Institute on Aging.

**Healthy Aging in Neighborhoods of Diversity Across the Life Span Study (HANDLS)** was supported by the Intramural Research Program of the NIH, National Institute on Aging (project Z01-AG000513 and human subjects' protocol 09 AGN248). Data analyses for HANDLS utilized the high-performance computational resources of the Biowulf Linux cluster at the National Institutes of Health, Bethesda, MD (http://hpc.nih.gov).

**Hispanic Community Health Study/Study of Latinos (HCHS/SOL)** is a collaborative study supported by contracts from the National Heart, Lung, and Blood Institute (NHLBI) to the University of North Carolina (HHSN268201300001I / N01-HC-65233), University of Miami (HHSN268201300004I / N01-HC-65234), Albert Einstein College of Medicine (HHSN268201300002I / N01-HC-65235), University of Illinois at Chicago (HHSN268201300003I / N01-HC-65236 Northwestern Univ), and San Diego State University (HHSN268201300005I / N01-HC-65237). The following Institutes/Centers/Offices have contributed to the HCHS/SOL through a transfer of funds to the NHLBI: National Institute on Minority Health and Health Disparities, National Institute on Deafness and Other

Communication Disorders, National Institute of Dental and Craniofacial Research, National Institute of Diabetes and Digestive and Kidney Diseases, National Institute of Neurological Disorders and Stroke, NIH Institution-Office of Dietary Supplements. The Genetic Analysis Center at the University of Washington was supported by NHLBI and NIDCR contracts (HHSN268201300005C AM03 and MOD03).

**Hong Kong Diabetes Registry (HKDR)** acknowledge support from the Theme-based Research Scheme from the Research Grants Council of the Hong Kong Special Administrative Region, China (Project no: T12-402/13-N), the Research Grants Council Research Impact Fund (R4012-18), the Hong Kong Foundation for Research and Development in Diabetes, the Vice-Chancellor One-off Discretionary Fund, the Focused Innovations Scheme, the Postdoctoral Fellowship Scheme of the Chinese University of Hong Kong, and the Croucher Foundation Senior Medical Research Fellowship.

**Health Professionals' Follow-Up Study (HPFS) and Nurses Health Study (NHS)** acknowledge assistance with data cleaning that was provided by the National Center for Biotechnology Information. Support for collection of datasets and samples was provided by the Collaborative Study on the Genetics of Alcoholism (COGA; U10 AA008401), the Collaborative Genetic Study of Nicotine Dependence (COGEND; P01 CA089392), and the Family Study of Cocaine Dependence (FSCD; R01 DA013423). Funding support for genotyping, which was performed at the Johns Hopkins University Center for Inherited Disease Research, was provided by the NIH GEI (U01HG004438), the National Institute on Alcohol Abuse and Alcoholism, the National Institute on Drug Abuse, and the NIH contract "High throughput genotyping for studying the genetic contributions to human disease" (HHSN268200782096C). The datasets used for the analyses described in this manuscript were obtained from dbGaP at http://www.ncbi.nlm.nih.gov/projects/gap/cgi-bin/study.cgi?study_id=phs000091.v1.p1 through dbGaP accession number phs000091.v1.p.

**Mexican American Hypertension and Insulin Resistance (HTNIR)** was supported by grant HL0597974.

**Howard University Family Study (HUFS)** was supported by National Institutes of Health grants S06GM008016-320107 to CNR and S06GM008016-380111 to AA. Participant enrollment was carried out at the Howard University General Clinical Research Center, supported by National Institutes of Health grant 2M01RR010284. Genotyping support was provided by the Coriell Institute for Medical Research. This research was supported by the Intramural Research Program of the Center for Research on Genomics and Global Health (CRGGH). The CRGGH is supported by the National Human Genome Research Institute, the National Institute of Diabetes and Digestive and Kidney Diseases, the Center for Information Technology, and the Office of the Director at the National Institutes of Health (Z01HG200362).

**Indian Diabetes Consortium (INDICO)** was majorly supported by Council of Scientific and Industrial Research (CSIR), Government of India through CARDIOMED project Grant Number: BSC0122 provided to CSIR-Institute of Genomics and Integrative Biology. INDICO was also partially funded by Department of Science and Technology-PURSE-II

(DST/SR/PURSE II/11) given to Jawaharlal Nehru University. INDICO are very much thankful to all the volunteers who have participated in the study.

**INTERHEART (INTERHEART)** was funded by: the Canadian Institutes of Health Research, the Heart and Stroke Foundation of Ontario, and the International Clinical Epidemiology Network (INCLEN); unrestricted grants from several pharmaceutical companies (with major contributions from AstraZeneca, Novartis, Hoechst Marion Roussel [now Aventis], Knoll Pharmaceuticals [now Abbott], Bristol-Myers Squibb, King Pharma, and Sanofi-Synthelabo); and various national bodies in different countries (see Online Appendix at http://image.thelancet.com/extras/04art8001webappendix2.pdf). Funding sources had no involvement in the study design; in the collection, analysis, and interpretation of data; or the writing of the manuscript.

**Jackson Heart Study (JHS)** is supported and conducted in collaboration with Jackson State University (HHSN268201800013I), Tougaloo College (HHSN268201800014I), the Mississippi State Department of Health (HHSN268201800015I) and the University of Mississippi Medical Center (HHSN268201800010I, HHSN268201800011I and HHSN268201800012I) contracts from the National Heart, Lung, and Blood Institute (NHLBI) and the National Institute on Minority Health and Health Disparities (NIMHD). The authors also wish to thank the staff and participants of the JHS.

**Korean Association Resource (KARE)** was supported by grants from Korea Centers for Disease Control and Prevention (4845–301, 4851–302, 4851–307) and intramural grants from the Korea National Institute of Health (2016-NI73001-00, 2019-NG-053-00). KARE was performed with bioresources from National Biobank of Korea, the Centers for Disease Control and Prevention, Republic of Korea.

**Korean Biobank Array from the Korean Genome and Epidemiology (KoGES) Consortium (KBA)** was supported by grants from Korea Centers for Disease Control and Prevention (4845–301, 4851–302, 4851–307) and intramural grants from the Korea National Institute of Health (2016-NI73001-00, 2019-NG-053-00). KBA was performed with bioresources from National Biobank of Korea, the Centers for Disease Control and Prevention, Republic of Korea. Genotype data were provided by the Collaborative Genome Program for Fostering New Post-Genome Industry (3000-3031b).

**Collaborative Health Research in the Region of Augsburg (KORA)** research platform was initiated and financed by the Helmholtz Zentrum München – German Research Center for Environmental Health, which is funded by the German Federal Ministry of Education and Research and by the State of Bavaria. Furthermore, KORA research was supported within the Munich Center of Health Sciences (MC Health), Ludwig-Maximilians-Universität, as part of LMUinnovativ and by the German Center for Diabetes Research (DZD).

**Los Angeles Latino Eye Study (LALES)** acknowledges funding from NEI grant U10EY011753.

**London Life Sciences Prospective Population (LOLIPOP)** is supported by the National Institute for Health Research (NIHR) Comprehensive Biomedical Research Centre Imperial College Healthcare NHS Trust, the British Heart Foundation (SP/04/002), the Medical

Research Council (G0601966, G0700931), the Wellcome Trust (084723/Z/08/Z, 090532 & 098381) the NIHR (RP-PG-0407-10371), the NIHR Official Development Assistance (ODA, award 16/136/68), the European Union FP7 (EpiMigrant, 279143) and H2020 programs (iHealth-T2D, 643774). LOLIPOP acknowledges support of the MRC-PHE Centre for Environment and Health, and the NIHR Health Protection Research Unit on Health Impact of Environmental Hazards. The work was carried out in part at the NIHR/Wellcome Trust Imperial Clinical Research Facility. The views expressed are those of the author(s) and not necessarily those of the Imperial College Healthcare NHS Trust, the NHS, the NIHR or the Department of Health. LOLIPOP thanks the participants and research staff who made the study possible.

**Mexican American Study of Coronary Artery Disease (MACAD)** was supported by grant HL088457.

**Mexico City (MC)** was supported, in Mexico, by the Fondo Sectorial de Investigación en Salud y Seguridad Social (SSA/IMSS/ISSSTECONACYT, project 150352), Temas Prioritarios de Salud Instituto Mexicano del Seguro Social (2014-FIS/IMSS/PROT/PRIO/14/34), and the Fundación IMSS. MC thanks Jaime Gómez Zamudio and Araceli Méndez Padrón for technical support. In Canada, computations were performed on the GPC supercomputer at the SciNet HPC Consortium. SciNet is funded by: the Canada Foundation for Innovation under the auspices of Compute Canada; the Government of Ontario; Ontario Research Fund - Research Excellence; and the University of Toronto.

**Multi-Ethnic Study of Atherosclerosis (MESA).** MESA and the MESA SHARe projects are conducted and supported by the National Heart, Lung, and Blood Institute (NHLBI) in collaboration with MESA investigators. Support for MESA is provided by contracts 75N92020D00001, HHSN268201500003I, N01-HC-95159, 75N92020D00005, N01-HC-95160, 75N92020D00002, N01-HC-95161, 75N92020D00003, N01-HC-95162, 75N92020D00006, N01-HC-95163, 75N92020D00004, N01-HC-95164, 75N92020D00007, N01-HC-95165, N01-HC-95166, N01-HC-95167, N01-HC-95168, N01-HC-95169, UL1-TR-000040, UL1-TR-001079, and UL1-TR-001420, UL1TR001881, DK063491, and R01HL105756. Funding for SHARe genotyping was provided by NHLBI Contract N02-HL-64278. Genotyping was performed at Affymetrix (Santa Clara, California, USA) and the Broad Institute of Harvard and MIT (Boston, Massachusetts, USA) using the Affymetrix Genome-Wide Human SNP Array 6.0. The authors thank the other investigators, the staff, and the participants of the MESA study for their valuable contributions. A full list of participating MESA investigators and institutes can be found at http://www.mesa-nhlbi.org.

**Metabolic Syndrome in Men (METSIM)** was supported by the Academy of Finland (contract 124243), the Finnish Heart Foundation, the Finnish Diabetes Foundation, Tekes (contract 1510/31/06), and the Commission of the European Community (HEALTH-F2-2007 201681), and the US National Institutes of Health grants DK093757, DK072193, DK062370, and ZIA-HG000024.

**Mass General Brigham Biobank (MGB)** acknowledges the Partners HealthCare System for support of the MGB biobank and MGB patients for providing samples, genomic data, and health information data, as well as research support by NIDDK K24 DK110550 (to J.C.F.), K24 DK080140 (to J.B.M.) and NIDDK K23DK114551 (to M.S.U).

**Michigan Genomics Initiative (MGI)** was supported by NIH research grants HL117626 and HG007022. MGI was supported by internal research funds from the University of Michigan School of Public Health, the University of Michigan Medical School, and the University of Michigan President's Office. MGI are especially grateful to the generosity of all research participants.

**VA Million Veteran Program (MVP).** This research is based on data from the MVP, Office of Research and Development, Veterans Health Administration and was supported by award MVP000. This publication does not represent the views of the Department of Veterans Affairs, the US Food and Drug Administration, or the US Government. This research was also supported by funding from the Department of Veterans Affairs awards I01- BX003362 (P.S.T. and K.-M.C.). K.-M.C. and P.S.T. are supported by the VA Cooperative Studies Program. Research support for this study was generously provided by the Department of Veterans Affairs (VA) Informatics and Computing Infrastructure (VINCI) (VA HSR RES 13-457).

**Nagahama Study (NAGAHAMA)** was supported by a university grant, The Center of Innovation Program, The Global University Project, and a Grant-in-Aid for Scientific Research (25293141, 26670313, 26293198, 17H04182, 17H04126, 17H04123, 18K18450) from the Ministry of Education, Culture, Sports, Science and Technology of Japan, the Practical Research Project for Rare/Intractable Diseases (ek0109070, ek0109070, ek0109196, ek0109348), the Comprehensive Research on Aging and Health Science Research Grants for Dementia R&D (dk0207006, dk0207027), the Program for an Integrated Database of Clinical and Genomic Information (kk0205008), the Practical Research Project for Life-style-related Diseases including Cardiovascular Diseases and Diabetes Mellitus (ek0210066, ek0210096, ek0210116), and the Research Program for Health Behavior Modification by Utilizing IoT (le0110005) from Japan Agency for Medical Research and Development (AMED); Takeda Medical Research Foundation, and Mitsubishi Foundation, Daiwa Securities Health Foundation, and Sumitomo Foundation.

**Netherlands Epidemiology of Obesity (NEO)** thanks all individuals who participated in the study, all participating general practitioners for inviting eligible participants and all research nurses for collection of the data. NEO thank the study group, Pat van Beelen, Petra Noordijk and Ingeborg de Jonge for the coordination, lab and data management of the study. Genotyping was supported by the Centre National de Génotypage (Paris, France), headed by Jean-Francois Deleuze. NEO is supported by the participating Departments, the Division and the Board of Directors of the Leiden University Medical Center, and by the Leiden University, Research Profile Area Vascular and Regenerative Medicine.

**NIDDM-Atherosclerosis Study Hispanic Cohorts (NIDDM)** was supported by grant HL055798.

**Northewestern University Genetics (NUGENE)** was funded by the Northwestern University's Center for Genetic Medicine, Northwestern University, and Northwestern Memorial Hospital. Samples and data used in this study were provided by the NUgene

Project (www.nugene.org). Assistance with phenotype harmonization was provided by the eMERGE Coordinating Center (Grant number U01HG04603). This study was funded through the NIH, NHGRI eMERGE Network (U01HG004609). Funding support for genotyping, which was performed at The Broad Institute, was provided by the NIH (U01HG004424). Assistance with phenotype harmonization and genotype data cleaning was provided by the eMERGE Administrative Coordinating Center (U01HG004603) and the National Center for Biotechnology Information (NCBI). The datasets used for the analyses described in this manuscript were obtained from dbGaP at http://www.ncbi.nlm.nih.gov/gap through dbGaP accession number phs000237.v1.p1.

**Prospective Investigation of the Vasculature in Uppsala Seniors (PIVUS)** was supported by Wellcome Trust Grants (WT098017, WT064890, WT090532), Uppsala University, Uppsala University Hospital, the Swedish Research Council, and the Swedish Heart-Lung Foundation.

**Penn Medicine BioBank (PMBB).** We acknowledge the PMBB for providing data and thank the patient-participants of Penn Medicine who consented to participate in this research program. We would also like to thank the Penn Medicine BioBank team and Regeneron Genetics Center for providing genetic variant data for analysis. The PMBB is approved under IRB protocol# 813913 and supported by Perelman School of Medicine at University of Pennsylvania, a gift from the Smilow family, and the National Center for Advancing Translational Sciences of the National Institutes of Health under CTSA award number UL1TR001878.

**Pakistan Risk of Myocardial Infarction Study (PROMIS)** was funded by the Wellcome Trust, UK, and Pfizer (genotyping) and was supported through funds available to investigators at the Center for Non-Communicable Diseases, Pakistan, and the University of Cambridge, UK (fieldwork). Biomarker assays in PROMIS have been funded through grants awarded by the National Institutes of Health (RC2HL101834 and RC1TW008485) and the Fogarty International (RC1TW008485).

**Prospective Study of Pravastatin in the Elderly at Risk (PROSPER)** was supported by an investigator-initiated grant obtained from Bristol-Myers Squibb. Prof. J.W.J. is an Established Clinical Investigator of the Netherlands Heart Foundation (grant 2001 D 032). Support for genotyping was provided by the seventh framework program of the European commission (grant 223004) and by the Netherlands Genomics Initiative (Netherlands Consortium for Healthy Aging grant 050-060-810).

**Sea Islands Genetic Network Reasons for Geographic and Racial Differences in Stroke (REGARDS)** is supported by cooperative agreement U01 NS041588 co-funded by the National Institute of Neurological Disorders and Stroke (NINDS) and the National Institute on Aging (NIA), National Institutes of Health, Department of Health and Human Service. The content is solely the responsibility of the authors and does not necessarily represent the official views of the NINDS or the NIA. Additional funding was from R01 DK084350 from the National Institutes of Health.

**Ragama Health Study (RHS)** was supported by a grant from the National Center for Global Health and Medicine (NCGM).

**Rotterdam Study (RS)** are grateful to the participants and staff involved in the study, and the participating general practitioners and pharmacists. RS is funded by Erasmus Medical Center and Erasmus University, Rotterdam, Netherlands Organization for the Health Research and Development (ZonMw), the Research Institute for Diseases in the Elderly (RIDE), the Ministry of Education, Culture and Science, the Ministry for Health, Welfare and Sports, the European Commission (DG XII), and the Municipality of Rotterdam.

**Shanghai Breast Cancer Study and Shanghai Women's Health Study (SBCS/SWHS)** was supported in part by US National Institutes of Health grants R01CA64277 and R01CA124558, as well as Ingram Professorship and Research Reward funds from the Vanderbilt University School of Medicine. We want to thank participants and research staff of the study, Regina Courtney for plasma and DNA sample preparation, and Hui Cai, Ben Zhang and Jing He for data processing and analyses.

**Singapore Chinese Eye Study (SCES)** is supported by the National Medical Research Council (NMRC), Singapore (grants 0796/2003, 1176/2008, 1149/2008, STaR/0003/2008, 1249/2010, CG/SERI/2010, CIRG/1371/2013, and CIRG/1417/2015), and Biomedical Research Council (BMRC), Singapore (08/1/35/19/550 and 09/1/35/19/616).

**Starr County Health (SCH)** was supported by grants from the National Institutes of Health (DK073541, DK085501, HL102830 and DK116378) and funds from the State of Texas. SCH thank the field staff in Starr County for their careful collection of these data and are especially grateful to the participants who so graciously cooperated and gave of their time. Starr County Health

**Singapore Chinese Health Study (SCHS)** was supported by the US National Institutes of Health grants R01DK08072, R01CA144034 and UM1CA182876.

**Slim Initiative for Genomic Medicine in the Americas (SIGMA).** This work was conducted as part of the Slim Initiative for Genomic Medicine, a joint U.S.-Mexico project funded by the Carlos Slim Health Institute. The UNAM/INCMNSZ diabetes study was supported by Consejo Nacional de Ciencia y Tecnología grants 138826, 128877, CONACyT- SALUD 2009-01-115250, and a grant from Dirección General de Asuntos del Personal Académico, UNAM, IT 214711. The Diabetes in Mexico Study was supported by Consejo Nacional de Ciencia y Tecnología grant 86867 and by Instituto Carlos Slim de la Salud, A.C. The Mexico City Diabetes Study was supported by National Institutes of Health (NIH) grant R01HL24799 and by the Consejo Nacional de Ciencia y Tenologia grants: 2092, M9303, F677-M9407, 251M, and 2005-C01-14502, SALUD 2010-2-151165. The Multiethnic Cohort was supported by NIH grants CA164973, CA054281, and CA063464.

**Singapore Malay Eye Study (SIMES)** is supported by the National Medical Research Council (NMRC), Singapore (grants 0796/2003, 1176/2008, 1149/2008, STaR/0003/2008, 1249/2010, CG/SERI/2010, CIRG/1371/2013, and CIRG/1417/2015), and Biomedical Research Council (BMRC), Singapore (08/1/35/19/550 and 09/1/35/19/616).

**Singapore Indian Eye Study (SINDI)** is supported by the National Medical Research Council (NMRC), Singapore (grants 0796/2003, 1176/2008, 1149/2008, STaR/0003/2008, 1249/2010, CG/SERI/2010, CIRG/1371/2013, and CIRG/1417/2015), and Biomedical Research Council (BMRC), Singapore (08/1/35/19/550 and 09/1/35/19/616).

**Samsung Medical Center (SMC)** was supported by a grant from Samsung Biomedical Research Institute. Genotyping of the patients and control subjects from SMC was conducted by Duk-Hwan Kim in the Dept. of Molecular Cell Biology, Sungkyunkwan University School of Medicine, and was supported by a grant from Samsung Biomedical Research Institute.

**Seoul National University Hospital (SNUH)** was supported by a grant from the Korea Health Technology R&D Project through the Korea Health Industry Development Institute, funded by the Ministry of Health & Welfare (grant numbers HI15C1595, HI14C0060, HI15C3131).

**Taiwan Metabochip Consortium Zhonghua (TAICHI-G)** was supported by grants from: the National Health Research Institutes, Taiwan (PH-099-PP-03, PH-100-PP-03, and PH-101-PP-03); the National Science Council, Taiwan (NSC 101-2314-B-075A-006-MY3, MOST 104-2314-B-075A-006-MY3, MOST 104-2314-B-075A-007, and MOST 105-2314-B-075A-003); and the Taichung Veterans General Hospital, Taiwan (TCVGH-1020101C, TCVGH-1020102D, TCVGH-1023102B, TCVGH-1023107D, TCVGH-1030101C, TCVGH- 1030105D, TCVGH-1033503C, TCVGH-1033102B, TCVGH-1033108D, TCVGH-1040101C, TCVGH-1040102D, TCVGH-1043504C, and TCVGH-1043104B). TAICHI-G was also supported in part by the National Center for Advancing Translational Sciences (CTSI grant UL1TR001881).

**Taiwan Type 2 Diabetes (TWT2D)** was supported by the GMM Study, Academia Sinica, Taiwan.

**Danish T2D Case-Control Study (UCPH)** was undertaken by the Novo Nordisk Foundation Center for Basic Metabolic Research, which is an independent Research Center, based at the University of Copenhagen, Denmark and partially funded by an unconditional donation from the Novo Nordisk Foundation (www.cbmr.ku.dk, Grant number NNF18CC0034900). Included study samples were supported by the Danish Research Fund and the National Danish Research Fund (The Vejle Diabetes Biobank), the Velux Foundation, The Danish Medical Research Council and Danish Agency for Science, Technology and Innovation (Health 2006); the Danish Research Council, the Danish Centre for Health Technology Assessment and Novo Nordisk Inc. (Inter99), the Timber Merchant Vilhelm Bang's Foundation and the Danish Heart Foundation (Health 2008), TrygFonden, the Lundbeck Foundation and the Novo Nordisk Foundation (NNF15OC0015896, DanFunD).

**UK Biobank (UKBB)** analyses were conducted using the UK Biobank resource under applications 236, 9161, and 10035. This research was supported by the British Heart Foundation (grant SP/13/2/30111). Large-scale comprehensive genotyping of UK Biobank for cardiometabolic traits and diseases: UK CardioMetabolic Consortium (UKCMC).

**Uppsala Longitudinal Study of Adult Men (ULSAM)** was supported by Wellcome Trust Grants (WT098017, WT064890, WT090532), Uppsala University, Uppsala University Hospital, the Swedish Research Council, and the Swedish Heart-Lung Foundation.

**Wake Forest School of Medicine (WFSM)** was supported by NIH grants K99 DK081350, R01 DK066358, R01 DK053591, R01 DK087914, U01 DK105556, R01 HL56266, R01 DK070941 and in part by the General Clinical Research Center of the Wake Forest School of Medicine grant M01 RR07122. Genotyping services were provided by the Center for Inherited Disease Research (CIDR), which is fully funded through a federal contract from the National Institutes of Health to The Johns Hopkins University, contract number HHSC268200782096C.

**Women's Health Initiative (WHI).** The WHI program is funded by the National Heart, Lung, and Blood Institute, National Institutes of Health, U.S. Department of Health and Human Services through 75N92021D00001, 75N92021D00002, 75N92021D00003, 75N92021D00004, 75N92021D00005. Funding for WHI SHARe genotyping was provided by NHLBI Contract N02-HL-64278. The Molecular Epidemiology of Diabetes in the WHI is supported by R01DK125403 (to S.Liu). The contents of this publication are solely the responsibility of the authors and do not necessarily represent the official view of the National Institutes of Health. The funders had no role in study design, data collection and analysis, decision to publish, or preparation of the manuscript. A list of WHI investigators is available at: https://www-whi-org.s3.us-west-2.amazonaws.com/wp-content/uploads/WHI-Investigator-Short-List.pdf.

**Wellcome Trust Case Control Consortium (WTCCC)** analysis and genotyping was supported by: Wellcome Trust funding 090367, 098381, 090532, 083948, 085475, 101630 and 203141; MRC (G0601261); EU (Framework 7) HEALTH-F4-2007-201413; and NIDDK DK098032 and U01-DK105535.

**Personal acknowledgements.** K.Suzuki was supported by Japan Agency for Medical Research and Development (JP21km0405213, JP20km0405202, JP21tm0424218). R.Mandla was supported by NHGRI U01HG011723, 1-19-ICTS-068. A.H.-C. was supported by NHGRI U01HG011723, 1-19-ICTS-068. L.E.P. was supported by R01GM133169, R01HL142302, R01DK127084. P.S. was supported by NHGRI U01HG011723, 1-19-ICTS-068. F.B. was supported by BHF Centre of Research Excellence, Oxford (RE/13/1/30181). W.Zhang acknowledges support from iHealth-T2D, 643774 and the National Institute for Health Research/Wellcome Trust Imperial Clinical Research Facility. R.A.S. acknowledges support from the Medical Research Council Epidemiology Unit (MC_UU_12015/1). D.T. acknowledges funding from US National Institutes of Health grant DK062370. E.J.P. was supported by the Canadian Institutes of Health Research (CIHR) and the Banting and Best Diabetes Center, University of Toronto. M.W. was supported by the Deutsche Forschungsgemeinschaft (DFG, German Research Foundation) – CRC 1453 Project-ID 431984000. C.Sarnowski acknowledges support from NIA R00 AG066849. D.N. acknowledges support from NIEHS grant T32ES013678. S.-H.K. acknowledges funding from Korea Health Technology R&D Project through the Korea Health Industry Development Institute (grant number HI15C3131). A.W. is supported by a PhD studentship funded by the Wellcome Trust. L.S.A. acknowledges support from the National Institute for Health (NIH), the Eunice Kennedy Shriver National Institute of Child Health and Human Development (NICHD) for R01 HD30880, National Institute on Aging (NIA) for R01 AG065357, National Institute of Diabetes and Digestive and Kidney Diseases (NIDDK) for R01 DK104371 and R01 HL108427. C.F.B. acknowledges funding from the Dr. Robert C. and Veronica Atkins Foundation. J.Chen acknowledges Inês Barroso for supervision and support (Wellcome WT098051 and WT206194). J.Danesh holds a British Heart Foundation Professorship and a NIHR Senior Investigator Award, and this work was supported by core funding from the: British Heart Foundation (RG/13/13/30194; RG/18/13/33946) and NIHR Cambridge Biomedical Research Centre (BRC-1215-20014).S.K.D. acknowledges support from the NIH/NIDDK grant R01 DK090111. S.D. acknowledges funding from the US National Institutes of Health Fogarty grant D43 TW009077, the National Institute for Health (NIH), the Eunice Kennedy Shriver National Institute of Child Health and Human Development (NICHD) for R01 HD30880, National Institute on Aging (NIA) for R01 AG065357, National Institute of Diabetes and Digestive and Kidney Diseases (NIDDK) for R01DK104371 and R01HL108427. D.S.E. acknowledges support from the US National Institutes of Health U24AG051129. P.G.-L. acknowledges support from the National Institute for Health (NIH), the Eunice Kennedy Shriver National Institute of Child Health and Human Development (NICHD) for R01 HD30880, National Institute on Aging (NIA) for R01 AG065357, National Institute of Diabetes and Digestive and Kidney Diseases (NIDDK) for R01DK104371 and R01HL108427. A.T.H. acknowledges support from a Wellcome Trust Senior Investigator award (grant number 098395/Z/12/Z). K.Läll acknowledges funding from the Estonian Research Council grant 1911. N.R.L. acknowledges funding from the US National Institutes of Health TW008288. C.M.L. is supported by the Li Ka Shing Foundation, WT-SSI/John Fell funds Oxford, NIHR Oxford Biomedical Research Centre, Widenlife, and NIH (5P50HD028138-27). A.E.L. acknowledges funding from US National Institutes of Health grant DK062370. J.Luan acknowledges support from the Medical Research Council Epidemiology Unit (MC_UU_12015/1). S.Maeda is supported by the grant for Okinawa innovation/eco-system promotion project from the Okinawa prefecture. M.A.N. was supported in part by the Intramural Research Program of the NIH, National Institute on Aging (NIA), National Institutes of Health, Department of Health and Human Services (project number ZO1 AG000535), as well as the National Institute of Neurological Disorders and Stroke (NINDS); participation in this project was part of a competitive contract awarded to Data Tecnica International LLC by the National Institutes of Health to support open science research. Y.O. was supported by JSPS KAKENHI (22H00476), and AMED (JP21gm4010006, JP22km0405211, JP22ek0410075, JP22km0405217, JP22ek0109594, JP223fa627002, JP223fa627010, JP233fa627011), JST Moonshot R&D (JPMJMS2021, JPMJMS2024), Takeda Science Foundation, Bioinformatics Initiative of Osaka University Graduate School of Medicine, Institute for Open and Transdisciplinary Research Initiatives and Center for Infectious Disease Education and Research (CiDER), Osaka University. H.G.P. was supported by R01GM133169, R01HL142302, R01DK127084. N.Sattar is supported by British Heart Foundation Centre of Excellence Grant RE/18/6/34217. N.Shojima was supported by Japan Agency for Medical Research and Development (JP20km0405202, JP21tm0424218). E.W. acknowledges Inês Barroso for supervision and support (Wellcome WT098051 and WT206194), and acknowledges support from the Medical Research Council Epidemiology Unit (MC_UU_12015/1). Y.S.C. acknowledges support from the National Research Foundation of Korea (NRF) Grant funded by the Ministry of Education (NRF-2020R1I1A2075302). E.Ingelsson was supported by NIH/NIDDK 1R01DK106236-01A1. J.-

Y.W. was supported by Academia Sinica GMM Study. R.C.W.M. acknowledges funding from the Research Grants Council Theme-based Research Scheme (T12-402/13-N), the RGC Research Impact Fund (R4012-18), and a Croucher Foundation Senior Medical Research Fellowship. F.S.C. acknowledges support from United States' National Institutes of Health (NIH) grant ZIA-HG000024. K.-S.P. acknowledges funding from Korea Health Technology R&D Project through the Korea Health Industry Development Institute (grant numbers HI15C1595, HI14C0060). R.M.-C. acknowledges support from grants NIH U10 EY 11753 and NIH U10 EY 11753. C.-Y.C. acknowledges funding from the National Medical Research Council (NMRC), Singapore (CSA-SI/0012/2017). J.Dupuis is supported by R01 DK078616 and U01 DK078616. A.Köttgen was supported by the by the Deutsche Forschungsgemeinschaft (DFG, German Research Foundation) KO 3598/5-1 and CRC 1453 Project-ID 431984000. D.W.B. acknowledges support from the US National Institutes of Health U01DK105556 and R01DK66358. K.E.N. acknowledges support by R01HD057194, R01DK122503, R01HG010297, R01HL142302, R01HL143885, R01HG009974, and R01DK101855. D.S. has received funding from NHLBI, NINDS, the British Heart Foundation, Pfizer, Regeneron, Genentech, and Eli Lilly pharmaceuticals. N.J.W. acknowledges support from the Medical Research Council Epidemiology Unit (MC_UU_12015/1). M.O.G. acknowledges support from the US National Institutes of Health grants P30DK063491 and UL1TR001881, as well as the Eris M. Field Chair in Diabetes Research. K.L.M. acknowledges funding from the US National Institutes of Health R01DK072193, R01DK093757, U01DK105561. C.L. acknowledges support from the Medical Research Council Epidemiology Unit (MC_UU_12015/1). R.J.F.L. acknowledges support from R01DK110113, R01DK107786, R01HL142302, and R56HG010297. J.C.F. is a Massachusetts General Hospital Research Scholar and was supported by NIDDK U01 DK105554 and NIDDK K24 DK110550. J.C.D. acknowledges support from United States' National Institutes of Health (NIH) grant ZIA-HG200417. T.Y. was supported by Japan Agency for Medical Research and Development (JP20km0405202, JP21tm0424218). T.Kadowaki was supported by Japan Agency for Medical Research and Development (JP20km0405202, JP21tm0424218). J.C.C. acknowledges support from the Singapore Ministry of Health's National Medical Research Council under its Singapore Translational Research Investigator (STaR) Award (NMRC/STaR/0028/2017), iHealth-T2D 643774, and the National Institute for Health Research/Wellcome Trust Imperial Clinical Research Facility. M.C.Y.N. acknowledges support from the US National Institutes of Health U01DK105556, R01DK66358, and a supplement to R01DK78616-06S1. J.E.B. acknowledges support from R01GM133169, R01HL142302, and R01DK127084. M.I.M. acknowledges funding from: The European Commission (ENGAGE: HEALTH-F4-2007-201413); MRC (G0601261, L020149); National Institutes of Health (RC2-DK088389, DK085545, R01-DK098032, U01-DK105535); Wellcome (083948, 085475, 090367, 090532, 098381, 101630, 203141, 212259). J.B.M. acknowledges funding through NIH grants R01DK078616, U01DK078616 and K24DK080140. C.N.S. was supported by American Heart Association Postdoctoral Fellowship 15POST24470131 and 17POST33650016, and American Diabetes Association 11-22-JDFPM-06. J.M.M. is funded by American Diabetes Association Innovative and Clinical Translational Award 1-19-ICTS-068, and NHGRI U01HG011723. M.B. acknowledges funding from US National Institutes of Health grant DK062370. M.V. acknowledges support from the Corporal Michael J. Crescenz VA Medical Center Research Department. B.F.V. acknowledges support from the NIH/NIDDK (DK126194). A.P.M. acknowledges support from US National Institutes of Health U01DK105535, Versus Arthritis (grant reference 21754), and NIHR Manchester Biomedical Research Centre (NIHR203308).

The views expressed in this article are those of the authors and do not necessarily represent those of: the UK National Health Service, the UK National Institute for Health Research, or the UK Department of Health and Social Care; the US National Heart, Lung, and Blood Institute, the US National Institute of Neurological Disorders and Stroke, the US National Institute on Aging, the US National Institutes of Health, the US Department of Health and Human Services, the US Department of Veterans Affairs, the US Food and Drug Administration, or the US Government.

### Ethics statements

**Anti-aging study cohort (AASC).** The ethics committees of Ehime University Graduate School of Medicine approved all study procedures. Written informed consent was obtained from all participants.

**All Of Us Research Program (AOURP).** All research was conducted under the guidelines defined by the All of Us Ethical Conduct of Research Policy.

**Atherosclerosis Risk in Communities (ARIC).** Institutional Review Board approvals were obtained at all study sites: National Heart, Lung, and Blood Institute, University of North Carolina at Chapel Hill, Wake Forest Baptist Medical Center, University of Mississippi Medical Center, University of Minnesota, and Johns Hopkins University. All participants provided written informed consent.

**Biobank Japan (BBJ).** All participants provided written informed consent as approved by the ethical committees of the RIKEN Yokohama Institute and the Institute of Medical Science, the University of Tokyo. Ethical approvals of AMED GRIFIN Diabetes Initiative Japan were gained from the Ethics Committees of Osaka University and the University of Tokyo.

**Beijing Eye Study (BES).** Approval was obtained from the Medical Ethics Committee of the Beijing Tongren Hospital. All participants gave written informed consent.

**BioMe Biobank (BIOME).** Approval was obtained from the Institutional Review Board at the Icahn School of Medicine at Mount Sinai. All participants provided written informed consent for genomic data sharing.

**Vanderbilt University Medical Center's BioVU (BIOVU).** Analyses of DIAMANTE data at Vanderbilt University Medical Center are approved under IRB #190891 and analysis of BioVU data are approved under IRBs #210163 and #171279. In all three cases, the data analyzed received non-human subject determinations.

**Bangladesh Population Cohort (BPC).** The conduct of the BPC was reviewed and approved by Ethical Committees of the Bangladesh Medical Research Council and Institutional Review Boards of the University of Chicago.

**Cardiometabolic Genome Epidemiology (CAGE-AMAGASKI and CAKE-GWAS).** Approval was obtained from the Institutional Review Boards at the National Center for Global Health and Medicine. All participants provided written informed consent.

**Cardiometabolic Genome Epidemiology (CAGE-KING).** Approval was obtained from the ethics committees of Aichi Gakuin University, Jichi Medical University, Nagoya University and Kyushu University. All participants provided written informed consent.

**Coronary Artery Risk Development in Young Adults (CARDIA).** Participating centers (Northwestern University, University of Alabama Birmingham, University of Minnesota, and

Kaiser Foundation Research Institute) provided ethics approval for the CARDIA study, and all participants provided written informed consent to participate.

**Cleveland Family Study (CFS).** Approval was obtained from the Institutional Review Board of Mass General Brigham (formerly Partners HealthCare). Written informed consent was obtained from all participants.

**China Health and Nutrition Survey (CHNS).** Approval was obtained from the Institutional review Boards at the University of North Carolina at Chapel Hill, the Chinese National Human Genome Center at Shanghai, and the Institute of Nutrition and Food Safety at the China Centers for Disease Control. All participants provided written informed consent.

**Cardiovascular Health Study (CHS).** Approval was obtained from the Institutional Review Boards at Wake Forest University, University of California, Davis, Johns Hopkins, University of Pittsburgh, and the University of Washington, Seattle. All participants provided written informed consent.

**China Kadoorie Biobank (CKB).** All participants provided written informed consent. Ethical approval was obtained from Oxford Tropical Research Ethics Committee (OxTREC) and from the Ethical Review Committees of the Chinese Centre for Disease Control and Prevention and the Chinese Academy of Medical Sciences/Peking Union Medical College.

**Cebu Longitudinal Health and Nutrition Survey (CLHNS).** Written informed consent was obtained from all participants. Study protocols were approved by the University of North Carolina Institutional review Board for the Protection of Human Subjects.

**Diabetic Cohort and Singapore Prospective Study Program (DC/SP2).** Study protocols were approved by the Singapore General Hospital Ethics Committee, and National University of Singapore Institutional Review Board. All participants provided written informed consent.

**Durban Diabetes Study and Durban Diabetes Case Control (DDS/DCC).** Approvals were granted by the Biomedical Research Ethics Committee at the University of KwaZulu-Natal and the UK National Research Ethics Service. All participants provided written informed consent.

**deCODE genetics (DECODE).** The study was approved by the Icelandic National Bioethics Committee (approval no. VSN-16-112) after evaluation by the Icelandic Data Protection Authority. We obtained written informed consent for all participants in this study who donated samples. All data processing complies with the Icelandic Data Protection Authority (no. PV_2017060950ÞS).

**Diabetes Gene Discovery Group (DGDG).** All participants signed informed consent, and the protocol was approved by the French ethics committee.

**Diabetes Genetics Initiative (DGI).** The study was approved by the Ethics Committees of the Helsinki University Hospital, Helsinki, Finland, and Lund University, Sweden.

**Estonian Genome Center of the University of Tartu (EGCUT).** All analyses were approved by the Ethics Review Committee of the University of Tartu. All participants provided written informed consent.

**Electronic Medical Records and Genomics Network (EMERGE).** Approval was obtained from the Institutional Review Boards at Boston Children's Hospital, Children's Hospital of Philadelphia, Cincinnati Children's Hospital Medical Center, Essentia Institute of Rural Health, Geisinger Clinic, Group Health Cooperative, Marshfield Clinic Research Foundation, Mayo Clinic, Icahn School of Medicine at Mount Sinai, Northwestern University, Pennsylvania State University, Vanderbilt University Medical Center, and University of Washington. All participants provided written informed consent.

**European Prospective Investigation into Cancer and Nutrition (EPIC-INTERACT).** The EPIC-InterAct study was approved by the local ethics committee in the participating countries and the Internal Review Board of the International Agency for Research on Cancer. All participants gave written informed consent. The study was coordinated by the Medical Research Council Epidemiology Unit at the University of Cambridge.

**Epidemiologic Study of the Screenees for Diabetes Reduction Assessment with Ramipril and Rosiglitazone Medication (EPIDREAM).** All study participants consented to analysis of blood samples. Approval was granted by the Hamilton Integrated Research Ethics Board, at McMaster University, Hamilton, Canada.

**Family Heart Study (FAMHS).** Approval was obtained from the Institutional Review Board at Washington University, St. Louis. Written informed consent, including consent to participate in genetic studies, was obtained from all participants.

**Framingham Heart Study (FHS).** Approval was obtained from the Institutional review Board of Boston University Medical Campus. All study participants provided written informed consent.

**Finland-United States Investigation of NIDDM Genetics (FUSION).** Approval was obtained from the coordinating Ethics Committee of the Hospital District of Helsinki and Uusimaa. All participants provided written informed consent.

**German Chronic Kidney Disease (GCKD).** All participants provided written informed consent. The study was registered in the national registry for clinical studies (DRKS 00003971) and was approved by local ethics committees.

**Genetic Study of Atherosclerosis Risk (GENESTAR).** Approval was obtained from the Johns Hopkins Medicine Institutional Review Board. All participants gave written informed consent.

**Genetic Epidemiology Network of Arteriosclerosis (GENOA).** Approval was granted by Institutional Review Boards of the University of Michigan, University of Mississippi Medical Center and Mayo Clinic. Written informed consent was obtained from all participants.

**Resource for Genetic Epidemiology on Adult Heath and Aging (GERA).** The Institutional Review Boards for Human Subjects Research of both Kaiser Permanente Medical Care Plan (Northern California Region) and the University of California at San Francisco approved the project.

**Genetics of Diabetes and Audit Research in Tayside Scotland (GODARTS).** Approval was obtained from the Tayside Medical Ethics Committee. Informed consent was obtained for all participants.

**Genetics of Latinos Diabetic Retinopathy (GOLDR).** Approval was granted by the Institutional Review Board of the Lundquist Institute for Biomedical Innovation at Harbor-UCLA Medical Center.

**Genetic Overlap Between Metabolic and Psychiatric Traits and Teens of Attica: Genes and Environment (GOMAP-TEENAGE).** Ethical permission for TEENAGE was obtained from the Bioethics Committee of Harokopio University, Athens. Ethical permission for GOMAP was obtained from the Dromokaiteio Scientific Committee, Dromokaiteio Management Committee, Dafni Scientific Committee, Eginitio Scientific Committee and Harokopio Ethics Committee. All participants of GOMAP-TEENAGE gave written informed consent.

**Genomic Research Cohort for CCMB Diabetes Study (GRCCDS).** Ethics committees of CSIR-Centre for Cellular and Molecular Biology and KEM Hospital and Research Centre approved the project.

**Health, Aging and Body Composition Study (HABC).** The Institutional Review Boards at the University of Memphis and the University of Pittsburgh granted approval to conduct the Health ABC Study, and all participants provided written informed consent.

**Healthy Aging in Neighborhoods of Diversity Across the Life Span Study (HANDLS).** Approval was granted by the National Institutes of Health Institutional Review Board (study number 09AGN248). All participants provided written informed consent.

**Hispanic Community Health Study/Study of Latinos (HCHS/SOL).** Approval was obtained from Institutional Review Boards at the University of North Carolina at Chapel Hill, Albert Einstein College of Medicine, University of Illinois at Chicago, University of Miami, and San Diego State University. All participants provided written informed consent.

**Hong Kong Diabetes Registry (HKDR).** Approval was obtained from the Chinese University of Hong Kong Clinical Research Ethics Committee.

**Health Professionals' Follow-Up Study (HPFS).** Approval was obtained from the Human Research Committee at the Brigham and Women's Hospital. All participants provided written informed consent.

**Mexican American Hypertension and Insulin Resistance (HTNIR).** Approval was granted by Human Subjects Protection Institutional Review Boards at the University of California at Los

Angeles, University of Southern California, Lundquist/LABioMed/Harbor-UCLA and Cedars-Sinai Medical Center.

**Howard University Family Study (HUFS).** All human participants from the HUFS included in the analyses of this manuscript provided written informed consent prior to enrollment. The HUFS study was approved by the Institutional Review Board at Howard University.

**Indian Diabetes Consortium (INDICO).** Approval was obtained by the Human Ethics Committees of All India Institute of Medical Sciences, New Delhi and CSIR-Institute of Genomics and Integrative Biology, New Delhi, India, and was conducted in accordance with the principles of Helsinki Declarations. Informed written consent was obtained from all of participants.

**INTERHEART (INTERHEART).** All study participants consented to analysis of blood samples. Approval was granted by the Hamilton Integrated Research Ethics Board, at McMaster University, Hamilton, Canada.

**Jackson Heart Study (JHS).** Approval was obtained from Institutional Review Boards at Jackson State University, Tougaloo College and the University of Mississippi Medical Center. All participants provided written informed consent.

**Korean Association Resource (KARE).** Approval was granted by the Institutional review Board at the Korean National Institute of Health. All participants provided written informed consent.

**Korean Biobank Array from the Korean Genome and Epidemiology (KoGES) Consortium (KBA).** Approval was granted by the Institutional Review Board of the Korean National Institute of Health. All participants provided written informed consent.

**Collaborative Health Research in the Region of Augsburg (KORA).** Approval was granted by the Ethics Committee of the Medical Association of Bavaria (number 06068). All participants provided informed consent.

**Los Angeles Latino Eye Study (LALES).** Approval was obtained from the Los Angeles County/University of Southern California Institutional Review Board, and Western Institutional Review Board at Southern California Eye Institute. All participants provided written informed consent.

**London Life Sciences Prospective Population (LOLIPOP).** Approval was obtained from the London-Fulham Research Ethics Committee (ref 07/H0712/150). All participants gave an written informed consent.

**Mexican American Study of Coronary Artery Disease (MACAD).** Approval was granted by Human Subjects Protection Institutional Review Boards at the University of California at Los Angeles, University of Southern California, Lundquist/LABioMed/Harbor-UCLA and Cedars-Sinai Medical Center.

**Mexico City (MC).** Approval was obtained from Institutional Review Boards at the Ethics and Scientific Commission members and the AUTHORIZATION is issued with registration number R-2011-785-018 and the Conacyt SALUD-2010-02-150352. In Canada, approval was obtained from the Research Ethics Board from the University of Toronto (Protocol 15770).

**Multi-Ethnic Study of Atherosclerosis (MESA).** Approval was obtained from Institutional Review Boards at the University of Washington, Wake Forest School of Medicine, Northwestern University, University of Minnesota, Columbia University, Johns Hopkins University, Cedars-Sinai Medical Center, and the University of California at Los Angeles.

**Metabolic Syndrome in Men (METSIM).** Approval was granted by the Ethics Committee of the University of Kuopio and the Kuopio University Hospital. All participants gave written informed consent.

**Mass General Brigham Biobank (MGB).** The MGB Biobank protocol and informed consent documents are reviewed annually by the Partners-MGB Institutional Review Board (#2009P002312). All patients who participate in the MGB Biobank are consented for their samples to be linked to their identified clinical information. They have also consented for their information to be used for a broad range of research and for their deidentified information to be shared outside of MGB.

**Michigan Genomics Initiative (MGI).** Approval was granted by the IRBMED Institutional Review Board of the University of Michigan. All participants gave written informed consent.

**VA Million Veteran Program (MVP).** All participating studies were conducted in compliance with the Declaration of Helsinki and comply with all relevant ethical and local regulatory requirements. Specifically, the contributing genetic association studies were approved by the Department of Veteran's Affairs central IRB.

**Nagahama Study (NAGAHAMA).** Approval was granted by the ethics committees of Kyoto University Graduate School of Medicine. Written informed consent was obtained from all participants.

**Netherlands Epidemiology of Obesity (NEO).** Approval was obtained from the Medical Ethics Committee of Leiden University Medical Center. All participants gave written informed consent.

**Nurses Health Study (NHS).** Approval was obtained from the Human Research Committee at the Brigham and Women's Hospital. All participants provided written informed consent**.**

**NIDDM-Atherosclerosis Study Hispanic Cohorts (NIDDM).** Approval was granted by Human Subjects Protection Institutional Review Boards at the University of California at Los Angeles, University of Southern California, City of Hope, Lundquist/LABioMed/Harbor-UCLA and Cedars-Sinai Medical Center.

**Northewestern University Genetics (NUGENE).** Approval was obtained from Institutional Review Boards at Northwestern University and Vanderbilt University.

**Prospective Investigation of the Vasculature in Uppsala Seniors (PIVUS).** Approval was granted by the Ethics Committee of Uppsala University. All participants provided written informed consent.

**Penn Medicine BioBank (PMBB).** All participating studies were conducted in compliance with the Declaration of Helsinki and comply with all relevant ethical and local regulatory requirements. Specifically, the contributing genetic association studies were approved by the IRB of Perelman School of Medicine at the University of Pennsylvania (IRB protocol #813913).

**Pakistan Risk of Myocardial Infarction Study (PROMIS).** The study was approved by the Institutional Review Board of the Center for Non-Communicable Diseases Pakistan and by regional Ethical Review Committees in the different centres across Pakistan involved in the study. Institutional Review Boards at the National Institute of Cardiovascular Disorders, Karachi, Punjab Institute of Cardiology, Lahore, and Tabba Heart Institute, Karachi approved the study. All participants provided written informed consent.

**Prospective Study of Pravastatin in the Elderly at Risk (PROSPER).** Approval was obtained from the Institutional Ethics Review Boards of Cork University (Ireland), Glasgow University (UK) and Leiden University Medical Center (The Netherlands). All participants gave written informed consent.

**Sea Islands Genetic Network Reasons for Geographic and Racial Differences in Stroke (REGARDS).** The REGARDS study protocol was approved by the institutional review boards of each participating institution, and written informed consents were obtained from all participants.

**Ragama Health Study (RHS).** Approval was obtained from Institutional Review Boards at the National Center for Global Health and the University of Kelaniya (P38/09/2006). All participants provided written informed consent.

**Rotterdam Study (RS).** Approval was granted by the Institutional review Board at Erasmus University Medical Center. All participants provided written informed consent.

**Shanghai Breast Cancer Study and Shanghai Women's Health Study (SBCS/SWHS).** Approval was obtained from Institutional review Boards at Vanderbilt University Medical Center and Shanghai Cancer Institute. A written informed consent form was obtained from all study participants.

**Singapore Chinese Eye Study (SCES).** The study adhered to the Declaration of Helsinki. Ethical approval was obtained from the SingHealth Institutional Review Board and National University of Singapore Institutional Review Board. Written informed consent was obtained from all participants.

**Starr County Health (SCH).** All protocols were reviewed and approved by the Institutional Committee for the Protection of Human Subjects (HSC-SPH-02-042). All participants provided written informed consent permitting the collection and sharing of data.

**Singapore Chinese Health Study (SCHS).** Approval was obtained from the Institutional Review Board at the National University of Singapore. All participants provided written informed consent.

**Slim Initiative for Genomic Medicine in the Americas (SIGMA).** Approval was obtained from the Institutional Review Board of the Instituto Nacional de Ciencas Medicas y Nutricion Salvador Zubiran. All participants provided written informed consent.

**Singapore Malay Eye Study (SIMES).** The study adhered to the Declaration of Helsinki. Ethical approval was obtained from the SingHealth Institutional Review Board and National University of Singapore Institutional Review Board. Written informed consent was obtained from all participants.

**Singapore Indian Eye Study (SINDI).** The study adhered to the Declaration of Helsinki. Ethical approval was obtained from the SingHealth Institutional Review Board and National University of Singapore Institutional Review Board. Written informed consent was obtained from all participants.

**Samsung Medical Center (SMC).** Approval was obtained from the Institutional Review Board of the Samsung Medical Center (No. 2004-12-005). All participants provided written informed consent.

**Seoul National University Hospital (SNUH).** The Institutional Review Board of the Biomedical Research Institute at Seoul National University Hospital approved the study protocol (1205–130–411). Written informed consent was obtained from each participant.

**Taiwan Metabochip Consortium Zhonghua (TAICHI-G).** Approval was granted by Institutional Review Boards at Stanford University School of Medicine, Hudson-Alpha Biotechnology Institute, Lundquist/LABioMed/Harbor-UCLA, Cedars-Sinai Medical Center, Taichung Veterans General Hospital, Taipei Veterans General Hospital, National Health Research Institute, Tri-Service General Hospital, and National Taiwan University Hospital.

**Taiwan Type 2 Diabetes (TWT2D).** Approval was obtained from Institutional Review Boards at China Medical University Hospital, Chia-Yi Christian Hospital, and National Taiwan University Hospital.

**Danish T2D Case-Control Study (UCPH).** The studies included in the Danish T2D Case-Control Study (UCPH) were conducted in accordance with the Declaration of Helsinki II and were approved by the local Ethical Committees of Copenhagen County, the Capital Region of Denmark, or the Region of Southern Denmark.

**UK Biobank (UKBB).** Approval was obtained from the North West Centre for Research Ethics Committee (11/NW/0382).

**Uppsala Longitudinal Study of Adult Men (ULSAM).** Approval was granted by the Ethics Committee of Uppsala University. All participants provided written informed consent.

**Wake Forest School of Medicine (WFSM).** Approval was granted by the Institutional Review Board at Wake Forest School of Medicine. All participants provided written informed consent.

**Women's Health Initiative (WHI).** Approval was granted by the Institutional review Board at the Fred Hutchinson Cancer Research Centre in accordance with the US Department of Health and Human Services regulations at 45 CFR 46 (approval number IR# 3467-EXT). All participants provided written informed consent. Additional written consent to review medical records was obtained. The Fred Hutchinson Cancer Research Centre has an approved FWA on file with the Office for Human Research Protections under assurance number 0001920.

**Wellcome Trust Case Control Consortium (WTCCC).** Approval for the study was obtained from Peterborough & Fenland Local Research Ethics Committee, National Research Ethics Service, Leeds (East) Research Ethics Committee, South West Multicentre Research Ethics Committee, Tayside Committee on Medical Research Ethics and Oxford Tropical Research Ethics Committee.

**VA Million Veteran Program: Core Acknowledgement for Publications**

**MVP Program Office**

- Program Director - Sumitra Muralidhar, Ph.D.
  US Department of Veterans Affairs, 810 Vermont Avenue NW, Washington, DC 20420
- Associate Director, Scientific Programs - Jennifer Moser, Ph.D.
  US Department of Veterans Affairs, 810 Vermont Avenue NW, Washington, DC 20420
- Associate Director, Cohort Management & Public Relations - Jennifer E. Deen, B.S.
  US Department of Veterans Affairs, 810 Vermont Avenue NW, Washington, DC 20420

**MVP Executive Committee**

- Co-Chair: J. Michael Gaziano, M.D., M.P.H.
  VA Boston Healthcare System, 150 S. Huntington Avenue, Boston, MA 02130
- Co-Chair: Sumitra Muralidhar, Ph.D.
  US Department of Veterans Affairs, 810 Vermont Avenue NW, Washington, DC 20420
- Jean Beckham, Ph.D.
  Durham VA Medical Center, 508 Fulton Street, Durham, NC 27705
- Kyong-Mi Chang, M.D.
  Philadelphia VA Medical Center, 3900 Woodland Avenue, Philadelphia, PA 19104
- Philip S. Tsao, Ph.D.
  VA Palo Alto Health Care System, 3801 Miranda Avenue, Palo Alto, CA 94304
- Shiuh-Wen Luoh, M.D., Ph.D.
  VA Portland Health Care System, 3710 SW US Veterans Hospital Rd, Portland, OR 97239
  US Department of Veterans Affairs, 810 Vermont Avenue NW, Washington, DC 20420
- Juan P. Casas, M.D., Ph.D., Ex-Officio
  VA Boston Healthcare System, 150 S. Huntington Avenue, Boston, MA 02130

**MVP Principal Investigators**

- J. Michael Gaziano, M.D., M.P.H.
  VA Boston Healthcare System, 150 S. Huntington Avenue, Boston, MA 02130
- Philip S. Tsao, Ph.D.
  VA Palo Alto Health Care System, 3801 Miranda Avenue, Palo Alto, CA 94304

**MVP Operations**

- MVP Executive Director – Juan P. Casas, M.D., Ph.D.
  VA Boston Healthcare System, 150 S. Huntington Avenue, Boston, MA 02130
- Director of Regulatory Affairs – Lori Churby, B.S.
  VA Palo Alto Health Care System, 3801 Miranda Avenue, Palo Alto, CA 94304
- MVP Cohort Management Director – Stacey B. Whitbourne, Ph.D.
  VA Boston Healthcare System, 150 S. Huntington Avenue, Boston, MA 02130

- MVP Recruitment/Enrollment Director - Jessica V. Brewer, M.P.H.
  VA Boston Healthcare System, 150 S. Huntington Avenue, Boston, MA 02130
- Director, VA Central Biorepository, Boston – Mary T. Brophy M.D., M.P.H.
  VA Boston Healthcare System, 150 S. Huntington Avenue, Boston, MA 02130
- Executive Director for MVP Biorepositories - Luis E. Selva, Ph.D.
  VA Boston Healthcare System, 150 S. Huntington Avenue, Boston, MA 02130
- MVP Informatics, Boston – Shahpoor (Alex) Shayan, M.S.
  VA Boston Healthcare System, 150 S. Huntington Avenue, Boston, MA 02130
- Director, MVP Data Operations/Analytics, Boston – Kelly Cho, M.P.H., Ph.D.
  VA Boston Healthcare System, 150 S. Huntington Avenue, Boston, MA 02130
- Director, Center for Computational and Data Science (C-DACS) & Genomics Core – Saiju Pyarajan Ph.D.
  VA Boston Healthcare System, 150 S. Huntington Avenue, Boston, MA 02130
- Director, Molecular Data Core – Philip S. Tsao, Ph.D.
  VA Palo Alto Health Care System, 3801 Miranda Avenue, Palo Alto, CA 94304
- Director, Phenomics Data Core – Kelly Cho, M.P.H, Ph.D.
  VA Boston Healthcare System, 150 S. Huntington Avenue, Boston, MA 02130
- Director, VA Informatics and Computing Infrastructure (VINCI) – Scott L. DuVall, Ph.D.
  VA Salt Lake City Health Care System, 500 Foothill Drive, Salt Lake City, UT 84148
- MVP Coordinating Centers
  - Cooperative Studies Program Clinical Research Pharmacy Coordinating Center, Albuquerque – Todd Connor, Pharm.D.; Dean P. Argyres, B.S., M.S.
    New Mexico VA Health Care System, 1501 San Pedro Drive SE, Albuquerque, NM 87108
  - Genomics Coordinating Center, Palo Alto – Philip S. Tsao, Ph.D.
    VA Palo Alto Health Care System, 3801 Miranda Avenue, Palo Alto, CA 94304
  - MVP Boston Coordinating Center, Boston - J. Michael Gaziano, M.D., M.P.H.
    VA Boston Healthcare System, 150 S. Huntington Avenue, Boston, MA 02130
  - MVP Information Center, Canandaigua – Brady Stephens, M.S.
    Canandaigua VA Medical Center, 400 Fort Hill Avenue, Canandaigua, NY 14424

**Current MVP Local Site Investigators**

- Atlanta VA Medical Center (Peter Wilson, M.D.)
  1670 Clairmont Road, Decatur, GA 30033
- Bay Pines VA Healthcare System (Rachel McArdle, Ph.D.)
  10,000 Bay Pines Blvd Bay Pines, FL 33744
- Birmingham VA Medical Center (Louis Dellitalia, M.D.)
  700 S. 19th Street, Birmingham AL 35233
- Central Western Massachusetts Healthcare System (Kristin Mattocks, Ph.D., M.P.H.)
  421 North Main Street, Leeds, MA 01053
- Cincinnati VA Medical Center (John Harley, M.D., Ph.D.)
  3200 Vine Street, Cincinnati, OH 45220
- Clement J. Zablocki VA Medical Center (Jeffrey Whittle, M.D., M.P.H.)
  5000 West National Avenue, Milwaukee, WI 53295

- VA Northeast Ohio Healthcare System (Frank Jacono, M.D.)
  10701 East Boulevard, Cleveland, OH 44106
- Durham VA Medical Center (Jean Beckham, Ph.D.)
  508 Fulton Street, Durham, NC 27705
- Edith Nourse Rogers Memorial Veterans Hospital (John Wells., Ph.D.)
  200 Springs Road, Bedford, MA 01730
- Edward Hines, Jr. VA Medical Center (Salvador Gutierrez, M.D.)
  5000 South 5th Avenue, Hines, IL 60141
- Veterans Health Care System of the Ozarks (Kathrina Alexander, M.D.)
  1100 North College Avenue, Fayetteville, AR 72703
- Fargo VA Health Care System (Kimberly Hammer, Ph.D.)
  2101 N. Elm, Fargo, ND 58102
- VA Health Care Upstate New York (James Norton, Ph.D.)
  113 Holland Avenue, Albany, NY 12208
- New Mexico VA Health Care System (Gerardo Villareal, M.D.)
  1501 San Pedro Drive, S.E. Albuquerque, NM 87108
- VA Boston Healthcare System (Scott Kinlay, M.B.B.S., Ph.D.)
  150 S. Huntington Avenue, Boston, MA 02130
- VA Western New York Healthcare System (Junzhe Xu, M.D.)
  3495 Bailey Avenue, Buffalo, NY 14215-1199
- Ralph H. Johnson VA Medical Center (Mark Hamner, M.D.)
  109 Bee Street, Mental Health Research, Charleston, SC 29401
- Columbia VA Health Care System (Roy Mathew, M.D.)
  6439 Garners Ferry Road, Columbia, SC 29209
- VA North Texas Health Care System (Sujata Bhushan, M.D.)
  4500 S. Lancaster Road, Dallas, TX 75216
- Hampton VA Medical Center (Pran Iruvanti, D.O., Ph.D.)
  100 Emancipation Drive, Hampton, VA 23667
- Richmond VA Medical Center (Michael Godschalk, M.D.)
  1201 Broad Rock Blvd., Richmond, VA 23249
- Iowa City VA Health Care System (Zuhair Ballas, M.D.)
  601 Highway 6 West, Iowa City, IA 52246-2208
- Eastern Oklahoma VA Health Care System (River Smith, Ph.D.)
  1011 Honor Heights Drive, Muskogee, OK 74401
- James A. Haley Veterans' Hospital (Stephen Mastorides, M.D.)
  13000 Bruce B. Downs Blvd, Tampa, FL 33612
- James H. Quillen VA Medical Center (Jonathan Moorman, M.D., Ph.D.)
  Corner of Lamont & Veterans Way, Mountain Home, TN 37684
- John D. Dingell VA Medical Center (Saib Gappy, M.D.)
  4646 John R Street, Detroit, MI 48201
- Louisville VA Medical Center (Jon Klein, M.D., Ph.D.)
  800 Zorn Avenue, Louisville, KY 40206
- Manchester VA Medical Center (Nora Ratcliffe, M.D.)
  718 Smyth Road, Manchester, NH 03104
- Miami VA Health Care System (Ana Palacio, M.D., M.P.H.)
  1201 NW 16th Street, 11 GRC, Miami FL 33125

- Michael E. DeBakey VA Medical Center (Olaoluwa Okusaga, M.D.)
  2002 Holcombe Blvd, Houston, TX 77030
- Minneapolis VA Health Care System (Maureen Murdoch, M.D., M.P.H.)
  One Veterans Drive, Minneapolis, MN 55417
- N. FL/S. GA Veterans Health System (Peruvemba Sriram, M.D.)
  1601 SW Archer Road, Gainesville, FL 32608
- Northport VA Medical Center (Shing Shing Yeh, Ph.D., M.D.)
  79 Middleville Road, Northport, NY 11768
- Overton Brooks VA Medical Center (Neeraj Tandon, M.D.)
  510 East Stoner Ave, Shreveport, LA 71101
- Philadelphia VA Medical Center (Darshana Jhala, M.D.)
  3900 Woodland Avenue, Philadelphia, PA 19104
- Phoenix VA Health Care System (Samuel Aguayo, M.D.)
  650 E. Indian School Road, Phoenix, AZ 85012
- Portland VA Medical Center (David Cohen, M.D.)
  3710 SW U.S. Veterans Hospital Road, Portland, OR 97239
- Providence VA Medical Center (Satish Sharma, M.D.)
  830 Chalkstone Avenue, Providence, RI 02908
- Richard Roudebush VA Medical Center (Suthat Liangpunsakul, M.D., M.P.H.)
  1481 West 10th Street, Indianapolis, IN 46202
- Salem VA Medical Center (Kris Ann Oursler, M.D.)
  1970 Roanoke Blvd, Salem, VA 24153
- San Francisco VA Health Care System (Mary Whooley, M.D.)
  4150 Clement Street, San Francisco, CA 94121
- South Texas Veterans Health Care System (Sunil Ahuja, M.D.)
  7400 Merton Minter Boulevard, San Antonio, TX 78229
- Southeast Louisiana Veterans Health Care System (Joseph Constans, Ph.D.)
  2400 Canal Street, New Orleans, LA 70119
- Southern Arizona VA Health Care System (Paul Meyer, M.D., Ph.D.)
  3601 S 6th Avenue, Tucson, AZ 85723
- Sioux Falls VA Health Care System (Jennifer Greco, M.D.)
  2501 W 22nd Street, Sioux Falls, SD 57105
- St. Louis VA Health Care System (Michael Rauchman, M.D.)
  915 North Grand Blvd, St. Louis, MO 63106
- Syracuse VA Medical Center (Richard Servatius, Ph.D.)
  800 Irving Avenue, Syracuse, NY 13210
- VA Eastern Kansas Health Care System (Melinda Gaddy, Ph.D.)
  4101 S 4th Street Trafficway, Leavenworth, KS 66048
- VA Greater Los Angeles Health Care System (Agnes Wallbom, M.D., M.S.)
  11301 Wilshire Blvd, Los Angeles, CA 90073
- VA Long Beach Healthcare System (Timothy Morgan, M.D.)
  5901 East 7th Street Long Beach, CA 90822
- VA Maine Healthcare System (Todd Stapley, D.O.)
  1 VA Center, Augusta, ME 04330
- VA New York Harbor Healthcare System (Peter Liang, M.D., M.P.H.)
  423 East 23rd Street, New York, NY 10010

- VA Pacific Islands Health Care System (Daryl Fujii, Ph.D.)
  459 Patterson Rd, Honolulu, HI 96819
- VA Palo Alto Health Care System (Philip Tsao, Ph.D.)
  3801 Miranda Avenue, Palo Alto, CA 94304-1290
- VA Pittsburgh Health Care System (Patrick Strollo, Jr., M.D.)
  University Drive, Pittsburgh, PA 15240
- VA Puget Sound Health Care System (Edward Boyko, M.D.)
  1660 S. Columbian Way, Seattle, WA 98108-1597
- VA Salt Lake City Health Care System (Jessica Walsh, M.D.)
  500 Foothill Drive, Salt Lake City, UT 84148
- VA San Diego Healthcare System (Samir Gupta, M.D., M.S.C.S.)
  3350 La Jolla Village Drive, San Diego, CA 92161
- VA Sierra Nevada Health Care System (Mostaqul Huq, Pharm.D., Ph.D.)
  975 Kirman Avenue, Reno, NV 89502
- VA Southern Nevada Healthcare System (Joseph Fayad, M.D.)
  6900 North Pecos Road, North Las Vegas, NV 89086
- VA Tennessee Valley Healthcare System (Adriana Hung, M.D., M.P.H.)
  1310 24th Avenue, South Nashville, TN 37212
- Washington DC VA Medical Center (Jack Lichy, M.D., Ph.D.)
  50 Irving St, Washington, D. C. 20422
- W.G. (Bill) Hefner VA Medical Center (Robin Hurley, M.D.)
  1601 Brenner Ave, Salisbury, NC 28144
- White River Junction VA Medical Center (Brooks Robey, M.D.)
  163 Veterans Drive, White River Junction, VT 05009
- William S. Middleton Memorial Veterans Hospital (Prakash Balasubramanian, M.D.)
  2500 Overlook Terrace, Madison, WI 53705

**Contributors to AMED GRIFIN Diabetes Initiative Japan**

Ken Suzuki. Centre for Genetics and Genomics Versus Arthritis, Centre for Musculoskeletal Research, Division of Musculoskeletal and Dermatological Sciences, University of Manchester, Manchester, UK. Department of Diabetes and Metabolic Diseases, Graduate School of Medicine, The University of Tokyo, Tokyo, Japan. Department of Statistical Genetics, Osaka University, Graduate School of Medicine, Suita, Japan.

Kyoto Sonehara. Department of Statistical Genetics, Osaka University Graduate School of Medicine, Suita, Japan. Integrated Frontier Research for Medical Science Division, Institute for Open and Transdisciplinary Research Initiatives, Osaka University, Suita, Japan.

Shinichi Namba. Department of Statistical Genetics, Osaka University, Graduate School of Medicine, Suita, Japan.

Kenichi Yamamoto. Department of Statistical Genetics, Osaka University Graduate School of Medicine, Suita, Japan. Department of Pediatrics, Osaka University Graduate School of Medicine, Suita, Japan. Laboratory of Statistical Immunology, Immunology Frontier Research Center (WPI-IFReC), Osaka University, Suita, Japan.

Nobuhiro Shojima. Department of Diabetes and Metabolic Diseases, Graduate School of Medicine, The University of Tokyo, Tokyo, Japan.

Momoko Horikoshi. Laboratory for Genomics of Diabetes and Metabolism, RIKEN Center for Integrative Medical Sciences, Kanagawa, Japan.

Shiro Maeda. Department of Advanced Genomic and Laboratory Medicine, Graduate School of Medicine, University of the Ryukyus, Okinawa, Japan. Division of Clinical Laboratory and Blood Transfusion, University of the Ryukyus Hospital, Okinawa, Japan Laboratory for Genomics of Diabetes and Metabolism, RIKEN Center for Integrative Medical Sciences, Kanagawa, Japan.

Koichi Matsuda. Department of Computational Biology and Medical Sciences, Graduate School of Frontier Sciences, The University of Tokyo, Tokyo, Japan

Yukinori Okada. Department of Statistical Genetics, Osaka University Graduate School of Medicine, Suita, Japan. Department of Genome Informatics, Graduate School of Medicine, the University of Tokyo, Tokyo, Japan. Laboratory for Systems Genetics, RIKEN Center for Integrative Medical Sciences, Kanagawa, Japan. Laboratory of Statistical Immunology, Immunology Frontier Research Center (WPI-IFReC), Osaka University, Suita, Japan.

Toshimasa Yamauchi. Department of Diabetes and Metabolic Diseases, Graduate School of Medicine, The University of Tokyo, Tokyo, Japan.

Takashi Kadowaki. Department of Diabetes and Metabolic Diseases, Graduate School of Medicine, The University of Tokyo, Tokyo, Japan. Toranomon Hospital, Tokyo, Japan.

**Contributors to Biobank Japan Project**

Koichi Matsuda. Laboratory of Genome Technology, Human Genome Center, Institute of Medical Science, The University of Tokyo, Tokyo, Japan. Laboratory of Clinical Genome Sequencing, Graduate School of Frontier Sciences, The University of Tokyo, Tokyo, Japan.

Yuji Yamanashi. Division of Genetics, The Institute of Medical Science, The University of Tokyo, Tokyo, Japan.

Yoichi Furukawa. Division of Clinical Genome Research, Institute of Medical Science, The University of Tokyo, Tokyo, Japan.

Takayuki Morisaki. Division of Molecular Pathology, IMSUT Hospital Department of Internal Medicine, Institute of Medical Science, The University of Tokyo, Tokyo, Japan.

Yoshinori Murakami. Department of Cancer Biology, Institute of Medical Science, The University of Tokyo, Tokyo, Japan.

Yoichiro Kamatani. Laboratory of Complex Trait Genomics, Graduate School of Frontier Sciences, The University of Tokyo, Tokyo, Japan. Laboratory of Clinical Genome Sequencing, Graduate School of Frontier Sciences, The University of Tokyo, Tokyo, Japan.

Kaori Muto. Department of Public Policy, Institute of Medical Science, The University of Tokyo, Tokyo, Japan.

Akiko Nagai. Department of Public Policy, Institute of Medical Science, The University of Tokyo, Tokyo, Japan.

Wataru Obara. Department of Urology, Iwate Medical University, Iwate, Japan.

Ken Yamaji. Department of Internal Medicine and Rheumatology, Juntendo University Graduate School of Medicine, Tokyo, Japan.

Kazuhisa Takahashi. Department of Respiratory Medicine, Juntendo University Graduate School of Medicine, Tokyo, Japan.

Satoshi Asai. Division of Pharmacology, Department of Biomedical Science, Nihon University School of Medicine, Tokyo, Japan. Division of Genomic Epidemiology and Clinical Trials, Clinical Trials Research Center, Nihon University. School of Medicine, Tokyo, Japan.

Yasuo Takahashi. Division of Genomic Epidemiology and Clinical Trials, Clinical Trials Research Center, Nihon University School of Medicine, Tokyo, Japan.

Takao Suzuki. Tokushukai Group, Tokyo, Japan.

Nobuaki Sinozaki. Tokushukai Group, Tokyo, Japan.

Hiroki Yamaguchi. Department of Hematology, Nippon Medical School, Tokyo, Japan.

Shiro Minami. Department of Bioregulation, Nippon Medical School, Kawasaki, Japan.

Shigeo Murayama. Tokyo Metropolitan Geriatric Hospital and Institute of Gerontology, Tokyo, Japan.

Kozo Yoshimori. Fukujuji Hospital, Japan Anti-Tuberculosis Association, Tokyo, Japan.

Satoshi Nagayama. The Cancer Institute Hospital of the Japanese Foundation for Cancer Research, Tokyo, Japan.

Daisuke Obata. Center for Clinical Research and Advanced Medicine, Shiga University of Medical Science, Shiga, Japan.

Masahiko Higashiyama. Department of General Thoracic Surgery, Osaka International Cancer Institute, Osaka, Japan.

Akihide Masumoto. Iizuka Hospital, Fukuoka, Japan.

Yukihiro Koretsune. National Hospital Organization Osaka National Hospital, Osaka, Japan.

**<u>Penn Medicine BioBank Banner Author List and Contribution Statements</u>**

**PMBB Leadership Team**
Daniel J. Rader, M.D., Marylyn D. Ritchie, Ph.D., Michael D. Feldman M.D.

Contribution: All authors contributed to securing funding, study design and oversight. All authors reviewed the final version of the manuscript.

**Patient Recruitment and Regulatory Oversight**
JoEllen Weaver, Nawar Naseer, Ph.D., M.P.H., Afiya Poindexter, Ashlei Brock, Khadijah Hu-Sain, Yi-An Ko

Contributions: JW manages patient recruitment and regulatory oversight of study. N.N. manages participant engagement, assists with regulatory oversight, and researcher access. A.P., A.B., K.H., Y.K. perform recruitment and enrollment of study participants.

**Lab Operations**
JoEllen Weaver, Meghan Livingstone, Fred Vadivieso, Ashley Kloter, Stephanie DerOhannessian, Teo Tran, Linda Morrel, Ned Haubein, Joseph Dunn

Contribution: J.W., M.L., F.V., S.D. conduct oversight of lab operations. M.L., F.V., A.K., S.D., T.T., L.M. perform sample processing. N.H., J.D. are responsible for sample tracking and the laboratory information management system.

**Clinical Informatics**
Anurag Verma, Ph.D., Colleen Morse, M.S., Marjorie Risman, M.S., Renae Judy, B.S.

Contribution: All authors contributed to the development and validation of clinical phenotypes used to identify study subjects and (when applicable) controls.

**Genome Informatics**
Anurag Verma Ph.D., Shefali S. Verma, Ph.D., Yuki Bradford, M.S., Scott Dudek, M.S., Theodore Drivas, M.D., PH.D.

Contribution: A.V., S.S.V. are responsible for the analysis, design, and infrastructure needed to quality control genotype and exome data. Y.B. performs the analysis. T.D. and A.V. provides variant and gene annotations and their functional interpretation of variants.

**Regeneron Genetics Center Banner Author List and Contribution Statements**

**RGC Management and Leadership Team**
Goncalo Abecasis, D.Phil., Aris Baras, M.D., Michael Cantor, M.D., Giovanni Coppola, M.D., Andrew Deubler, Aris Economides, Ph.D., Luca A. Lotta, M.D., Ph.D., John D. Overton, Ph.D., Jeffrey G. Reid, Ph.D., Katherine Siminovitch, M.D., Alan Shuldiner, M.D.

**Sequencing and Lab Operations**
Christina Beechert, Caitlin Forsythe, M.S., Erin D. Fuller, Zhenhua Gu, M.S., Michael Lattari, Alexander Lopez, M.S., John D. Overton, Ph.D., Maria Sotiropoulos Padilla, M.S., Manasi Pradhan, M.S., Kia Manoochehri, B.S., Thomas D. Schleicher, M.S., Louis Widom, Sarah E. Wolf, M.S., Ricardo H. Ulloa, B.S.

**Clinical Informatics**
Amelia Averitt, Ph.D., Nilanjana Banerjee, Ph.D., Michael Cantor, M.D., Dadong Li, Ph.D., Sameer Malhotra, M.D., Deepika Sharma, M.H.I., Jeffrey Staples, Ph.D.

**Genome Informatics**
Xiaodong Bai, Ph.D., Suganthi Balasubramanian, Ph.D., Suying Bao, Ph.D., Boris Boutkov, Ph.D., Siying Chen, Ph.D., Gisu Eom, B.S., Lukas Habegger, Ph.D., Alicia Hawes, B.S., Shareef Khalid, Olga Krasheninina, M.S., Rouel Lanche, B.S., Adam J. Mansfield, B.A., Evan K. Maxwell, Ph.D., George Mitra, B.A., Mona Nafde, M.S., Sean O'Keeffe, Ph.D., Max Orelus, B.B.A., Razvan Panea, Ph.D., Tommy Polanco, B.A., Ayesha Rasool, M.S., Jeffrey G. Reid, Ph.D., William Salerno, Ph.D., Jeffrey C. Staples, Ph.D., Kathie Sun, Ph.D.

**Analytical Genomics and Data Science**
Goncalo Abecasis, D.Phil., Joshua Backman, Ph.D., Amy Damask, Ph.D., Lee Dobbyn, Ph.D., Manuel Allen Revez Ferreira, Ph.D., Arkopravo Ghosh, M.S., Christopher Gillies, Ph.D., Lauren Gurski, B.S., Eric Jorgenson, Ph.D., Hyun Min Kang, Ph.D., Michael Kessler, Ph.D., Jack Kosmicki, Ph.D., Alexander Li, Ph.D., Nan Lin, Ph.D., Daren Liu, M.S., Adam Locke, Ph.D., Jonathan Marchini, Ph.D., Anthony Marcketta, M.S., Joelle Mbatchou, Ph.D., Arden Moscati, Ph.D., Charles Paulding, Ph.D., Carlo Sidore, Ph.D., Eli Stahl, Ph.D., Kyoko Watanabe, Ph.D., Bin Ye, Ph.D., Blair Zhang, Ph.D., Andrey Ziyatdinov, Ph.D.

**Therapeutic Area Genetics**
Ariane Ayer, B.S., Aysegul Guvenek, Ph.D., George Hindy, Ph.D., Giovanni Coppola, M.D., Jan Freudenberg, M.D., Jonas Bovijn M.D., Katherine Siminovitch, M.D., Kavita Praveen, Ph.D., Luca A. Lotta, M.D., Manav Kapoor, Ph.D., Mary Haas, Ph.D., Moeen Riaz, Ph.D., Niek Verweij, Ph.D., Olukayode Sosina, Ph.D., Parsa Akbari, Ph.D., Priyanka Nakka, Ph.D., Sahar Gelfman, Ph.D., Sujit Gokhale, B.E., Tanima De, Ph.D., Veera Rajagopal, Ph.D., Alan Shuldiner, M.D., Bin Ye, Ph.D., Gannie Tzoneva, Ph.D., Juan Rodriguez-Flores, Ph.D.

**Research Program Management and Strategic Initiatives**
Esteban Chen, M.S., Marcus B. Jones, Ph.D., Michelle G. LeBlanc, Ph.D., Jason Mighty, Ph.D., Lyndon J. Mitnaul, Ph.D., Nirupama Nishtala, Ph.D., Nadia Rana, Ph.D., Jaimee Hernandez

**Contributors to eMERGE Consortium**

Debra Abrams[3], Samuel E Adunyah[4], Ladia Albertson-Junkans[5], Berta Almoguera[6], Darren C Ames[7], Paul Appelbaum[8], Samuel Aronson[9], Sharon Aufox[10], Lawrence J Babb[11], Adithya Balasubramanian[1,12], Hana Bangash[13], Melissa Basford[14], Lisa Bastarache[15], Samantha Baxter[11], Meckenzie Behr[3], Barbara Benoit[16], Elizabeth Bhoj[3], Suzette J Bielinski[17], Sarah T Bland[15], Carrie Blout[18], Kenneth Borthwick[19], Erwin P Bottinger[20], Mark Bowser[21], Harrison Brand[22], Murray Brilliant[23], Wendy Brodeur[24], Pedro Caraballo[25], David Carrell[5], Andrew Carroll[26], Lisa Castillo[27], Victor Castro[28], Gauthami Chandanavelli[1], Theodore Chiang[29], Rex L Chisholm[30], Kurt D Christensen[31], Wendy Chung[32], Christopher G Chute[33], Brittany City[14], Beth L Cobb[34], John J Connolly[3], Paul Crane[35], Katherine Crew[36], David R Crosslin[37], Jyoti Dayal[38], Mariza De Andrade[17], Jessica De la Cruz[1,12], Josh C Denny[39], Shawn Denson[1,2], Tim DeSmet[11], Ozan Dikilitas[13], Michael J Dinsmore[11], Sheila Dodge[11], Phil Dunlea[11], Todd L Edwards[40], Christine M Eng[12], David Fasel[41], Alex Fedotov[42], Qiping Feng[43], Mark Fleharty[11], Andrea Foster[1,2], Robert Freimuth[44], Christopher Friedrich[11], Stephanie M Fullerton[45], Birgit Funke[46], Stacey Gabriel[24], Vivian Gainer[47], Ali Gharavi[48], Richard A Gibbs[1,12], Andrew M Glazer[49], Joseph T Glessner[50], Jessica Goehringer[51], Adam S Gordon[52,53], Chet Graham[54], Robert C Green[55], Justin H Gundelach[13], Heather S Hain[56], Hakon Hakonarson[57], Maegan V Harden[24], John Harley[58], Margaret Harr[59], Andrea Hartzler[60], M Geoffrey Hayes[61], Scott Hebbring[62], Nora Henrikson[63], Andrew Hershey[64], Christin Hoell[30], Ingrid Holm[65], Kayla M Howell[14], George Hripcsak[41,66], Jianhong Hu[1], Elizabeth Duffy Hynes[21], Gail P Jarvik[52,67], Joy C Jayaseelan[1], Yunyun Jiang[1,12], Yoonjung Yoonie Joo[68], Sheethal Jose[38], Navya Shilpa Josyula[69], Anne E Justice[70], Sara E Kalla[1], Divya Kalra[1], Elizabeth W Karlson[71], Brendan J Keating[72], Melissa A Kelly[73], Eimear E Kenny[74], Dustin Key[5], Krzysztof Kiryluk[75], Terrie Kitchner[23], Barbara Klanderman[76], Eric Klee[77], David C Kochan[78], Viktoriya Korchina[1], Leah Kottyan[79], Christie Kovar[1], Emily Kudalkar[54], Alanna Kulchak Rahm[80], Iftikhar J Kullo[81], Philip Lammers[82], Eric B Larson[83], Matthew S Lebo[84], Magalie Leduc[85], Ming Ta Lee[86], Niall J Lennon[24], Kathleen A Leppig[87], Nancy D Leslie[88], Rongling Li[89], Wayne H Liang[90], Chiao-Feng Lin[91], Jodell E Linder[14], Noralane M Lindor[92], Todd Lingren[93], James G Linneman[23], Cong Liu[94], Wen Liu[1], Xiuping Liu[1], John Lynch[95], Hayley Lyon[96], Alyssa Macbeth[97], Harshad Mahadeshwar[1], Lisa Mahanta[98], Bradley Malin[99], Teri Manolio[38], Maddalena Marasa[100], Keith Marsolo[101], Michelle L McGowan[102], Elizabeth McNally[53], Jim Meldrim[24], Frank Mentch[3], Hila Milo Rasouly[103], Jonathan Mosley[104], Shubhabrata Mukherjee[35], Thomas E Mullen[24], Jesse Muniz[1], David R Murdock[1,12], Shawn Murphy[105], Mullai Murugan[106], Donna Muzny[107], Melanie F Myers[108], Bahram Namjou[34,109], Yizhao Ni[110], Robert C Onofrio[24], Aniwaa Owusu Obeng[111,112], Thomas N Person[113], Josh F Peterson[114], Lynn Petukhova[115], Cassandra J Pisieczko[116], Siddharth Pratap[117], Cynthia A Prows[118], Megan J Puckelwartz[119], Ritika Raj[1], James D Ralston[120], Arvind Ramaprasan[5], Andrea Ramirez[121], Luke Rasmussen[122], Laura Rasmussen-Torvik[123], Soumya Raychaudhuri[124], Heidi L Rehm[125], Marylyn D Ritchie[126], Catherine Rives[127], Beenish Riza[128], Dan M Roden[129], Elisabeth A Rosenthal[130], Avni Santani[131], Dan Schaid[17], Steven Scherer[1,12], Stuart Scott[132], Aaron Scrol[133], Soumitra Sengupta[134], Ning Shang[41], Himanshu Sharma[135], Richard R Sharp[136], Rajbir Singh[137], Patrick M A Sleiman[138], Kara Slowik[139], Joshua C Smith[140], Maureen E Smith[141], Duane T Smoot[142], Jordan W Smoller[143], Sunghwan Sohn[144], Ian B Stanaway[37], Justin Starren[145], Mary Stroud[15], Jessica Su[146], Casey Overby Taylor[147], Kasia Tolwinski[148], Sara L Van Driest[149,150], Sean M Vargas[151], Matthew Varugheese[152], David Veenstra[153], Eric Venner[1,12], Miguel Verbitsky[154], Gina Vicente[155], Michael Wagner[156], Kimberly Walker[157], Theresa Walunas[158], Liwen

Wang[159], Qiaoyan Wang[160], Wei-Qi Wei[15], Scott T Weiss[161], Quinn S Wells[162], Chunhua Weng[163], Peter S White[164], Georgia L Wiesner[165], Ken L Wiley Jr[38], Janet L Williams[166], Marc S Williams[167], Michael W Wilson[24], Leora Witkowski[168], Laura Allison Woods[14], Betty Woolf[24], Tsung-Jung Wu[1], Julia Wynn[169], Yaping Yang[170], Victoria Yi[1], Ge Zhang[171,172], Lan Zhang[1], Hana Zouk[173].

[1]Human Genome Sequencing Center, Baylor College of Medicine, Houston, TX, USA. [2]Department of Molecular and Human Genetics, Baylor College of Medicine, Houston, TX, USA. [3]Center for Applied Genomics, Children's Hospital of Philadelphia, Philadelphia, PA, USA. [4]Department of Biochemistry and Cancer Biology, Meharry Medical College, Nashville, TN, USA. [5]Kaiser Permanente of WA Health Research Institute, Seattle, WA, USA. [6]Center for Applied Genomics, Children's Hospital of Philadelphia, Philadelphia, PA, USA. [7]DNAnexus Inc, Mountain View, CA, USA. [8]Department of Psychiatry, Columbia University, New York State Psychiatric Institute, NYSPI, New York, NY, USA. [9]Partners HealthCare, Cambridge, MA, USA. [10]Center for Genetic Medicine, Northwestern University, Chicago, IL, USA. [11]Broad Institute, Massachusetts, MA, USA. [12]Department of Molecular and Human Genetics, Baylor College of Medicine, One Baylor Plaza, Houston, TX, USA. [13]Department of Cardiovascular Medicine, Mayo Clinic, Rochester, MN, USA. [14]Vanderbilt Institute for Clinical & Translational Research, Vanderbilt University Medical Center, Nashville, TN, USA. [15]Department of Biomedical Informatics, Vanderbilt University Medical Center, Nashville, TN, USA. [16]Research Information Science and Computing, Partners Healthcare, Somerville, MA, USA. [17]Department of Health Sciences Research, Mayo Clinic, Rochester, MN, USA. [18]Brigham and Women's Hospital, Boston, MA, USA. [19]Geisinger, Hood Center for Health Research, Danville, PA, USA. [20]Hasso Plattner Institute for Digital Health at Mount Sinai, Icahn School of Medicine at Mount Sinai, New York, NY, USA. [21]Partners HealthCare Personalized Medicine, Cambridge, MA, USA. [22]Massachusetts General Hospital, Boston, MA, USA. [23]Marshfield Clinic Research Institute, Marshfield, WI, USA. [24]Broad Institute of MIT and Harvard, Massachusetts, MA, USA. [25]Mayo Clinic, Rochester, MN, USA. [26]Google Inc, Mountain View, CA, USA. [27]Center for Genetic Medicine, Feinberg School of Medicine, Northwestern University, Department of Cardiology, The Louis A Simpson and Kimberly K Querrey Biomedical Research Center Room 5-408, Chicago, IL, USA. [28]Research Information Science and Computing (RISC), Partners Healthcare, Somerville, MA, USA. [29]Baylor College of Medicine, One Baylor Plaza, Houston, USA. [30]Center for Genetic Medicine, Feinberg School of Medicine, Northwestern University, Chicago, IL, USA. [31]Division of Genetics, Department of Medicine, Brigham and Women's Hospital, Department of Medicine, Harvard Medical School, Boston, MA, USA. [32]Departments of Pediatrics and Medicine, Columbia University, New York, NY, USA. [33]Schools of Medicine, Public Health, and Nursing, Johns Hopkins University, Baltimore, MD, USA. [34]Center for Autoimmune Genomics and Etiology, Cincinnati Children's Hospital Medical Center (CCHMC), Cincinnati, OH, USA. [35]Department of Medicine, University of Washington, Seattle, WA, USA. [36]Columbia University Irving Medical Center, New York, NY, USA. [37]Department of Biomedical Informatics and Medical Education, University of Washington, Seattle, WA, USA. [38]National Human Genome Research Institute, Maryland, MD, USA. [39]Departments of Biomedical Informatics and Medicine, Vanderbilt University, Nashville, TN, USA. [40]Division of Epidemiology, Department of Medicine, Vanderbilt Genetics Institute, Vanderbilt University Medical Center, Nashville, TN, USA. [41]Department of Biomedical Informatics, Columbia University, New York, NY, USA. [42]Irving Institute for Clinical and Translational Research, Columbia University, New York, NY, USA. [43]Department of Medicine, Division of Clinical Pharmacology, Vanderbilt University Medical Center, Nashville, TN, USA. [44]Department of Health Sciences Research, Mayo Clinic, Center for Individualized Medicine, Mayo Clinic, Rochester, MN, USA. [45]Department of Bioethics & Humanties, University of Washington, Seattle, WA, USA. [46]Harvard Medical School, Boston, MA, USA. [47]Partners HealthCare, Somerville, MA, USA. [48]Department of Medicine, Division of Nephrology, Columbia University Vagelos College of Physicians and Surgeons, New York, NY, USA. [49]Vanderbilt University Medical Center, Department of Medicine, Nashville, TN, USA. [50]Center for Applied Genomics Children's Hospital of Philadelphia, Division of Human Genetics Children's Hospital of Philadelphia, Department of Pediatrics Perelman School of Medicine University of Pennsylvania, Philadelphia, PA, USA. [51]Geisinger Medical Center, Danville, PA, USA. [52]Department of Medicine (Medical Genetics), University of Washington School of Medicine, Seattle, WA, USA. [53]Center for Genetic Medicine, Northwestern University Feinberg School of Medicine, Chicago, IL, USA. [54]Laboratory for Molecular Medicine, Partners Healthcare Personalized Medicine, Cambridge, MA, USA. [55]Brigham and Women's Hospital, Broad Institute, Harvard Medical School, EC Alumnae Building, Boston, MA, USA. [56]Center for Applied Genomics Children's Hospital of Philadelphia, Division of Human Genetics Children's Hospital of Philadelphia, Philadelphia, PA, USA. [57]Center for Applied Genomics Children's Hospital of Philadelphia, Divisions of Human Genetics and Pulmonary

Medicine Children's Hospital of Philadelphia, Department of Pediatrics Perelman School of Medicine University of Pennsylvania, Philadelphia, PA, USA. [58]Cincinnati Children's Hospital Medical Center, University of Cincinnati College of Medicine, US Department of Veterans Affairs Medical Center, Cincinnati, Cincinnati, OH, USA. [59]Center for Applied Genomics Children's Hospital of Philadelphia, Philadelphia, PA, USA. [60]Department of Biomedical Informatics and Medical Education, University of Washignton School of Medicine, KP Washington Health Research Institute, Seattle, WA, USA. [61]Division of Endocrinology, Metabolism, and Molecular Medicine, Department of Medicine, Northwestern University Feinberg School of Medicine, Center for Genetic Medicine, Northwestern University Feinberg School of Medicine, Department of Anthropology, Northwestern University, Chicago, IL, USA. [62]Center for Precision Medicine Research, Marshfield Clinic Research Institute, Marshfield, WI, USA. [63]KP Washington Health Research Institute, Univ of Washington School of Public Health, Dept of Health Services, Seattle, WA, USA. [64]Cincinnati Children's Hospital Medical Center (CCHMC), University of Cincinnati College of Medicine, Cincinnati, OH, USA. [65]Division of Genetics and Genomics, Boston Children's Hospital, Department of Pediatrics, Harvard Medical School, Boston, MA, USA. [66]Medical Informatics Services, NewYork-Presbyterian Hospital, New York, NY, USA. [67]Department of Genome Sciences, University of Washington School of Medicine, Seattle, WA, USA. [68]Department of Medicine, Northwestern University Feinberg School of Medicine, Chicago, IL, USA. [69]Biomedical and Translational Informatics, Geisinger, Fremont, CA, USA. [70]Biomedical and Translational Informatics, Geisinger, Danville, PA, USA. [71]Brigham & Women's Hospital, Harvard Medical School, Boston, MA, USA. [72]Children's Hospital of Philadelphia, Department of Surgery, University of Pennsylvania, Department of Surgery, University of Pennsylvania, Philadelphia, PA, USA. [73]Geisinger, Danville, PA, USA. [74]Center for Genomic Health, Icahn School of Medicine at Mount Sinai, The Charles Bronfman Institute of Personalized Medicine, Icahn School of Medicine at Mount Sinai, Departments of Genetics and Medicine, Icahn School of Medicine at Mount Sinai, New York, NY, USA. [75]Columbia University, New York, NY, USA. [76]Laboratory for Molecular Medicine, Partners Healthcare Personalized Medicine, Brigham and Women's Hospital, Cambridge, MA, USA. [77]Department of Health Sciences Research, Mayo Clinic, Rochester, MN, USA. [78]Department of Cardiovascular Medicine, Mayo Clinic, Rochester, MN, USA. [79]Department of Pediatrics, University of Cincinnati college of Medicine, University of Cincinnati, Center of Autoimmune Genomics and Etiology, Division of Allergy & Immunology, Cincinnati Children's Hospital Medical Center, Cincinnati, OH, USA. [80]Geisinger Genomic Medicine Institute, Danville, PA, USA. [81]Department of Cardiovascular Medicine, Mayo Clinic, Rochester, MN, USA. [82]Meharry Medical College, Baptist Cancer Center, Memphis, TN, USA. [83]Kaiser Permanente Washington Health Research Institute, Seattle, WA, USA. [84]Partners Healthcare Personalized Medicine, Brigham and Women's Hospital, Harvard Medical School, Cambridge, MA, USA. [85]Baylor College of Medicine, One Baylor Plaza, Houston, TX, USA. [86]Geisinger, Danville, PA, USA. [87]Genetic Services Kaiser Permanente of Washington, Seattle, WA, USA. [88]Cincinnati Children's Hospital Medical Center, Cincinnati, OH, USA. [89]National Human Genome Research Institute, National Institutes of Health, Bethesda, MD, USA. [90]University of Alabama at Birmingham, Birmingham, AL, USA. [91]Partners Healthcare Personalized Medicine, Harvard Medical School, Mountain View, CA, USA. [92]Mayo Clinic, Scottsdale, AZ, USA. [93]Cincinnati Children's Hospital Medical Center, Cincinnati, OH, USA. [94]Department of Biomedical Informatics, Columbia University Medical Center, Columbia University, New York, NY, USA. [95]Department of Communication, University of Cincinnati, Cincinnati, OH, USA. [96]Broad Institute of MIT & Harvard, Cambridge, MA, USA. [97]Broad Institute of MIT & Harvard, Cambridge, MA, USA. [98]Partners Healthcare Personalized Medicine, Cambridge, MA, USA. [99]Department of Biomedical Informatics, Vanderbilt University, Nashville, TN, USA. [100]Department of Medicine, Division of Nephrology, Columbia University, New York, NY, USA. [101]Department of Population Health Sciences, Duke University School of Medicine, Durham, NC, USA. [102]Ethics Center, Cincinnati Children's Hospital Medical Center, Department of Pediatrics, University of Cincinnati, Cincinnati, OH, USA. [103]Department of Medicine, Division of Nephrology, Columbia University, New York, NY, USA. [104]Department of Internal Medicine, Vanderbilt University Medical Center, Nashville, TN, USA. [105]Massachusetts General Hospital, Partners Healthcare, Harvard Medical School, Somerville, MA, USA. [106]Human Genome Sequencing Center at the Baylor College of Medicine, One Baylor Plaza, Houston, TX, USA. [107]Human Genome Sequencing Center at the Baylor College of Medicine, One Baylor Plaza, Houston, TX, USA. [108]Division of Human Genetics, Cincinnati Children's Hospital Medical Center, College of Medicine, University of Cincinnati, Cincinnati, OH, USA. [109]Department of Pediatrics, University of Cincinnati, Cincinnati, OH, USA. [110]Biomedical Informatics, Cincinnati Children's Hospital Medical Center, Cincinnati, OH, USA. [111]The Charles Bronfman Institute for Personalized Medicine, Icahn School of Medicine at Mount Sinai, New York, NY, USA. [112]Pharmacy Department, Mount Sinai Hospital, New York, NY, USA. [113]Geisinger, Danville, PA, USA. [114]Department of Biomedical Informatics, Vanderbilt University Medical Center, Nashville, TN, USA. [115]Department of Dermatology, Columbia University, New York, NY, USA. [116]Geisinger, Danville, PA, USA. [117]School of Graduate Studies and Research, Meharry Medical College, Nashville, TN, USA. [118]Division of

Human Genetics, Division of Patient Services, Cincinnati Children's Hospital, Cincinnati, OH, USA. [119]Department of Pharmacology, Northwestern University Feinberg School of Medicine, Center for Genetic Medicine, Northwestern University, Chicago, IL, USA. [120]Kaiser Permanente Washington Health Research Institute, University of Washington Department of Biomedical Informatics and Medical Education, Seattle, WA, USA. [121]Vanderbilt University Medical Center Department of Medicine, Nashville, TN, USA. [122]Department of Preventive Medicine Northwestern University Feinberg School of Medicine, Chicago, IL, USA. [123]Department of Preventive Medicine Northwestern University Feinberg School of Medicine, Chicago, IL, USA. [124]Medical and Population Genetics, Broad Institute or MIT and Harvard, Center for Genomic Medicine, Massachusetts General Hospital, Boston, MA, USA. [125]Center for Genomic Medicine, Massachusetts General Hospital, Department of Pathology, Massachusetts General Hospital/Harvard Medical School, Broad Institue Clinical Research Sequencing Platform (CRSP), Simches Research Building, Boston, MA, USA. [126]University of Pennsylvania, Perelman School of Medicine, Philadelphia, PA, USA. [127]Human Genome Sequencing Center Baylor College of Medicine, Houston, TX, USA. [128]University of Texas at Arlington, Human Genome Sequencing Center Baylor College of Medicine, Houston, TX, USA. [129]Vanderbilt University Medical Center, Nashville, TN, USA. [130]Division of Medical Genetics, School of Medicine, University of Washington, Seattle, WA, USA. [131]Center for Applied Genomics, Children's Hospital of Philadelphia, Department of Pathology and Laboratory Medicine, University of Pennsylvania, Philadelphia, PA, USA. [132]Icahn School of Medicine at Mount Sinai, New York, NY, USA. [133]KP Washington Health Research Institute, Seattle, WA, USA. [134]Columbia University, New York, NY, USA. [135]Partners Healthcare, Cambridge, MA, USA. [136]Biomedical Ethics Program, Mayo Clinic, Department of Health Sciences Research, Mayo Clinic, Rochester, MN, USA. [137]Clinical and Translational Research Center, Meharry Medical College, Nashville, TN, USA. [138]Center for Applied Genomics, Children's Hospital of Philadelphia, Department of Pediatrics, Perelman School of Medicine, University of Pennsylvania, Philadelphia, PA, USA. [139]Broad Institute of MIT & Harvard, Cambridge, MA, USA. [140]Department of Biomedical Informatics, Vanderbilt University Medical Center, Center for Patient and Professional Advocacy, Vanderbilt University, Nashville, TN, USA. [141]Northwestern University, Chicago, IL, USA. [142]Department of Internal Medicine, Meharry Medical College, Nashville, TN, USA. [143]Department of Psychiatry and Center for Genomic Medicine, Massachusetts General Hospital, Stanley Center for Psychiatric Research, Simches Research Building, Boston, MA, USA. [144]Mayo Clinic, Rochester, MN, USA. [145]Feinberg School of Medicine, Northwestern University, Chicago, IL, USA. [146]Channing Division of Network Medicine, Brigham and Women's Hospital, Boston, MA, USA. [147]Johns Hopkins University, Geisinger, Baltimore, MD, USA. [148]Biomedical Ethics Unit, Social Studies of Medicine, Faculty of Medicine, McGill University, Montreal, QC, Canada. [149]Department of Pediatrics, Vanderbilt University Medical Center, Nashville, TN, USA. [150]Department of Medicine, Vanderbilt University Medical Center, Nashville, TN, USA. [151]The University of Texas at San Antonio, San Antonio, TX, USA. [152]Massachusetts General Hospital, Partners HealthCare, Cambridge, MA, USA. [153]Comparative Health Outcomes, Policy & Economics (CHOICE) Institute, Department of Pharmacy, University of Washington, Seattle, WA, USA. [154]Division of Nephrology Department of Medicine, Columbia University, New York, NY, USA. [155]Broad Institute, Cambridge, MA, USA. [156]Division of Biomedical Informatics, Cincinnati Children's Hospital Medical Center, Cincinnati, OH, USA. [157]Human Genome Sequencing Center, Baylor College of Medicine, Baylor College of Medicine, One Baylor Plaza, Houston, TX, USA. [158]Northwestern University, Chicago, IL, USA. [159]Human Genome Sequencing Center at the Baylor College of Medicine, One Baylor Plaza, Houston, TX, USA. [160]Human Genome Sequencing Center, Baylor College of Medicine, One Baylor Plaza, Houston, TX, USA. [161]Channing Division of Network Medicine, Brigham and Women's Hospital, Department of Medicine, Harvard Medical School, Boston, MA, USA. [162]Department of Medicine, Division of Cardiovascular Medicine, Vanderbilt University Medical Center, Nashville, TN, USA. [163]Columbia University, New York, NY, USA. [164]Department of Pediatrics, Cincinnati Children's Hospital Medical Center, Department of Biomedical Informatics, University of Cincinnati College of Medicine, Cincinnati, OH, USA. [165]Department of Medicine, Vanderbilt Ingram Cancer Center, Vanderbilt University Medical Center, Nashville, TN, USA. [166]Geisinger, Danville, PA, USA. [167]Geisinger, Danville, PA, USA. [168]Laboratory for Molecular Medicine, Partners Healthcare Personalized Medicine, Department of Pathology, Massachusetts General Hospital/Harvard Medical School, Cambridge, MA, USA. [169]Departments of Pediatrics, Columbia University, New York, NY, USA. [170]Department of Molecular and Genetics, Baylor College of Medicine, Houston, TX, USA. [171]Division of Human Genetics, Center for Prevention of Preterm Birth, Perinatal Institute and March of Dimes Prematurity Research Center Ohio Collaborative, Cincinnati Children's Hospital Medical Center, Cincinnati, Cincinnati, OH, USA. [172]Department of Pediatrics, University of Cincinnati College of Medicine, Cincinnati, OH, USA. [173]Laboratory for Molecular Medicine, Partners Healthcare Personalized Medicine, Department of Pathology, Massachusetts General Hospital/Harvard Medical School, Cambridge, MA, USA.

**Membership of the International Consortium of Blood Pressure**

Authors and contributors associated with the 2018 Nature Genetics publication: "Genetic analysis of over one million people identifies 535 new loci associated with blood pressure traits".

Evangelos Evangelou[1,2], Helen R Warren[3,4], He Gao[1,5], Georgios Ntritsos[2], Niki Dimou[2], Tonu Esko[16,17], Reedik Mägi[16], Lili Milani[16], Peter Almgren[18], Thibaud Boutin[19], Stéphanie Debette[20,21], Jun Ding[22], Franco Giulianini[23], Elizabeth G Holliday[24], Anne U Jackson[25], Ruifang Li-Gao[26], Wei-Yu Lin[27], Jian'an Luan[28], Massimo Mangino[29,30], Christopher Oldmeadow[24], Bram Peter Prins[31], Yong Qian[22], Muralidharan Sargurupremraj[21], Nabi Shah[32,33], Praveen Surendran[27], Sébastien Thériault[34,35], Niek Verweij[17,36,37], Sara M Willems[28], Jing-Hua Zhao[28], Philippe Amouyel[38], John Connell[39], Renée de Mutsert[26], Alex SF Doney[32], Martin Farrall[40,41], Cristina Menni[29], Andrew D Morris[42], Raymond Noordam[43], Guillaume Paré[34], Neil R Poulter[44], Denis C Shields[45], Alice Stanton[46], Simon Thom[47], Gonçalo Abecasis[48], Najaf Amin[49], Dan E Arking[50], Kristin L Ayers[51,52], Caterina M Barbieri[53], Chiara Batini[54], Joshua C Bis[55], Tineka Blake[54], Murielle Bochud[56], Michael Boehnke[25], Eric Boerwinkle[57], Dorret I Boomsma[58], Erwin P Bottinger[59], Peter S Braund[60,61], Marco Brumat[62], Archie Campbell[63,64], Harry Campbell[65], Aravinda Chakravarti[50], John C Chambers[1,5,66-68], Ganesh Chauhan[69], Marina Ciullo[70,71], Massimiliano Cocca[72], Francis Collins[73], Heather J Cordell[51], Gail Davies[74,75], Martin H de Borst[76], Eco J de Geus[58], Ian J Deary[74,75], Joris Deelen[77], Fabiola Del Greco M[78], Cumhur Yusuf Demirkale[79], Marcus Dörr[80,81], Georg B Ehret[50,82], Roberto Elosua[83,84], Stefan Enroth[85], A Mesut Erzurumluoglu[54], Teresa Ferreira[86,87], Mattias Frånberg[88-90], Oscar H Franco[91], Ilaria Gandin[62], Paolo Gasparini[62,72], Vilmantas Giedraitis[92], Christian Gieger[93-95], Giorgia Girotto[62,72], Anuj Goel[40,41], Alan J Gow[74,96], Vilmundur Gudnason[97,98], Xiuqing Guo[99], Ulf Gyllensten[85], Anders Hamsten[88,89], Tamara B Harris[100], Sarah E Harris[63,74], Catharina A Hartman[101], Aki S Havulinna[102,103], Andrew A Hicks[78], Edith Hofer[104,105], Albert Hofman[91,106], Jouke-Jan Hottenga[58], Jennifer E Huffman[19,107,108], Shih-Jen Hwang[107,108], Erik Ingelsson[109,110], Alan James[111,112], Rick Jansen[113], Marjo-Riitta Jarvelin[1,5,114-116], Roby Joehanes[107,117], Åsa Johansson[85], Andrew D Johnson[107,118], Peter K Joshi[65], Pekka Jousilahti[102], J Wouter Jukema[119], Antti Jula[102], Mika Kähönen[120,121], Sekar Kathiresan[17,36,122], Bernard D Keavney[123,124], Kay-Tee Khaw[125], Paul Knekt[102], Joanne Knight[126], Ivana Kolcic[127], Jaspal S Kooner[5,67,68,128], Seppo Koskinen[102], Kati Kristiansson[102], Zoltan Kutalik[56,129], Maris Laan[130], Marty Larson[107], Lenore J Launer[100], Benjamin Lehne[1], Terho Lehtimäki[131,132], David CM Liewald[74,75], Li Lin[82], Lars Lind[133], Cecilia M Lindgren[40,87,134], YongMei Liu[135], Ruth JF Loos[28,59,136], Lorna M Lopez[74,137,138], Yingchang Lu[59], Leo-Pekka Lyytikäinen[131,132], Anubha Mahajan[40], Chrysovalanto Mamasoula[139], Jaume Marrugat[83], Jonathan Marten[19], Yuri Milaneschi[140], Anna Morgan[62], Andrew P Morris[40,141], Alanna C Morrison[142], Peter J Munson[79], Mike A Nalls[143,144], Priyanka Nandakumar[50], Christopher P Nelson[60,61], Teemu Niiranen[102,145], Ilja M Nolte[146], Teresa Nutile[70], Albertine J Oldehinkel[147], Ben A Oostra[49], Paul F O'Reilly[148], Elin Org[16], Sandosh Padmanabhan[64,149], Walter Palmas[150], Aarno Palotie[103,151,152], Alison Pattie[75], Brenda WJH Penninx[140], Markus Perola[102,103,153], Annette Peters[94,95,154], Ozren Polasek[127,155], Peter P Pramstaller[78,156,157], Quang Tri Nguyen[79], Olli T Raitakari[158,159], Rainer Rettig[161], Kenneth Rice[162], Paul M Ridker[23,163], Janina S Ried[94], Harriëtte Riese[147], Samuli Ripatti[103,164], Antonietta Robino[72], Lynda M Rose[23], Jerome I Rotter[99], Igor Rudan[165], Daniela Ruggiero[70,71], Yasaman Saba[166], Cinzia F Sala[53], Veikko Salomaa[102], Nilesh J Samani[60,61], Antti-Pekka Sarin[103], Reinhold

Schmidt[104], Helena Schmidt[166], Nick Shrine[54], David Siscovick[167], Albert V Smith[97,98], Harold Snieder[146], Siim Sõber[130], Rossella Sorice[70], John M Starr[74,168], David J Stott[169], David P Strachan[170], Rona J Strawbridge[88,89], Johan Sundström[133], Morris A Swertz[171], Kent D Taylor[99], Alexander Teumer[81,172], Martin D Tobin[54], Maciej Tomaszewski[123,124], Daniela Toniolo[53], Michela Traglia[53], Stella Trompet[119,173], Jaakko Tuomilehto[174-177], Christophe Tzourio[21], André G Uitterlinden[91,178], Ahmad Vaez[146,179], Peter J van der Most[146], Cornelia M van Duijn[49], Germaine C Verwoert[91], Veronique Vitart[19], Uwe Völker[81,180], Peter Vollenweider[181], Dragana Vuckovic[62,182], Hugh Watkins[40,41], Sarah H Wild[183], Gonneke Willemsen[58], James F Wilson[19,65], Alan F Wright[19], Jie Yao[99], Tatijana Zemunik[184], Weihua Zhang[1,67], John R Attia[24], Adam S Butterworth[27,185], Daniel I Chasman[23,163], David Conen[186,187], Francesco Cucca[188,189], John Danesh[27,185], Caroline Hayward[19], Joanna MM Howson[27], Markku Laakso[190], Edward G Lakatta[191], Claudia Langenberg[28], Olle Melander[18], Dennis O Mook-Kanamori[26,192], Colin NA Palmer[32], Lorenz Risch[193-195], Robert A Scott[28], Rodney J Scott[24], Peter Sever[128], Tim D Spector[29], Pim van der Harst[196], Nicholas J Wareham[28], Eleftheria Zeggini[31], Daniel Levy[107,118], Patricia B Munroe[3,4], Christopher Newton-Cheh[134,197,198], Morris J Brown[3,4], Andres Metspalu[16], Bruce M. Psaty[201,202], Louise V Wain[54], Paul Elliott[1,5,203-205], Mark J Caulfield[3,4]

[1]Department of Epidemiology and Biostatistics, Imperial College London, London, UK. [2]Department of Hygiene and Epidemiology, University of Ioannina Medical School, Ioannina, Greece. [3]William Harvey Research Institute, Barts and The London School of Medicine and Dentistry, Queen Mary University of London, London, UK. [4]National Institute for Health Research, Barts Cardiovascular Biomedical Research Center, Queen Mary University of London, London, UK. [5]MRC-PHE Centre for Environment and Health, Imperial College London, London, UK. [7]Division of Epidemiology, Department of Medicine, Institute for Medicine and Public Health, Vanderbilt Genetics Institute, Vanderbilt University Medical Center, Tennessee Valley Healthcare System (626)/Vanderbilt University, Nashville, TN, USA. [8]Vanderbilt Genetics Institute, Vanderbilt Epidemiology Center, Department of Obstetrics and Gynecology, Vanderbilt University Medical Center; Tennessee Valley Health Systems VA, Nashville, TN, USA. [9]Department of Epidemiology, Emory University Rollins School of Public Health, Atlanta, GA, USA. [10]Department of Biomedical Informatics, Emory University School of Medicine, Atlanta, GA, USA. [11]Massachusetts Veterans Epidemiology Research and Information Center (MAVERIC), VA Boston Healthcare System, Boston, USA. [12]Division of Aging, Department of Medicine, Brigham and Women's Hospital, Boston, MA, Department of Medicine, Harvard Medical School, Boston, MA, USA. [13]Atlanta VAMC and Emory Clinical Cardiovascular Research Institute, Atlanta, GA, USA. [14]VA Palo Alto Health Care System; Division of Cardiovascular Medicine, Stanford University School of Medicine, CA, USA. [15]Nephrology Section, Memphis VA Medical Center and University of Tennessee Health Science Center, Memphis, TN, USA. [16]Estonian Genome Center, University of Tartu, Tartu, Estonia. [17]Program in Medical and Population Genetics, Broad Institute of Harvard and MIT, Cambridge, MA, USA. [18]Department Clinical Sciences, Malmö, Lund University, Malmö, Sweden. [19]MRC Human Genetics Unit, MRC Institute of Genetics and Molecular Medicine, University of Edinburgh, Western General Hospital, Edinburgh, Scotland, UK. [20]Department of Neurology, Bordeaux University Hospital, Bordeaux, France. [21]Univ. Bordeaux, Inserm, Bordeaux Population Health Research Center, CHU Bordeaux, Bordeaux, France. [22]Laboratory of Genetics and Genomics, NIA/NIH, Baltimore, MD, USA. [23]Division of Preventive Medicine, Brigham and Women's Hospital, Boston, MA, USA. [24]Hunter Medical Reseach Institute and Faculty of Health, University of Newcastle, New Lambton Heights, New South Wales, Australia. [25]Department of Biostatistics and Center for Statistical Genetics, University of Michigan, Ann Arbor, MI, USA. [26]Department of Clinical Epidemiology, Leiden University Medical Center, Leiden, the Netherlands. [27]MRC/BHF Cardiovascular Epidemiology Unit, Department of Public Health and Primary Care, University of Cambridge, Cambridge, UK. [28]MRC Epidemiology Unit, University of Cambridge School of Clinical Medicine, Cambridge, UK. [29]Department of Twin Research and Genetic Epidemiology, Kings College London, London, UK. [30]NIHR Biomedical Research Centre at Guy's and St Thomas' Foundation Trust, London, UK. [31]Wellcome Trust Sanger Institute, Hinxton, UK. [32]Division of Molecular and Clinical Medicine, School of Medicine, University of Dundee, UK. [33]Department of Pharmacy, COMSATS Institute of Information Technology, Abbottabad, Pakistan. [34]Department of Pathology and Molecular Medicine, McMaster University, Hamilton, Canada. [35]Institut universitaire de cardiologie et de pneumologie de Québec-Université Laval,

Quebec City, Canada. [36]Cardiovascular Research Center and Center for Human Genetic Research, Massachusetts General Hospital, Boston, Massachusetts, MA, USA. [37]University of Groningen, University Medical Center Groningen, Department of Cardiology, Groningen, The Netherlands. [38]University of Lille, Inserm, Centre Hosp. Univ Lille, Institut Pasteur de Lille, UMR1167 - RID-AGE - Risk factors and molecular determinants of aging-related diseases, Epidemiology and Public Health Department, Lille, France. [39]University of Dundee, Ninewells Hospital & Medical School, Dundee, UK. [40]Wellcome Trust Centre for Human Genetics, University of Oxford, Oxford, UK. [41]Division of Cardiovascular Medicine, Radcliffe Department of Medicine, University of Oxford, Oxford, UK. [42]Usher Institute of Population Health Sciences and Informatics, University of Edinburgh, UK. [43]Department of Internal Medicine, Section Gerontology and Geriatrics, Leiden University Medical Center, Leiden, The Netherlands. [44]Imperial Clinical Trials Unit, Stadium House, 68 Wood Lane, London, UK. [45]School of Medicine, University College Dublin, Ireland. [46]Molecular and Cellular Therapeutics, Royal College of Surgeons in Ireland, Dublin, Ireland. [47]International Centre for Circulatory Health, Imperial College London, London, UK. [48]Center for Statistical Genetics, Dept. of Biostatistics, SPH II, Washington Heights, Ann Arbor, MI, USA. [49]Genetic Epidemiology Unit, Department of Epidemiology, Erasmus MC, Rotterdam, the Netherlands. [50]Center for Complex Disease Genomics, McKusick-Nathans Institute of Genetic Medicine, Johns Hopkins University School of Medicine, Baltimore, MD, USA. [51]Institute of Genetic Medicine, Newcastle University, Newcastle upon Tyne, UK. [52]Sema4, a Mount Sinai venture, Stamford, CT, USA. [53]Division of Genetics and Cell Biology, San Raffaele Scientific Institute, Milano, Italy. [54]Department of Health Sciences, University of Leicester, Leicester, UK. [55]Cardiovascular Health Research Unit, Department of Medicine, University of Washington, Seattle, WA, USA. [56]Institute of Social and Preventive Medicine, University Hospital of Lausanne, Lausanne, Switzerland. [57]Human Genetics Center, School of Public Health, The University of Texas Health Science Center at Houston and Human Genome Sequencing Center, Baylor College of Medicine, One Baylor Plaza, Houston, TX, USA. [58]Department of Biological Psychology, Vrije Universiteit Amsterdam, EMGO+ institute, VU University medical center, Amsterdam, the Netherlands. [59]The Charles Bronfman Institute for Personalized Medicine, Icachn School of Medicine at Mount Sinai, NY, USA. [60]Department of Cardiovascular Sciences, University of Leicester, Leicester, UK. [61]NIHR Leicester Biomedical Research Centre, Glenfield Hospital, Groby Road, Leicester, UK. [62]Department of Medical, Surgical and Health Sciences, University of Trieste, Trieste, Italy. [63]Medical Genetics Section, Centre for Genomic and Experimental Medicine, Institute of Genetics and Molecular Medicine, University of Edinburgh, Edinburgh, UK. [64]Generation Scotland, Centre for Genomic and Experimental Medicine, University of Edinburgh, Edinburgh, UK. [65]Centre for Global Health Research, Usher Institute of Population Health Sciences and Informatics, University of Edinburgh, Edinburgh, Scotland, UK. [66]Lee Kong Chian School of Medicine, Nanyang Technological University, Singapore, Singapore. [67]Department of Cardiology, Ealing Hospital, Middlesex, UK. [68]Imperial College Healthcare NHS Trust, London, UK. [69]Centre for Brain Research, Indian Institute of Science, Bangalore, India. [70]Institute of Genetics and Biophysics "A. Buzzati-Traverso", CNR, Napoli, Italy. [71]IRCCS Neuromed, Pozzilli, Isernia, Italy. [72]Institute for Maternal and Child Health IRCCS Burlo Garofolo, Trieste, Italy. [73]Medical Genomics and Metabolic Genetics Branch, National Human Genome Research Institute, NIH, Bethesda, MD, USA. [74]Centre for Cognitive Ageing and Cognitive Epidemiology, University of Edinburgh, 7 George Square, Edinburgh, UK. [75]Department of Psychology, University of Edinburgh, 7 George Square, Edinburgh, UK. [76]Department of Internal Medicine, Division of Nephrology, University of Groningen, University Medical Center Groningen, Groningen, The Netherlands. [77]Department of Molecular Epidemiology, Leiden University Medical Center, Leiden, the Netherlands. [78]Institute for Biomedicine, Eurac Research, Bolzano, Italy - Affiliated Institute of the University of Lübeck, Lübeck, Germany. [79]Mathematical and Statistical Computing Laboratory, Office of Intramural Research, Center for Information Technology, National Institutes of Health, Bethesda, MD, USA. [80]Department of Internal Medicine B, University Medicine Greifswald, Greifswald, Germany. [81]DZHK (German Centre for Cardiovascular Research), partner site Greifswald, Greifswald, Germany. [82]Cardiology, Department of Medicine, Geneva University Hospital, Geneva, Switzerland. [83]CIBERCV & Cardiovascular Epidemiology and Genetics, IMIM. Dr Aiguader 88, Barcelona, Spain. [84]Faculty of Medicine, Universitat de Vic-Central de Catalunya, Vic, Spain. [85]Department of Immunology, Genetics and Pathology, Uppsala Universitet, Science for Life Laboratory, Uppsala, Sweden. [86]Wellcome Centre for Human Genetics, University of Oxford, Roosevelt Drive, Oxford, UK. [87]Big Data Institute, Li Ka Shing Center for Health for Health Information and Discovery, Oxford University, Old Road, Oxford, UK. [88]Cardiovascular Medicine Unit, Department of Medicine Solna, Karolinska Institutet, Stockholm, Sweden. [89]Centre for Molecular Medicine, L8:03, Karolinska Universitetsjukhuset, Solna, Sweden. [90]Department of Numerical Analysis and Computer Science, Stockholm University, Stockholm, Sweden. [91]Department of Epidemiology, Erasmus MC, Rotterdam, the Netherlands. [92]Department of Public Health and Caring Sciences, Geriatrics, Uppsala, Sweden. [93]Research Unit of Molecular Epidemiology, Helmholtz Zentrum München, German Research Center for Environmental Health, Neuherberg,

Germany. [94]Institute of Epidemiology, Helmholtz Zentrum München, German Research Center for Environmental Health, Neuherberg, Germany. [95]German Center for Diabetes Research (DZD e.V.), Neuherberg, Germany. [96]Department of Psychology, School of Social Sciences, Heriot-Watt University, Edinburgh, UK. [97]Faculty of Medicine, University of Iceland, Reykjavik, Iceland. [98]Icelandic Heart Association, Kopavogur, Iceland. [99]The Institute for Translational Genomics and Population Sciences, Department of Pediatrics, LABioMed at Harbor-UCLA Medical Center, Torrance, CA, USA. [100]Intramural Research Program, Laboratory of Epidemiology, Demography, and Biometry, National Institute on Aging, Bethesda, MD, USA. [101]Department of Psychiatry, University of Groningen, University Medical Center Groningen, Groningen, The Netherlands. [102]Department of Public Health Solutions, National Institute for Health and Welfare (THL), Helsinki, Finland. [103]Institute for Molecular Medicine Finland (FIMM), University of Helsinki, Helsinki, Finland. [104]Clinical Division of Neurogeriatrics, Department of Neurology, Medical University of Graz, Graz, Austria. [105]Institute for Medical Informatics, Statistics and Documentation, Medical University of Graz, Graz, Austria. [106]Department of Epidemiology, Harvard T.H. Chan School of Public Health, Boston, MA, USA. [107]National Heart, Lung and Blood Institute's Framingham Heart Study, Framingham, MA, USA. [108]The Population Science Branch, Division of Intramural Research, National Heart Lung and Blood Institute national Institute of Health, Bethesda, MD, USA. [109]Department of Medical Sciences, Molecular Epidemiology and Science for Life Laboratory, Uppsala University, Uppsala, Sweden. [110]Division of Cardiovascular Medicine, Department of Medicine, Stanford University School of Medicine, Stanford, CA, USA. [111]Department of Pulmonary Physiology and Sleep, Sir Charles Gairdner Hospital, Hospital Avenue, Nedlands, Australia. [112]School of Medicine and Pharmacology, University of Western Australia, Australia. [113]Department of Psychiatry, VU University Medical Center, Amsterdam Neuroscience, Amsterdam, the Netherlands. [114]Biocenter Oulu, University of Oulu, Oulu, Finland. [115]Center For Life-course Health Research, University of Oulu, Oulu, Finland. [116]Unit of Primary Care, Oulu University Hospital, Oulu, Finland. [117]Hebrew SeniorLife, Harvard Medical School, Boston, MA, USA. [118]Population Sciences Branch, National Heart, Lung and Blood Institute, National Institutes of Health, Bethesda, MD, USA. [119]Department of Cardiology, Leiden University Medical Center, Leiden, the Netherlands. [120]Department of Clinical Physiology, Tampere University Hospital, Tampere, Finland. [121]Department of Clinical Physiology, Finnish Cardiovascular Research Center - Tampere, Faculty of Medicine and Life Sciences, University of Tampere, Tampere, Finland. [122]Broad Institute of the Massachusetts Institute of Technology and Harvard University, Cambridge, MA, USA. [123]Division of Cardiovascular Sciences, Faculty of Biology, Medicine and Health, The University of Manchester, Manchester, UK. [124]Division of Medicine, Manchester University NHS Foundation Trust, Manchester Academic Health Science Centre, Manchester, UK. [125]Department of Public Health and Primary Care, Institute of Public Health, University of Cambridge, Cambridge, UK. [126]Data Science Institute and Lancaster Medical School, Lancaster, UK. [127]Department of Public Health, Faculty of Medicine, University of Split, Croatia. [128]National Heart and Lung Institute, Imperial College London, London, UK. [129]Swiss Institute of Bioinformatics, Lausanne, Switzerland. [130]Institute of Biomedicine and Translational Medicine, University of Tartu, Tartu, Estonia. [131]Department of Clinical Chemistry, Fimlab Laboratories, Tampere, Finland. [132]Department of Clinical Chemistry, Finnish Cardiovascular Research Center - Tampere, Faculty of Medicine and Life Sciences, University of Tampere, Tampere, Finland. [133]Department of Medical Sciences, Cardiovascular Epidemiology, Uppsala University, Uppsala, Sweden. [134]Program in Medical and Population Genetics, Broad Institute, Cambridge, MA, USA. [135]Division of Public Health Sciences, Wake Forest School of Medicine, Winston-Salem, NC, USA. [136]Mindich Child health Development Institute, The Icahn School of Medicine at Mount Sinai, New York, NY, USA. [137]Department of Psychiatry, Royal College of Surgeons in Ireland, Education and Research Centre, Beaumont Hospital, Dublin, Ireland. [138]University College Dublin, UCD Conway Institute, Centre for Proteome Research, UCD, Belfield, Dublin, Ireland. [139]Institute of Health and Society, Newcastle University, Newcastle upon Tyne, UK. [140]Department of Psychiatry, Amsterdam Public Health and Amsterdam Neuroscience, VU University Medical Center/GGZ inGeest, Amsterdam, The Netherlands. [141]Department of Biostatistics, University of Liverpool, Block F, Waterhouse Building, Liverpool, UK. [142]Department of Epidemiology, Human Genetics and Environmental Sciences, School of Public Health, University of Texas Health Science Center at Houston, Houston, TX, USA. [143]Data Tecnica International, Glen Echo, MD, USA. [144]Laboratory of Neurogenetics, National Institute on Aging, Bethesda, USA. [145]Department of Medicine, Turku University Hospital and University of Turku, Finland. [146]Department of Epidemiology, University of Groningen, University Medical Center Groningen, Groningen, The Netherlands. [147]Interdisciplinary Center Psychopathology and Emotion regulation (ICPE), University of Groningen, University Medical Center Groningen, Groningen, The Netherlands. [148]SGDP Centre, Institute of Psychiatry, Psychology and Neuroscience, King's College London, London, UK. [149]British Heart Foundation Glasgow Cardiovascular Research Centre, Institute of Cardiovascular and Medical Sciences, College of Medical, Veterinary and Life Sciences, University of Glasgow, Glasgow, UK. [150]Department of Medicine, Columbia University Medical Center, New York, NY, USA. [151]Analytic and

Translational Genetics Unit, Department of Medicine, Department of Neurology and Department of Psychiatry Massachusetts General Hospital, Boston, MA, USA. [152]The Stanley Center for Psychiatric Research and Program in Medical and Population Genetics, The Broad Institute of MIT and Harvard, Cambridge, MA, USA. [153]University of Tartu, Tartu, Estonia. [154]German Center for Cardiovascular Disease Research (DZHK), partner site Munich, Neuherberg, Germany. [155]Psychiatric hospital “Sveti Ivan”, Zagreb, Croatia. [156]Department of Neurology, General Central Hospital, Bolzano, Italy. [157]Department of Neurology, University of Lübeck, Lübeck, Germany. [158]Department of Clinical Physiology and Nuclear Medicine, Turku University Hospital, Turku, Finland. [159]Research Centre of Applied and Preventive Cardiovascular Medicine, University of Turku, Turku, Finland. [161]Institute of Physiology, University Medicine Greifswald, Karlsburg, Germany. [162]Department of Biostatistics University of Washington, Seattle, WA, USA. [163]Harvard Medical School, Boston, MA, USA. [164]Public health, Faculty of Medicine, University of Helsinki, Finland. [165]Centre for Global Health Research, Usher Institute of Population Health Sciences and Informatics, University of Edinburgh, Scotland, UK. [166]Gottfried Schatz Research Center for Cell Signaling, Metabolism & Aging, Molecular Biology and Biochemistry, Medical University of Graz, Graz, Austria. [167]The New York Academy of Medicine, New York, NY, USA. [168]Alzheimer Scotland Dementia Research Centre, University of Edinburgh, Edinburgh, UK. [169]Institute of Cardiovascular and Medical Sciences, Faculty of Medicine, University of Glasgow, UK. [170]Population Health Research Institute, St George's, University of London, London, UK. [171]Department of Genetics, University of Groningen, University Medical Center Groningen, Groningen, The Netherlands. [172]Institute for Community Medicine, University Medicine Greifswald, Greifswald, Germany. [173]Department of Gerontology and Geriatrics, Leiden University Medical Center, Leiden, the Netherlands. [174]Dasman Diabetes Institute, Dasman, Kuwait. [175]Chronic Disease Prevention Unit, National Institute for Health and Welfare, Helsinki, Finland. [176]Department of Public Health, University of Helsinki, Helsinki, Finland. [177]Saudi Diabetes Research Group, King Abdulaziz University, Jeddah, Saudi Arabia. [178]Department of Internal Medicine, Erasmus MC, Rotterdam, the Netherlands. [179]Research Institute for Primordial Prevention of Non-communicable Disease, Isfahan University of Medical Sciences, Isfahan, Iran. [180]Interfaculty Institute for Genetics and Functional Genomics, University Medicine Greifswald, Greifswald, Germany. [181]Department of Internal Medicine, University Hospital, CHUV, Lausanne, Switzerland. [182]Experimental Genetics Division, Sidra Medical and Research Center, Doha, Qatar. [183]Centre for Population Health Sciences, Usher Institute of Population Health Sciences and Informatics, University of Edinburgh, Scotland, UK. [184]Department of Biology, Faculty of Medicine, University of Split, Croatia. [185]The National Institute for Health Research Blood and Transplant Research Unit in Donor Health and Genomics, University of Cambridge, UK. [186]Division of Cardiology, University Hospital, Basel, Switzerland. [187]Division of Cardiology, Department of Medicine, McMaster University, Hamilton, Canada. [188]Institute of Genetic and Biomedical Research, National Research Council (CNR), Monserrato, Cagliari, Italy. [189]Department of Biomedical Sciences, University of Sassari, Sassari, Italy. [190]Institute of Clinical Medicine, Internal Medicine, University of Eastern Finland and Kuopio University Hospital, Kuopio, Finland. [191]Laboratory of Cardiovascular Science, NIA/NIH, Baltimore, MD, USA. [192]Department of Public Health and Primary Care, Leiden University Medical Center, Leiden, the Netherlands. [193]Labormedizinisches Zentrum Dr. Risch, Schaan, Liechtenstein. [194]Private University of the Principality of Liechtenstin, Triesen, Liechtenstein. [195]University Insitute of Clinical Chemistry, Inselspital, Bern University Hospital, University of Bern, Bern, Switzerland. [196]Department of Cardiology, University of Groningen, University Medical Center Groningen, Groningen, The Netherlands. [197]Center for Genomic Medicine, Massachusetts General Hospital, Boston, MA, USA. [198]Cardiovascular Research Center, Massachusetts General Hospital, Boston, MA, USA. [201]Cardiovascular Health Research Unit, Departments of Medicine, Epidemiology and Health Services, University of Washington, Seattle, WA, USA. [202]Kaiser Permanente Washington Health Research Institute, Seattle, WA, USA. [203]National Institute for Health Research Imperial Biomedical Research Centre, Imperial College Healthcare NHS Trust and Imperial College London, London, UK. [204]UK Dementia Research Institute (UK DRI) at Imperial College London, London, UK. [205]Health Data Research-UK London substantive site, London, UK.

### Membership of the Meta-Analyses of Glucose and Insulin-Related Traits Consortium

#### Authors and contributors associated with the 2023 American Journal of Human Genetics publication: "Loci for insulin processing and secretion provide insight into type 2 diabetes risk".

K Alaine Broadway[1], Xianyong Yin[2,3], Alice Williamson[4,5], Victoria A Parsons[1], Emma P Wilson[1], Anne H Moxley[1], Swarooparani Vadlamudi[1], Arushi Varshney[6], Anne U Jackson[3], Vasudha Ahuja[7], Stefan R Bornstein[8,9,10], Laura J Corbin[11,12], Graciela E Delgado[13], Om P Dwivedi[14,15], Lilian Fernandes Silva[16], Timothy M Frayling[17], Harald Grallert[18,19,10], Stefan Gustafsson[20], Liisa Hakaste[7], Ulf Hammar[21], Christian Herder[10,22,23], Sandra Herrmann[24,9], Kurt Højlund[25], David A Hughes[11,12], Marcus E Kleber[13,26], Cecilia M Lindgren[27,28,29,30], Ching-Ti Liu[31], Jian'an Luan[4], Anni Malmberg[32], Angela P Moissl[33,34,13,] Andrew P Morris[35], Nikolaos Perakakis[8,9,10], Annette Peters[19,10], John R Petrie[36], Michael Roden[22,23,10], Peter EH Schwarz[24,9,10], Sapna Sharma[10,18,19,37], Angela Silveira[38,39], Rona J Strawbridge[40,38], Tiinamaija Tuomi[7,15,41], Andrew R Wood[42], Peitao Wu[31], Björn Zethelius[43], Damiano Baldassarre[44,45], Johan G Eriksson[46,47,48], Tove Fall[21], Jose C Florez[49,50,51], Andreas Fritsche[52,53,10], Bruna Gigante[38], Anders Hamsten[38], Eero Kajantie[54,55,56,57], Markku Laakso[16], Jari Lahti[32], Deborah A Lawlor[11,12], Lars Lind[20], Winfried März[58,13], James B Meigs[59,51,60], Johan Sundström[20], Nicholas J Timpson[11,12], Robert Wagner[52,53,10], Mark Walker[61], Nicholas J Wareham[4,62], Hugh Watkins[63], Inês Barroso[64], Stephen O'Rahilly[65], Niels Grarup[66], Stephen CJ Parker[6,67,2], Michael Boehnke[2,3], Claudia Langenberg[4,68,69], Eleanor Wheeler[4], Karen L Mohlke[1].

[1]Department of Genetics, University of North Carolina, Chapel Hill, NC, USA. [2]Department of Biostatistics, University of Michigan, Ann Arbor, MI, USA. [3]Center for Statistical Genetics, University of Michigan, Ann Arbor, MI, USA. [4]MRC Epidemiology Unit, Institute of Metabolic Science, University of Cambridge School of Clinical Medicine, Cambridge, UK. [5]University of Cambridge Metabolic Research Laboratories, Wellcome Trust-MRC Institute of Metabolic Science, Department of Clinical Biochemistry, University of Cambridge, Cambridge, UK. [6]Department of Computational Medicine and Bioinformatics, University of Michigan, Ann Arbor, MI, USA. [7]Institute for Molecular Medicine Finland, University of Helsinki, Helsinki, Finland. [8]Department of Internal Medicine, Metabolic and Vascular Medicine, Medical Faculty Carl Gustav Carus, Dresden, Germany. [9]Helmholtz Zentrum München, Paul Langerhans Institute Dresden (PLID), University Hospital and Faculty of Medicine, TU Dresden, Dresden, Germany. [10]German Center for Diabetes Research, Neuherberg, Germany. [11]Medical Research Council Integrative Epidemiology Unit (MRC IEU) at the University of Bristol, Bristol, UK. [12]Population Health Sciences, Bristol Medical School, University of Bristol, Bristol, UK. [13]Medical Faculty Mannheim, Heidelberg University, Mannheim, BW, Germany. [14]University of Helsinki, Helsinki, Finland. [15]Folkhälsan Research Center, Helsinki, Finland. [16]Institute of Clinical Medicine, University of Eastern Finland, Kuopio, Finland. [17]College of Medicine and Health, Exeter University, Exeter, UK. [18]Research Unit of Molecular Epidemiology, Helmholtz Zentrum München-German Research Center for Environmental Health, Neuherberg, Germany. [19]Institute of Epidemiology, Helmholtz Zentrum München-German Research Center for Environmental Health, Neuherberg, Germany. [20]Department of Medical Sciences, Clinical Epidemiology, Uppsala University, Uppsala, Sweden. [21]Department of Medical Sciences, Molecular Epidemiology and Science for Life Laboratory, Uppsala University, Uppsala, Sweden. [22]Institute for Clinical Diabetology, German Diabetes Center, Leibniz Center for Diabetes Research at Heinrich Heine University Düsseldorf, Düsseldorf, Germany. [23]Department of Endocrinology and Diabetology, Medical Faculty and University Hospital Düsseldorf, Heinrich Heine University Düsseldorf, Düsseldorf, Germany. [24]Department of Internal Medicine, Prevention and Care of Diabetes, Medical Faculty Carl Gustav Carus, Dresden, Germany. [25]Steno Diabetes Center Odense, Odense, Denmark. [26]SYNLAB MVZ Humangenetik Mannheim, Mannheim, BW, Germany. [27]Oxford Big Data Institute, Li Ka Shing Centre for Health Information and Discovery, University of Oxford, Oxford, UK. [28]Nuffield Department of Population Health, University of Oxford, Oxford, UK. [29]Wellcome Trust Centre Human Genetics, University of Oxford, Oxford, UK. [30]Broad Institute, Cambridge, MA, USA. [31]Department of Biostatistics, Boston University School of Public Health, Boston, MA, USA. [32]Department of Psychology and Logopedics, Faculty of Medicine,

University of Helsinki, Helsinki, Finland. [33]Institute of Nutritional Sciences, Friedrich-Schiller-University, Jena, Germany. [34]Competence Cluster for Nutrition and Cardiovascular Health (nutriCARD), Halle-Jena-Leipzig, Germany. [35]Centre for Genetics and Genomics Versus Arthritis, Centre for Musculoskeletal Research, The University of Manchester, Manchester, UK. [36]School of Health and Wellbeing, University of Glasgow, Glasgow, UK. [37]Chair of Food Chemistry and Molecular Sensory Science, Technische Universität München, Freising, Germany. [38]Department of Medicine Solna, Division of Cardiovascular Medicine, Karolinska Institutet, Stockholm, Sweden. [39]Oxford Biomedical Research Centre, Wellcome Centre for Human Genetics, University of Oxford, Oxford, UK. [40]Institute of Health and Wellbeing, Mental Health and Wellbeing, University of Glasgow, Glasgow, UK. [41]Abdominal Center, Endocrinology, Helsinki University Hospital, Helsinki, Finland. [42]Genetics of Complex Traits, College of Medicine and Health, University of Exeter, Exeter, UK. [43]Department of Geriatrics, Uppsala University, Uppsala, Sweden. [44]Department of Medical Biotechnology and Translational Medicine, Università degli Studi di Milano, Milan, Italy. [45]Cardiovascular Prevention Area, Centro Cardiologico Monzino I.R.C.C.S., Milan, Italy. [46]Department of General Practice and Primary Health Care, Faculty of Medicine, University of Helsinki, Helsinki, Finland. [47]Folkhälsan Research Centre, Helsinki, Finland. [48]Department of Obstetrics and Gynecology, Yong Loo Lin School of Medicine, National University Singapore, Singapore, Singapore. [49]Diabetes Unit and Center for Genomic Medicine, Massachusetts General Hospital, Boston, MA, USA. [50]Programs in Metabolism and Medical & Population Genetics, Broad Institute, Cambridge, MA, USA. [51]Department of Medicine, Harvard Medical School, Boston, MA, USA. [52]Department of Internal Medicine, Diabetology, Tübingen, Germany. [53]Institute for Diabetes Research and Metabolic Diseases, Helmholtz Center Munich, University of Tübingen, Tübingen, Germany, Tübingen, Germany. [54]Population Health Unit, Finnish Institute for Health and Welfare, Helsinki, Finland. [55]PEDEGO Research Unit, MRC Oulu, Oulu University Hospital and University of Oulu, Oulu, Finland. [56]Department of Clinical and Molecular Medicine, Norwegian University of Science and Technology, Trondheim, Norway. [57]Children's Hospital, Helsinki University Hospital and University of Helsinki, Helsinki, Finland. [58]Synlab Academy, SYNLAB Holding Deutschland GmbH, Mannheim, BW, Germany. [59]Department of Medicine, Division of General Internal Medicine, Massachusetts General Hospital, Boston, MA, USA. [60]Program in Medical and Population Genetics, Broad Institute, Cambridge, MA, USA. [61]Faculty of Medical Sciences, Newcastle University, Newcastle upon Tyne, UK. [62]Health Data Research UK, Gibbs Building, London, UK. [63]Division of Cardiovascular Medicine, Radcliffe Department of Medicine, University of Oxford, Oxford, UK. [64]Exeter Centre of Excellence for Diabetes Research (EXCEED), Genetics of Complex Traits, University of Exeter Medical School, University of Exeter, Exeter, UK. [65]MRC Metabolic Diseases Unit, Wellcome Trust-Medical Research Council Institute of Metabolic Science, University of Cambridge, Cambridge, UK. [66]Novo Nordisk Foundation Center for Basic Metabolic Research, Faculty of Health and Medical Sciences, University of Copenhagen, Copenhagen, Denmark. [67]Department of Human Genetics, University of Michigan, Ann Arbor, MI, USA. [68]Computational Medicine, Berlin Institute of Health at Charité–Universitätsmedizin Berlin, Berlin, Germany. [69]Precision Healthcare University Research Institute, Queen Mary University of London, London, UK.

### Authors and contributors associated with the unpublished manuscript: "Genome-wide association study of postprandial glucose metabolism identifies candidate insulin-stimulated glucose uptake genes".

Alice Williamson[1,2], Dougall M Norris[2], Xianyong Yin[3,4], K. Alaine Broadway[5], Anne H Moxley[5], Swarooparani Vadlamudi[5], Emma P Wilson[5], Anne U Jackson[4], Vasudha Ahuja[6], Mette K Andersen[7], Zorayr Arzumanyan[8], Lori L Bonnycastle[9], Stefan R Bornstein[10,11,12], Maxi P. Bretschneider[10,11,12], Thomas A Buchanan[13], Yi-Cheng Chang[14], Lee-Ming Chuang[15], Ren-Hua Chung[16], Tine D Clausen[17,18], Peter Damm[19,20,21], Graciela E Delgado[22], Vanessa D de Mello[23], Josée Dupuis[24,25], Om P Dwivedi[6], Michael R Erdos[9], Lilian Fernandes Silva[26], Tim M Frayling[27], Christian Gieger[28,29], Mark O Goodarzi[30], Xiuqing Guo[8], Stefan Gustafsson[31], Liisa Hakaste[6], Ulf Hammar[32], Gad Hatem[33], Sandra Herrmann[34,35], Kurt Højlund[36], Katrin Horn[37,38], Willa A Hsueh[39], Yi-Jen Hung[40], Chii-Min Hwu[41], Anna Jonsson[7], Line L Kårhus[42], Marcus E Kleber[43,44], Peter Kovacs[45], Timo A Lakka[46,47,48], Marie Lauzon[49], I-Te Lee[50], Cecilia Lindgren[51,52,53,54], Jaana Lindström[55], Allan Linneberg[42,56], Ching-Ti Liu[57], Jian'an Luan[1], Dina Mansour Aly[58], Elisabeth Mathiesen[19,20,59], Angela P Moissl[60,61,43], Andrew P Morris[62], Narisu

Narisu[9], Nikolaos Perakakis[10,63,12], Annette Peters[28,29], Rashmi B Prasad[33,64], Roman N Rodionov[65,66], Kathryn Roll[8,67], Carsten F Rundsten[7], Chloé Sarnowski[68], Kai Savonen[48], Markus Scholz[37,38], Sapna Sharma[69,70], Sara E Stinson[7], Sufyan Suleman[7], Jingyi Tan[8], Kent D Taylor[8], Matti Uusitupa[71], Dorte Vistisen[72,73], Daniel R Witte[74,75], Romy Walther[76], Anny H Xiang[77], Björn Zethelius[78], The Meta-Analysis of Glucose and Insulin-related Traits Consortium (MAGIC)[79], Emma Ahlqvist[58], Richard N Bergman[80], Yii-Der Ida Chen[49], Francis S Collins[9], Tove Fall[32], Jose C Florez[81,82,83], Andreas Fritsche[84], Harald Grallert[85,28,29], Leif Groop[86,87], Torben Hansen[7], Heikki A Koistinen[88,89,90], Pirjo Komulainen[48], Markku Laakso[91], Lars Lind[92], Markus Loeffler[37,38], Winfried März[93,22], James B Meigs[94,95,96], Leslie J Raffel[97], Rainer Rauramaa[48], Jerome I Rotter[98], Peter E. H. Schwarz[34,99,12], Michael Stumvoll[45], Johan Sundström[31], Anke Tönjes[45], Tiinamaija Tuomi[100,101], Jaakko Tuomilehto[102,103], Robert Wagner[104], Inês Barroso[105], Mark Walker[106], Niels Grarup[107], Michael Boehnke[3,4], Nicholas J Wareham[1], Karen L Mohlke[*,#,5], Eleanor Wheeler[*,#,1], Stephen O'Rahilly[*,#,2], Daniel J Fazakerley[*,#,2], Claudia Langenberg[*,#,1,108,109]

[1]MRC Epidemiology Unit, Institute of Metabolic Science, University of Cambridge School of Clinical Medicine, Cambridge, CB2 0QQ, UK, [2]Metabolic Research Laboratories, Wellcome Trust-MRC Institute of Metabolic Science, Department of Clincial Biochemistry, University of Cambridge, Cambridge, CB2 0SL, UK, [3]Department of Biostatistics, University of Michigan, Ann Arbor, MI, USA, [4]Center for Statistical Genetics, University of Michigan, Ann Arbor, MI, USA, [5]Department of Genetics, University of North Carolina, Chapel Hill, NC, 27599, USA, [6]Institute for Molecular Medicine Finland, University of Helsinki, Helsinki, Finland, [7]Novo Nordisk Foundation Center for Basic Metabolic Research, Faculty of Health and Medical Sciences, University of Copenhagen, Copenhagen, 2200, Denmark, [8]Pediatrics, Genomic Outcomes, The Institute for Translational Genomics and Population Sciences, The Lundquist Institute for Biomedical Innovation at Harbor-UCLA Medical Center, Torrance, CA, 90502, USA, [9]Center for Precision Health research, National Human Genome Research Institute, National Institutes of Health, Bethesda, MD, 20892, USA, [10]Department of Internal Medicine III, Metabolic and Vascular Medicine, Medical Faculty Carl Gustav Carus, Dresden, o1307, Germany, [11]Helmholtz Zentrum München, Paul Langerhans Institute Dresden (PLID), University Hospital and Faculty of Medicine, TU Dresden, Dresden, o1308, Germany, [12]German Center for Diabetes Research (DZD e.V.), Neuherberg, Germany, [13]Medicine, Endocrine, Keck School of Medicine USC, Los Angeles, CA, 90033, USA, [14]Graduate Institute of Medical Genomics and Proteomics, National Taiwan University, 5F, No.2, Xuzhou Rd., Zhongzheng Dist., Taipei City, 100025, Taiwan, [15]Internal Medicine, National Taiwan University Hospital; No. 7, Chung-Shan South Road, Taipei City, 100225, Taiwan, [16]Institute of Population Health Sciences, National Health Research Institutes; 35 Keyan Road, Zhunan, Miaoli County, 35053, Taiwan, [17]Department of Gynecology and Obstetrics, Nordsjaellands Hospital, Hilleroed, 3400, Denmark, [18]Department of Clinical Medicine, University of Copenhagen, Copenhagen, Denmark, [19]Department of Clinical Medicine, Faculty of Health and Medical Sciences, University of Copenhagen, Copenhagen, 2200, Denmark, [20]Center for Pregnant Women with Diabetes, Rigshospitalet, Copenhagen, 2100, Denmark, [21]Department of Obstetrics, Rigshospitalet, Copenhagen, 2100, Denmark, [22]Vth Department of Medicine, Medical Faculty Mannheim, Heidelberg Univeristy, Mannheim, BW, 68167, Germany, [23]Institute of Public Health and Clinical Nutrition, University of Eastern Finland, Kuopio, Finland, [24]Department of Biostatistics, Boston University School of Public Health, Boston, MA, 2118, USA, [25]Department of Epidemiology, Biostatistics and Occupational Health, McGill University, Montréal, Canada, [26]Institute of Clinical Medicine, University of Eastern Finland, Kuopio, 70210, Finland, [27]College of Medicine and Health, Exeter University, Exeter, UK, [28]Institute of Epidemiology II, Helmholtz Zentrum München-German Research Center for Environmental Health, Neuherberg, Germany, [29]German Center for Diabetes Research, Neuherberg, Germany, [30]Medicine, Endocrinology, Diabetes & Metabolism, Cedars-Sinai Medical Center, Los Angeles, CA, 90048, USA, [31]Medical Sciences, Clinical Epidemiology, Uppsala University, Uppsala, Uppsala, 75185, Sweden, [32]Medical Sciences, Molecular Epidemiology, Uppsala University, Uppsala, Uppsala, 75185, Sweden, [33]Genomics, Diabetes and Endocrinology, Department of Clinical Sciences, Lund University, Malmö, Sweden, [34]Department of Internal Medicine III, Prevention and Care of Diabetes, Medical Faculty Carl Gustav Carus, Dresden, o1307, Germany, [35]Helmholtz Zentrum München, Paul Langerhans Institute Dresden (PLID), University Hospital and Faculty of Medicine, TU Dresden, Dresden, o1310, Germany, [36]Steno Diabetes Center Odense, Odense, Denmark, [37]Medical Faculty, Institute for Medical Informatics, Statistics and Epidemiology, Leipzig, 4107, Germany, [38]LIFE Research Center

for Civilization Diseases, Medical Faculty, Leipzig, 4103, Germany, [39]Internal Medicine, Endocrinology, Diabetes & Metabolism, The Ohio State University Wexner Medical Center, Columbus, OH, 43210, USA, [40]Institute of Preventive Medicine, National Defense Medical Center, Taipei, Taiwan; Postbox 90048~700, Sanhsia Dist, New Taipei City, 237010, Taiwan, [41]Medicine, Section of Endocrinology and Metabolism, Taipei Veterans General Hospital, Taipei, Taiwan, No. 201, Section 2, Shipai Road, Beitou District, Taipei City, 112201, Taiwan, [42]Center for Clinical Research and Prevention, Copenhagen University Hospital – Bispebjerg and Frederiksberg, Copenhagen, 2000, Denmark, [43]Vth Department of Medicine, Medical Faculty Mannheim, Heidelberg Univeristy, Mannheim, 68167, Germany, [44]SYNLAB MVZ Humangenetik Mannheim, Mannheim, 68163, Germany, [45]Medical Department III-Endocrinology, Nephrology, Rheumatology, University of Leipzig Medical Center, Leipzig, Germany, [46]Institute of Biomedicine, [47]Department of Clinical Physiology and Nuclear Medicine, Kuopio University Hospital, Kuopio, Finland, [48]Foundation for Research in Health Exercise and Nutrition, Kuopio Research Institute of Exercise Medicine, Kuopio, Finland, [49]Pediatrics, Genomic Outcomes, The Institute for Translational Genomics and Population Sciences, The Lundquist Institute at Harbor-UCLA Medical Center, Torrance, CA, 90502, USA, [50]Internal Medicine, Endocrinology and Metabolism, Taichung Veterans General Hospital; No. 1650, Sec. 4, Taiwan Boulevard, Xitun District, Taichung City, 40705, Taiwan, [51]Big Data Institute, Li Ka Shing Centre for Health Information and Discovery,, University of Oxford, Oxford, UK, [52]NDPH, NDPH, University of Oxford, Oxford, UK, [53]Wellcome Trust Centre Human Genetics, University of Oxford, Oxford, UK, [54]Broad Institute of Harvard and MIT, Havard and MIT, Boston, MA, USA, [55]Finnish Institute for Health and Welfare, Helsinki, Finland, [56]Department of Clinical Medicine, Faculty of Health and Medical Sciences, University of Copenhagen, Copenhagen, 2200, Denmark, [57]Biostatistics, Boston University School of Public Health, Boston, MA, 2118, USA, [58]Clinical Sciences, Genomics, Diabetes and Endocrinology, Lund University, Malmö, 20502, Sweden, [59]Department of Endocrinology, Rigshospitalet, Copenhagen, 2100, Denmark, [60]Institute of Nutritional Sciences, Friedrich-Schiller-University, Jena, 7743, Germany, [61]Competence Cluster for Nutrition and Cardiovascular Health (nutriCARD) Halle-Jena, Jena, 7743, Germany, [62]Centre for Genetics and Genomics Versus Arthritis, Centre for Musculoskeletal Research, The University of Manchester, Manchester, UK, [63]Paul Langerhans Institute Dresden (PLID), University Hospital and Faculty of Medicine, TU Dresden, Dresden, o1309, Germany, [64]Institute for Molecular Medicine Finland (FIMM), University of Helsinki, Helsinki, Finland, [65]Department of Internal Medicine III, University Center for Vascular Medicine, Medical Faculty Carl Gustav Carus, Dresden, o1308, Germany, [66]College of Medicine and Public Health, Flinders University and Flinders Medical Centre,, Adelaide, Australia, [67]Helmholtz Zentrum München, Paul Langerhans Institute Dresden (PLID), University Hospital and Faculty of Medicine, TU Dresden, Dresden, o1311, Germany, [68]Epidemiology, Human Genetics & Environmental Sciences, The University of Texas Health Science Center, Houston, TX, 77030, USA, [69]Research Unit of Molecular Epidemiology, Helmholtz Zentrum Muenchen, Helmholtz Zentrum München-German Research Center for Environmental Health, Neuherberg, Germany, [70]Chair of Food Chemistry and Molecular and Sensory Science, Technical University of Munich, Freising, Germany, [71]Department of Public Health and Clinical Nutrition, University of Eastern Finland, Kuopio, Finland, [72]Clinical Research, Steno Diabetes Center Copenhagen, Herlev, 2730, Denmark, [73]Department of Public Health, University of Copenhagen, Copenhagen, 1353, Denmark, [74]Steno Diabetes Center Aarhus, Aarhus, 8200, Denmark, [75]Department of Public Health, Aarhus University, Aarhus, 8200, Denmark, [76]Department of Internal Medicine III, Pathobiochemistry, Medical Faculty Carl Gustav Carus, Dresden, o1308, Germany, [77]Pediatrics, Genetic and Genomic medicine, University of California Irvine, Irvine, CA, 92697, USA, [78]Public Health and Caring Sciences, Geriatrics, Uppsala University, Uppsala, Uppsala, 75237, Sweden, [79](No affiliation data provided), [80]Diabetes and Obesity Research Institute, Cedars-Sinai Medical Center, Los Angeles, CA, USA, [81]Diabetes Unit and Center for Genomic Medicine, Massachusetts General Hospital, Boston, MA, 2114, USA, [82]Programs in Metabolism and Medical & Population Genetics, The Broad Institute, Boston, MA, 2142, USA, [83]Harvard Medical School, Boston, MA, 2115, USA, [84]Internal Medicine, Diabetology, Tübingen, 72076, Germany, [85]Research Unit of Molecular Epidemiology, Helmholtz Zentrum München-German Research Center for Environmental Health, Neuherberg, Germany, [86]Diabetes Centre, Lund University, Lund, Sweden, [87]Finnish Institute of Molecular Medicine, Helsinki University, Helsinki, Finland, [88]Department of Public Health and Welfare, Finnish Institute for Health and Welfare, Helsinki, FI-00271, Finland, [89]Department of Medicine, University of Helsinki and Helsinki University Hospital, Helsinki, FI-00029, Finland, [90]Minerva Foundation Institute for Medical Research, Helsinki, FI-00290, Finland, [91]Institute of Clinical Medicine, University of Eastern Finland, 70210, Finland, [92]Medical Sciences, Clinical Epidemiology, Uppsala University, Uppsala, 75185, Sweden, [93]Synlab Academy, SYNLAB Holding Deutschland GmbH, Mannheim, BW, 68167, Germany, [94]Medicine, Division of General Internal Medicine, Massachusetts General Hospital, Boston, MA, 2114, USA, [95]Medicine, Harvard Medical School, Boston, MA, 2115, USA, [96]Broad Institute, Cambridge, MA, 2146, USA, [97]Department of Pediatrics, Genetic and Genomic Medicine, University of California, Irvine, CA, USA, [98]The

Institute for Translational Genomics and Population Sciences, Department of Pediatrics, The Lundquist Institute for Biomedical Innovation at Harbor-UCLA Medical Center, Torrance, CA, USA, [99]Helmholtz Zentrum München, Paul Langerhans Institute Dresden (PLID), University Hospital and Faculty of Medicine, TU Dresden, Dresden, o1307, Germany, [100]Folkhälsan Research Center, Helsinki, Finland, [101]Finnish Institute for Molecular Medicine, University of Helsinki, Helsinki, Finland, [102]Public Health, University of Helsinki, Helsinki, Finland, [103]National Institute for Health and Welfare, Helsinki, Finland, [104]Intenal Medicine, Diabetology, Tübingen, 72076, Germany, [105]Exeter Centre of Excellence for Diabetes Research (EXCEED), Genetics of Complex Traits, University of Exeter Medical School, University of Exeter, Exeter, UK, [106]Faculty of Medical Sciences, Newcastle University, Newcastle upon Tyne, UK, [107]Novo Nordisk Foundation Center for Basic Metabolic Research, Faculty of Health and Medical Sciences, University of Copenhagen, Copenhagen, 2200, Denmark, [108]Computational Medicine, Berlin Institute of Health at Charité–Universitätsmedizin, Berlin, Germany, [109]Precision Healthcare University Research Institute, Queen Mary University of London, London, UK.

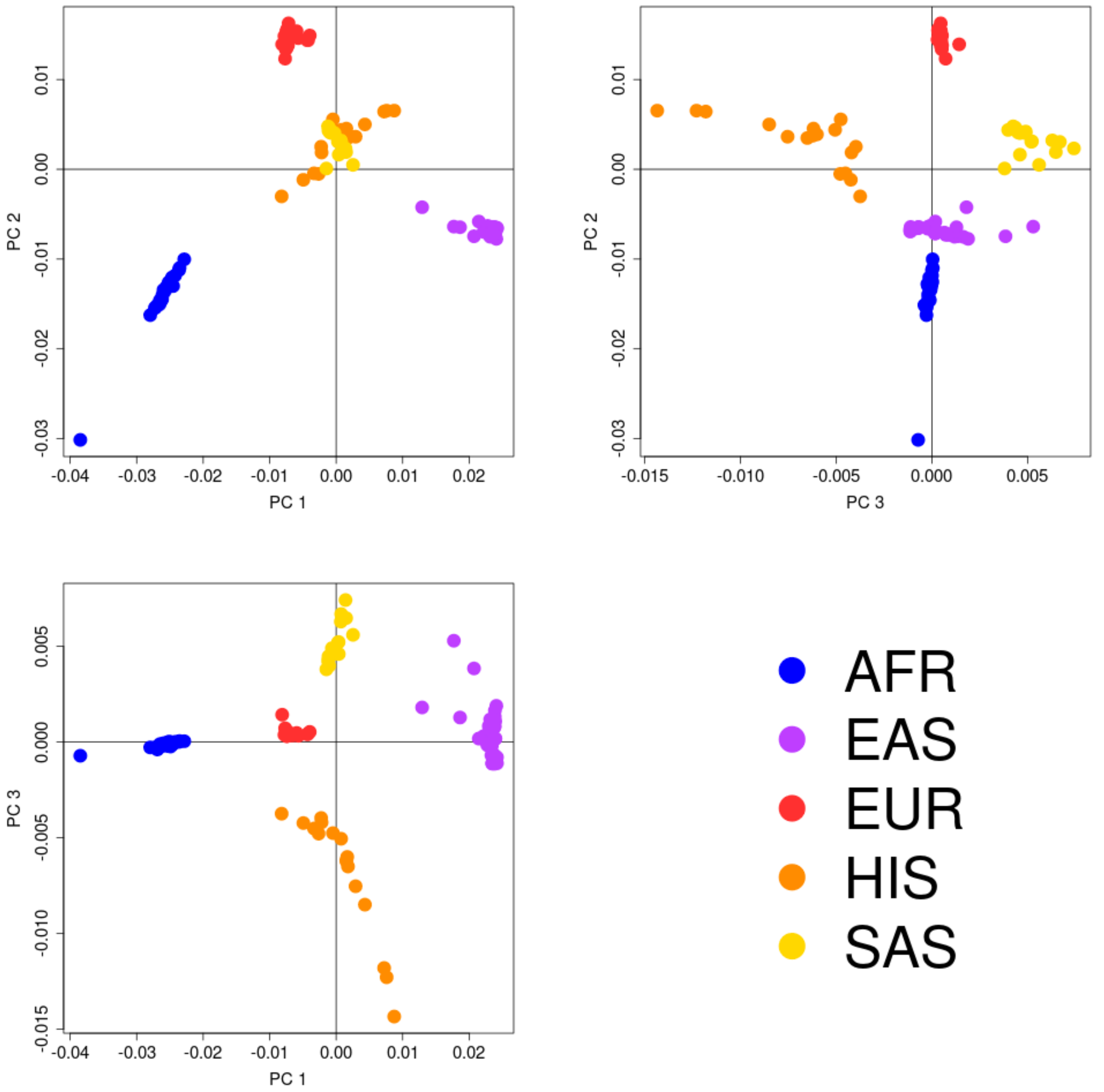


**Supplementary Figure 1. Axes of genetic variation separating GWAS of T2D across ancestry groups.** The first three axes of genetic variation (PC 1, PC 2 and PC 3) from multi-dimensional scaling of the Euclidean distance matrix between populations are sufficient to separate five ancestry groups: African (AFR), East Asian (EAS), European (EUR), Hispanic (HIS), and South Asian (SAS).

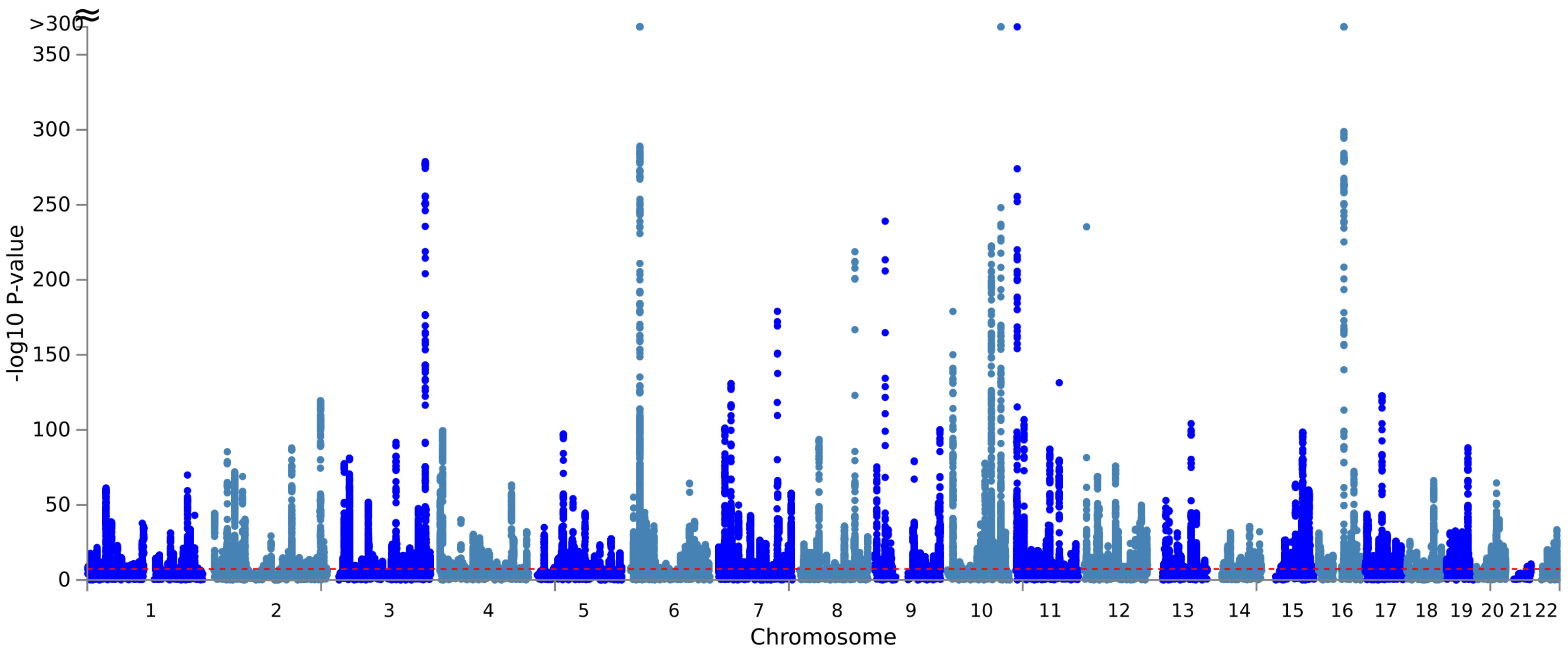


**Supplementary Figure 2. Manhattan plot of genome-wide T2D association from multi-ancestry meta-regression (MR-MEGA) of up to 428,452 T2D cases and 2,107,149 controls across multiple ancestry groups.** Each point represents a SNV passing quality control in the multi-ancestry meta-regression, plotted with their association p-value (on a $-\log_{10}$ scale, truncated at 300) as a function of genomic position (NCBI build 37). Genome-wide significance ($P<5\times10^{-8}$) is highlighted by the dashed horizontal red line.

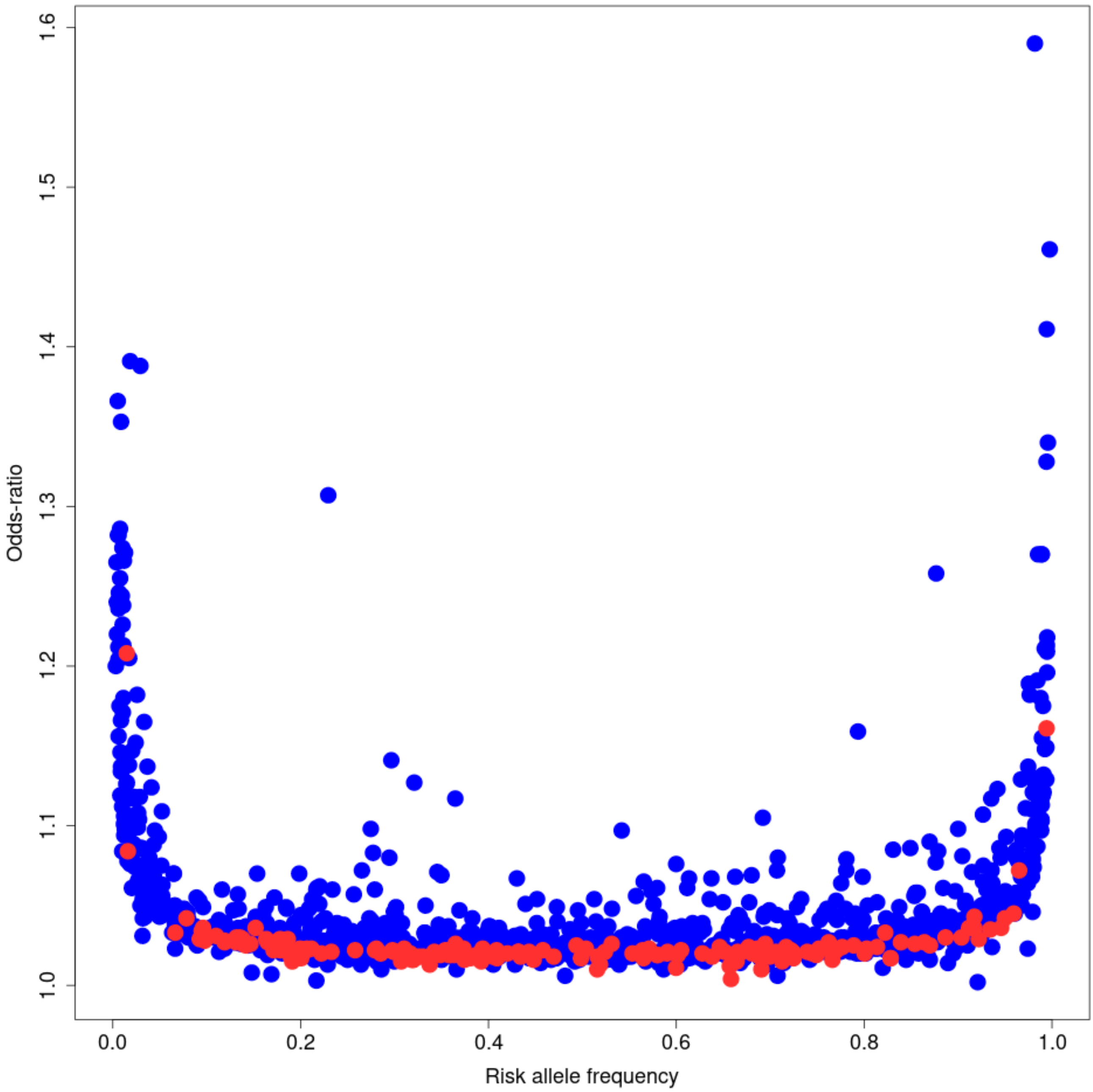


**Supplementary Figure 3. Distribution of risk allele frequency and odds-ratio at index SNVs for distinct T2D association signals.** Each point corresponds to an index SNV, plotted according to the mean risk allele frequency across GWAS (on the x-axis) and the odds-ratio from fixed-effects meta-analysis (on the y-axis). Index SNVs highlighted in blue map to previously reported loci for T2D susceptibility. Index SNVs highlighted in red do not map to previously reported loci for T2D susceptibility.

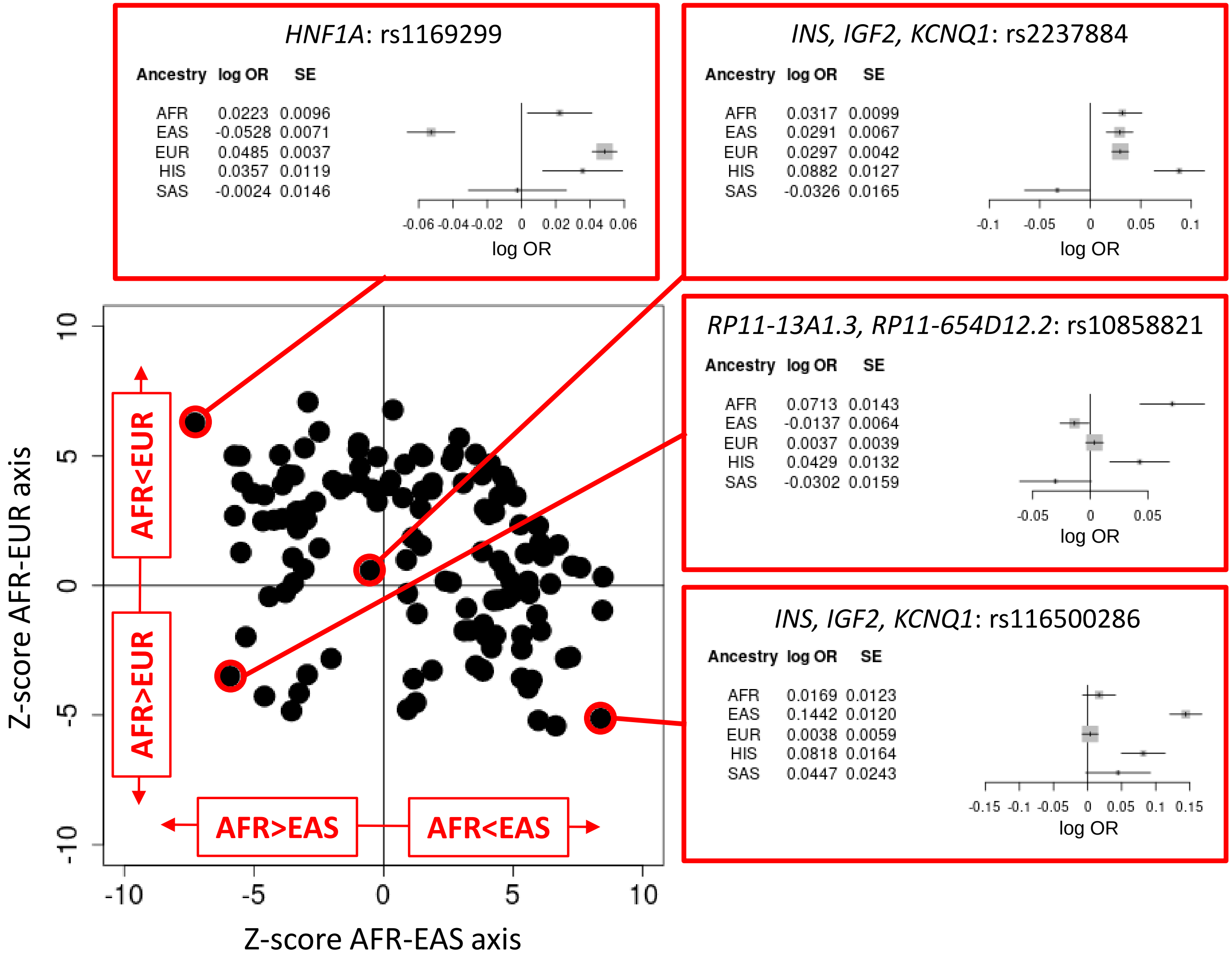


**Supplementary Figure 4. Ancestry-correlated heterogeneity is driven by differences in allelic effect sizes between African, East Asian, and European ancestry groups.** In the bottom left panel, index SNVs with significant evidence ($P_{HET}$<3.9x10$^{-5}$, Bonferroni correction for 1,289 signals) for ancestry-correlated heterogeneity are plotted according to their association (Z-score) with the first two axes of genetic variation. The first axis represents differences in allelic effect sizes between GWAS of African and East Asian ancestry (AFR-EAS axis), whilst the second axis represents differences in effect size between GWAS of African and European ancestry (AFR-EUR axis). The forest plots present examples of ancestry-correlated heterogeneity at index SNVs. In each forest plot, the allelic log-odds ratio (OR) from each ancestry group-specific fixed-effects meta-analysis is given by the black tick mark, the 95% confidence interval is given by the horizontal line, and the weight (inverse-variance) of each ancestry group by the grey box. AFR: African ancestry group. EAS: East Asian ancestry group. EUR: European ancestry group. HIS: Hispanic ancestry group. SAS: South Asian ancestry group.

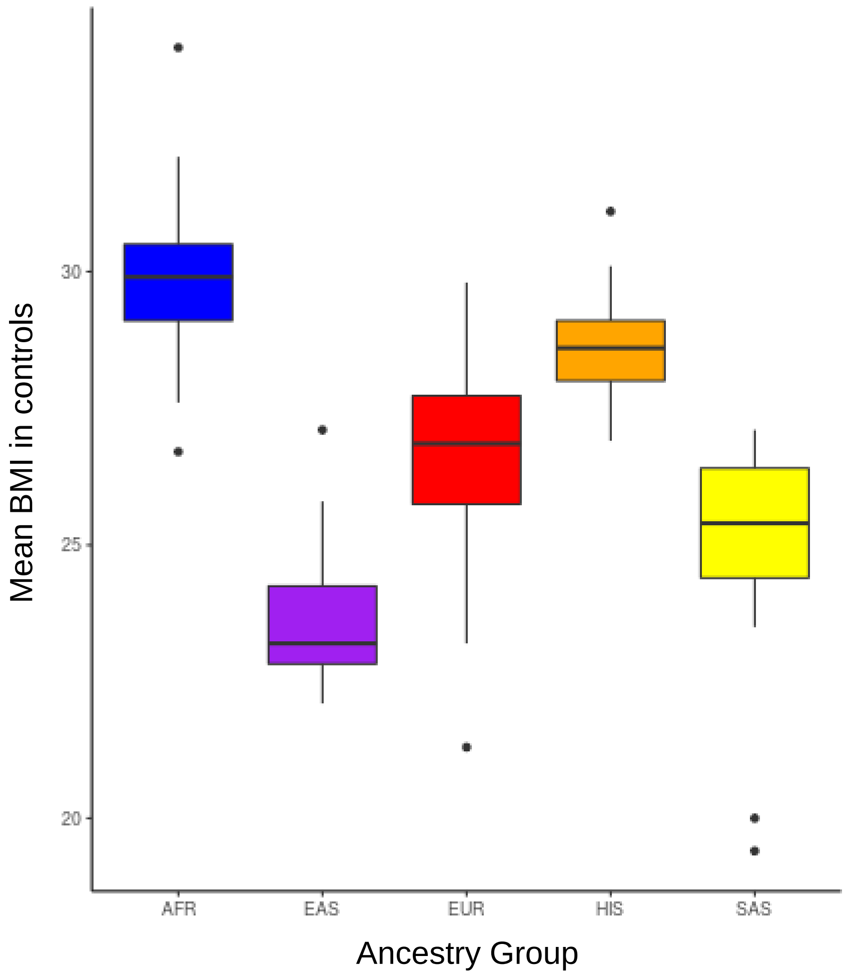


**Supplementary Figure 5. Distribution of study-level mean BMI in control individuals across ancestry groups.** Each box and whisker plot presents the median (back horizontal line), upper and lower quartiles (extremes of coloured boxes), minimum and maximum (excluding outliers, extremes of black vertical line), and outliers (more than 1.5x inter-quartile range, black dots). AFR: African ancestry group. EAS: East Asian ancestry group. EUR: European ancestry group. HIS: Hispanic ancestry group. SAS: South Asian ancestry group.

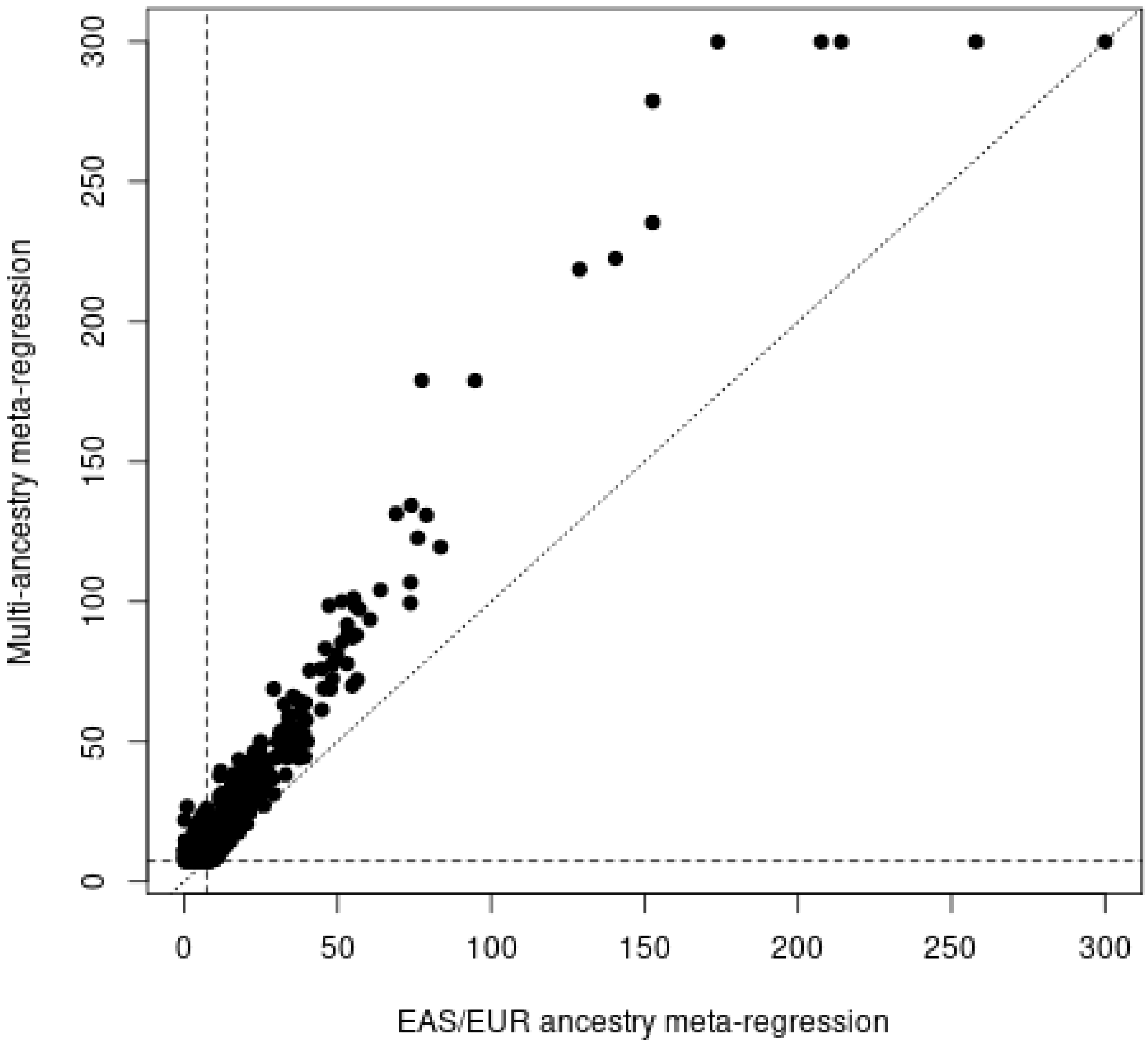


**Supplementary Figure 6. Comparison of association evidence at 1,289 T2D signals between multi-ancestry meta-regression and meta-regression of GWAS from East Asian and European ancestry groups.** Each point corresponds to an index SNV, plotted according to the $-\log_{10}$ P-value (cut-off at 300 for ease of presentation) for the multi-ancestry meta-regression on the x-axis and the East Asian and European ancestry meta-regression on the y-axis. The dashed horizontal and vertical lines define genome-wide significance ($P$<5x10$^{-8}$). T2D signals represented by index SNVs above the dotted y=x line show stronger evidence of association in the multi-ancestry meta-regression than in the meta-regression of GWAS from East Asian and European ancestry groups. EAS: East Asian ancestry group. EUR: European ancestry group.

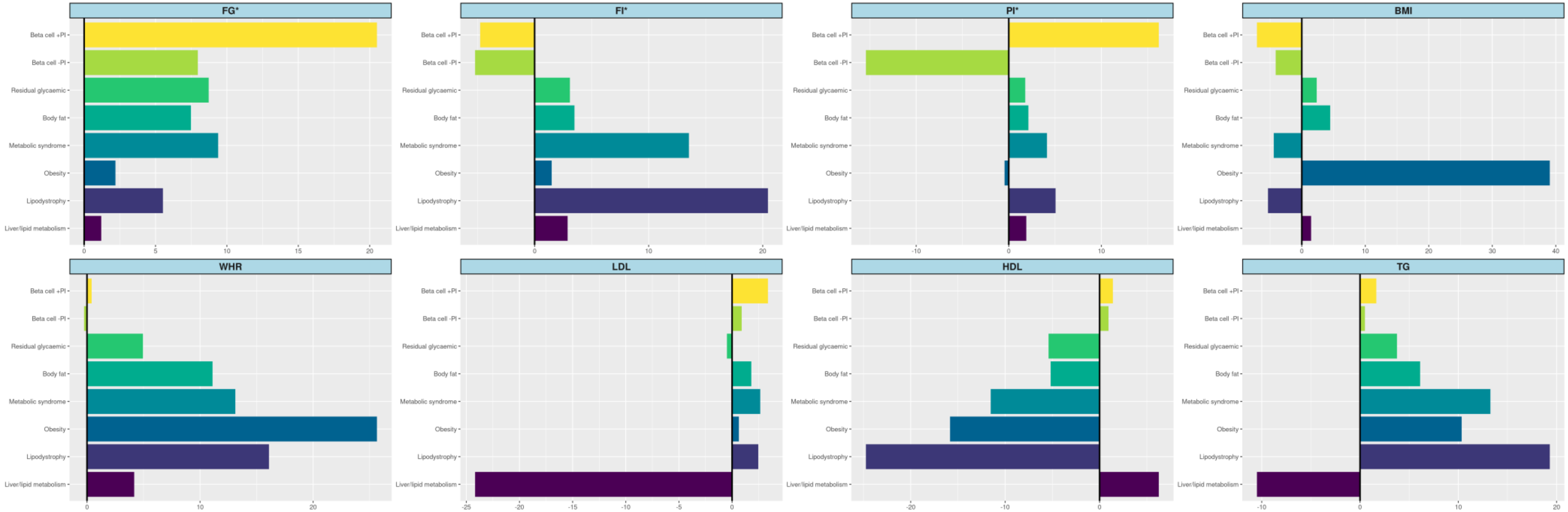


**Supplementary Figure 7. Cluster-specific associations of index SNVs with defining cardiometabolic phenotypes.** Each bar presents -$\log_{10}$ *P*-value for association, with effect direction aligned to the T2D risk allele. FG: fasting glucose. FI: fasting insulin. PI: proinsulin. BMI: body mass index. WHR: waist-hip ratio. LDL: low-density lipoprotein cholesterol. HDL: high-density lipoprotein cholesterol. TG: triglycerides. *Trait adjusted for BMI.

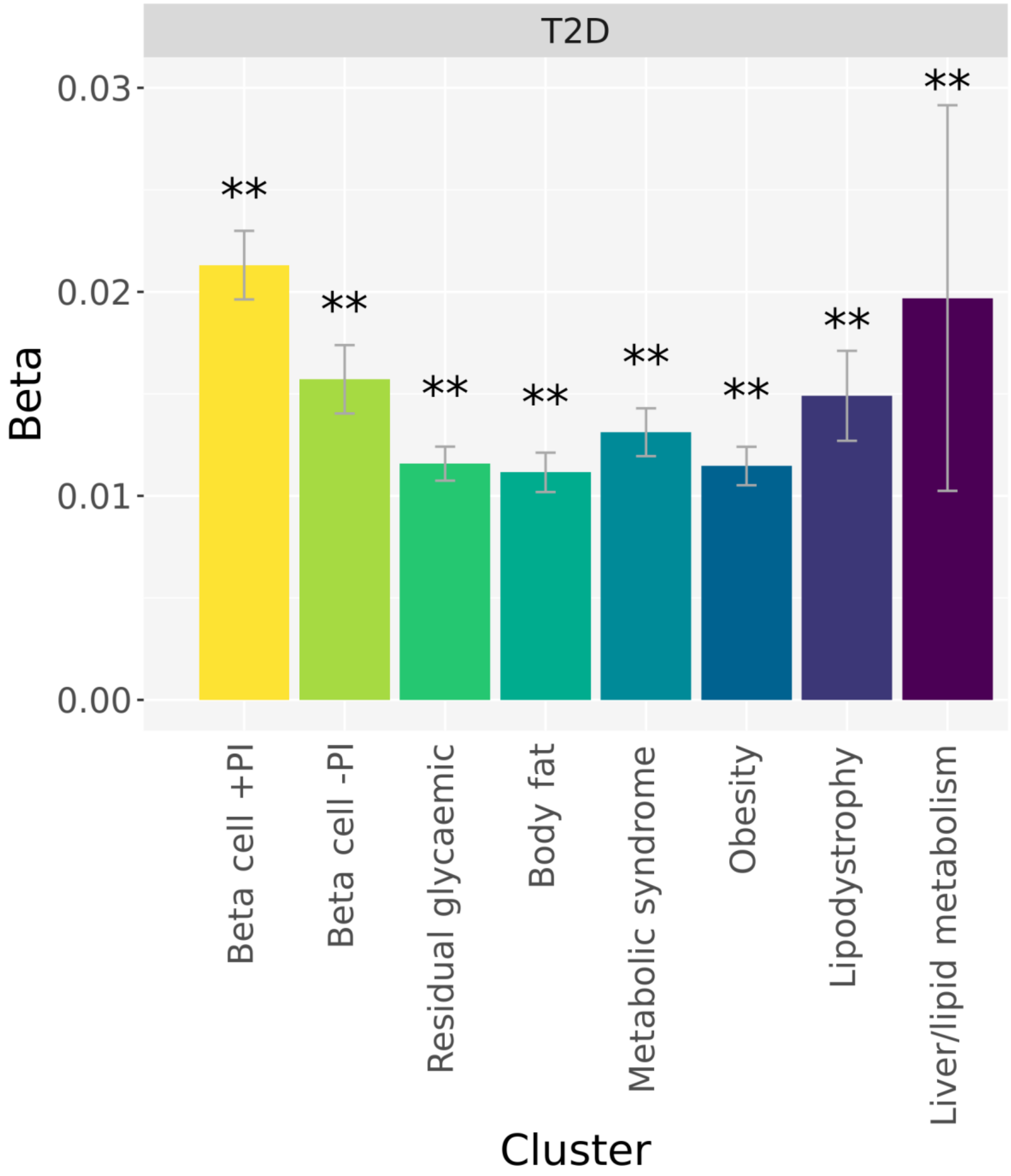


**Supplementary Figure 8. Cluster-specific associations of index SNVs with T2D.** The height of each bar corresponds to the log-odds ratio (beta), and the grey bar shows the 95% confidence interval. *$P$<0.05, nominal association. **$P$<0.0063, Bonferroni correction for eight clusters.

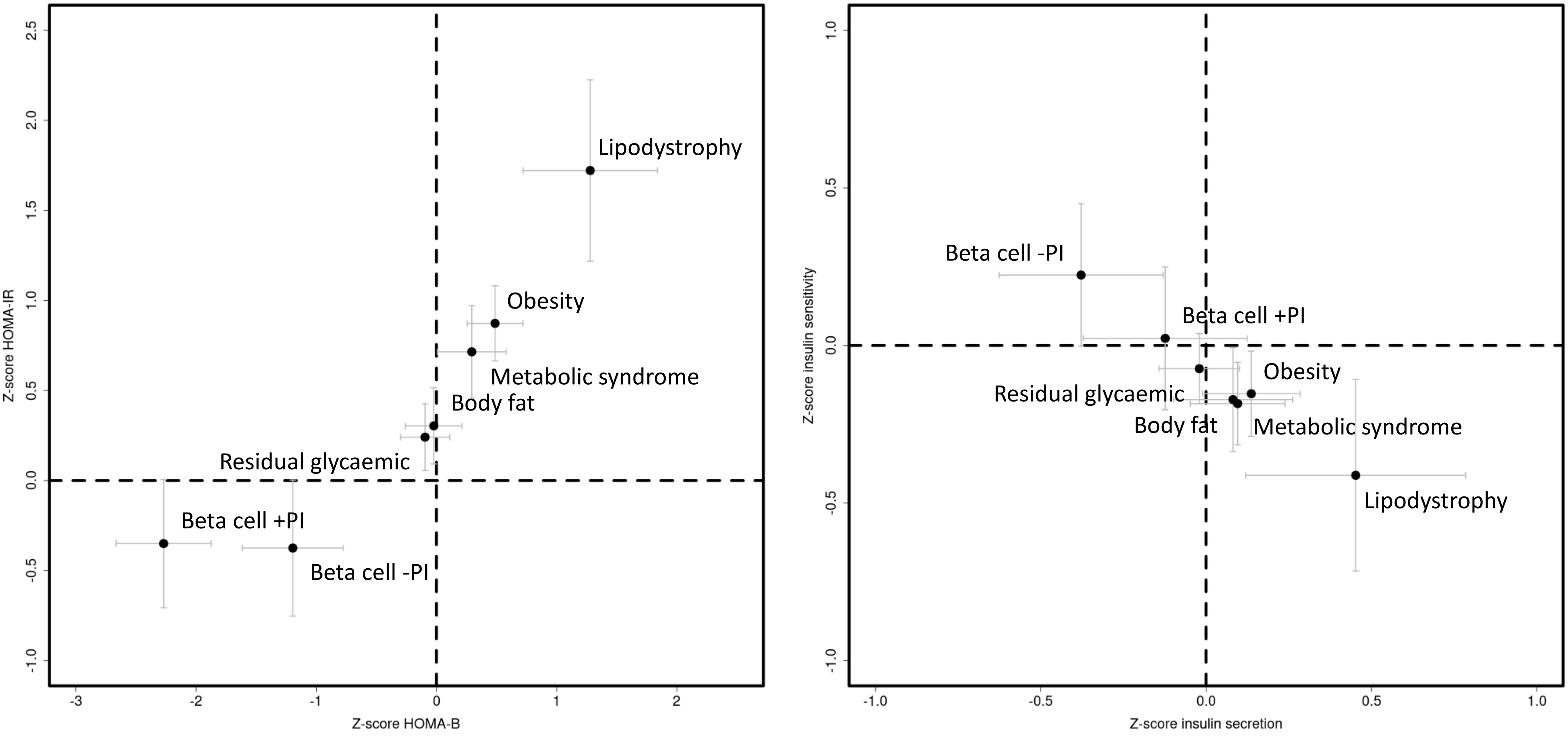


**Supplementary Figure 9. Cluster-specific associations of T2D risk alleles at index SNVs with insulin-related endophenotypes.** Measures of insulin secretion and insulin sensitivity were derived from hyperinsulinemic-euglycemic clamp assessments and oral glucose tolerance tests in up to 1,316 Mexican American participants without diabetes. Homeostatic model assessment measures of beta-cell function (HOMA-B) and insulin resistance (HOMA-IR) were obtained from 36,466 non-diabetic individuals of European ancestry. Each point corresponds to the cluster-specific mean Z-score for each trait, and grey bars represent 95% confidence intervals. The liver/lipid metabolism cluster has been removed for ease of presentation.

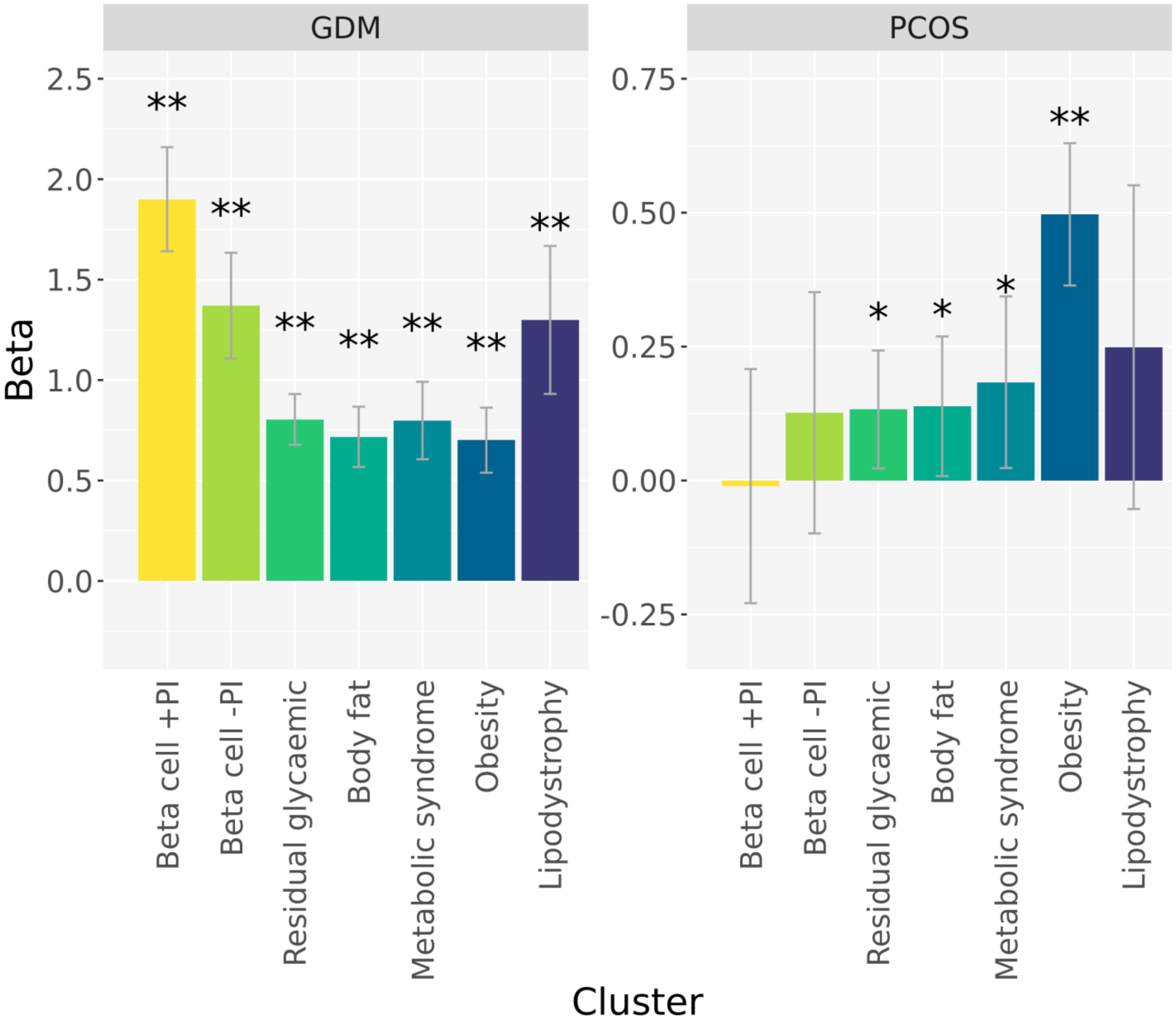


**Supplementary Figure 10. Cluster-specific associations of T2D risk alleles at index SNVs with insulin resistance-related disorders.** Association with gestational diabetes mellitus (GDM) was assessed in 5,485 cases and 347,856 female controls of diverse ancestry. Association with polycystic ovary syndrome (PCOS) was assessed in 10,074 cases and 103,164 female controls of European ancestry. The height of each bar corresponds to the mean Z-score, and the grey bar shows the 95% confidence interval. The liver/lipid metabolism cluster has been removed for ease of presentation. *$P<0.05$, nominal association. **$P<0.0063$, Bonferroni correction for eight clusters.

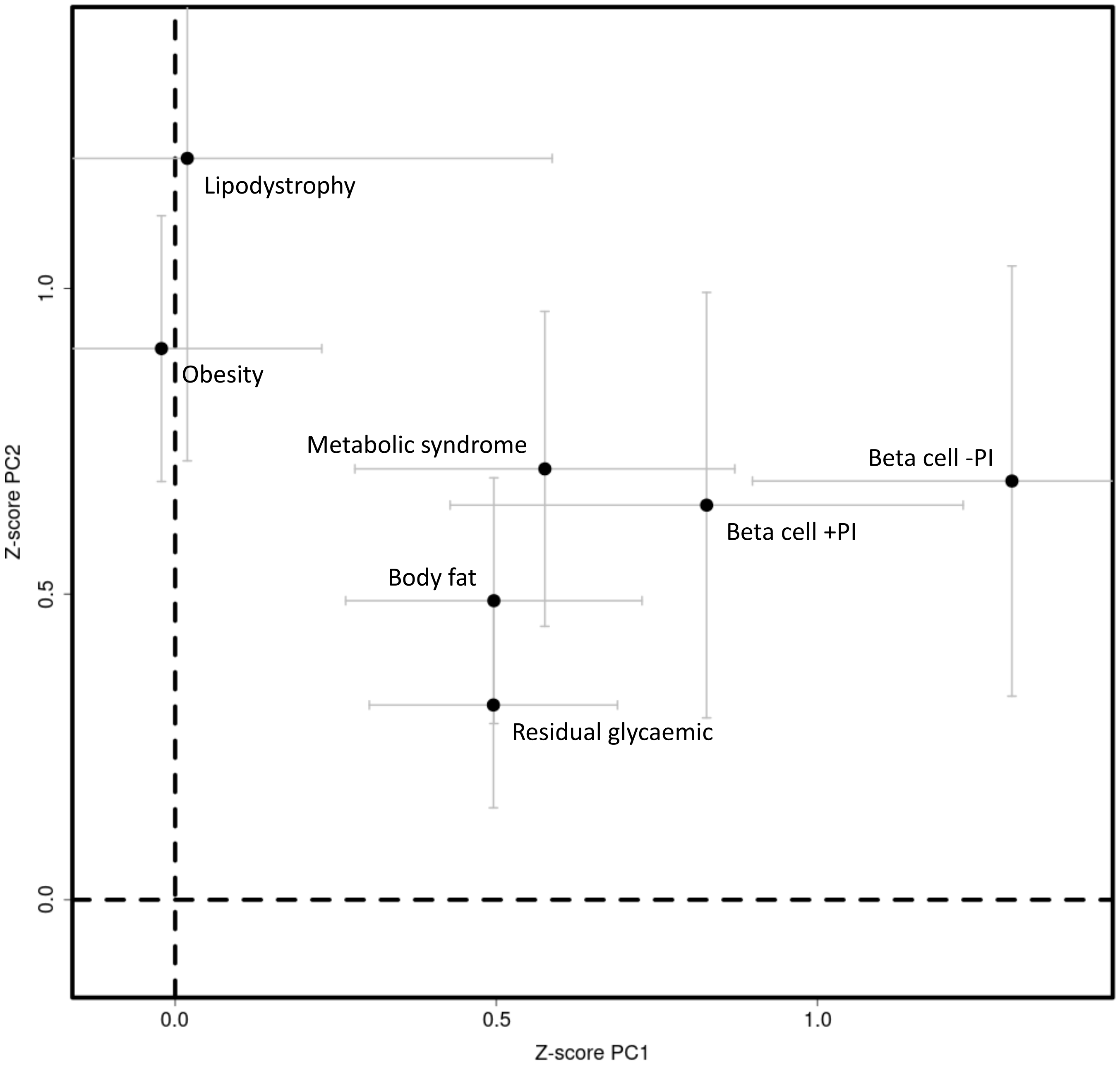


**Supplementary Figure 11. Cluster-specific associations of index SNVs with the first two axes of genetic variation.** Each point corresponds to a cluster, plotted according to the mean Z-score for association with the first two axes of genetic variation (PC1 and PC2) on the x-axis and y-axis, respectively. Grey bars correspond to 95% confidence intervals. The liver/lipid metabolism cluster has been removed for ease of presentation.

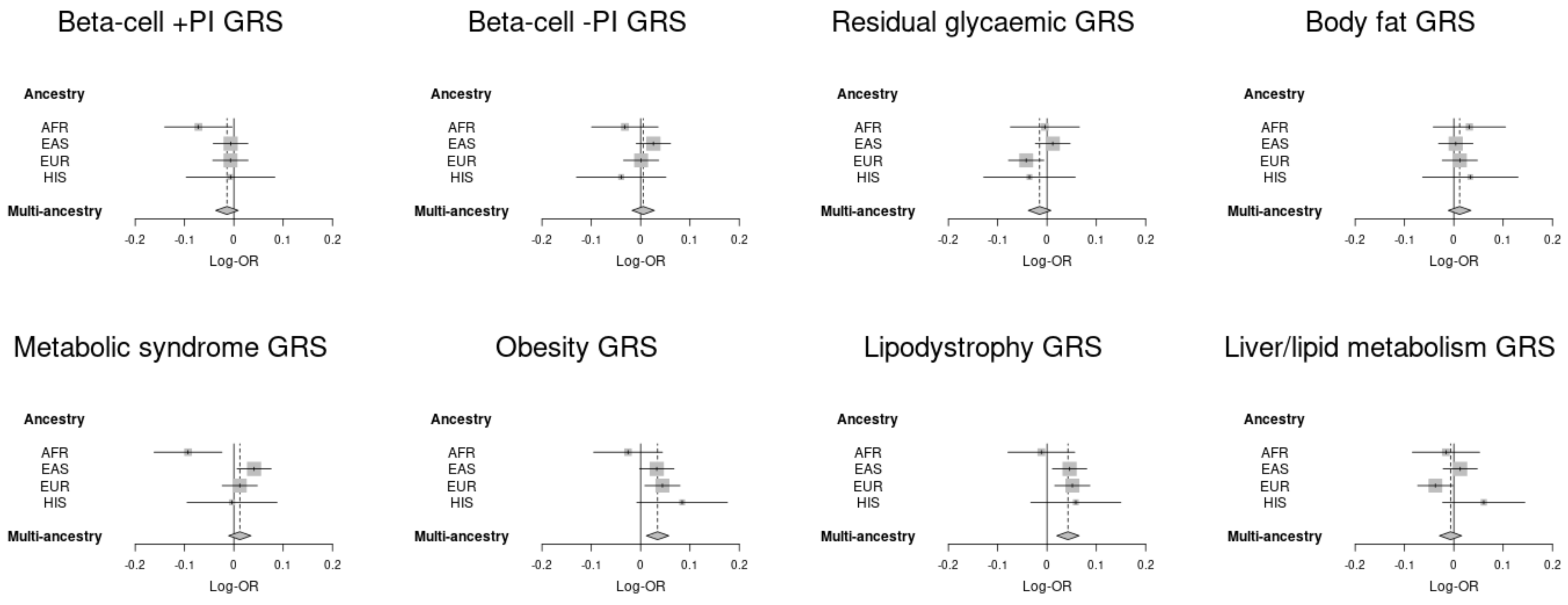


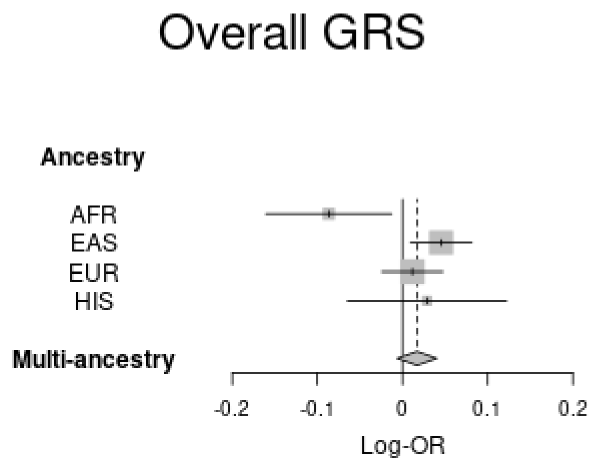


**Supplementary Figure 12. Association of overall T2D GRS and cluster-specific components of partitioned GRS with coronary artery disease across multiple ancestry groups.** In each forest plot, the log-odds ratio (log-OR) of the standardised GRS for each ancestry is presented, together with the 95% confidence interval (horizontal bar) and weight (inverse variance, size of grey box). The grey diamond corresponds to the fixed-effects estimate of the log-OR of the GRS across ancestry groups (upper/lower points of diamond) and corresponding 95% confidence interval (left/right points of diamond). Analyses were conducted in all participants, with adjustment for T2D status. AFR: African ancestry group. EAS: East Asian ancestry group. EUR: European ancestry group. HIS: Hispanic ancestry group.

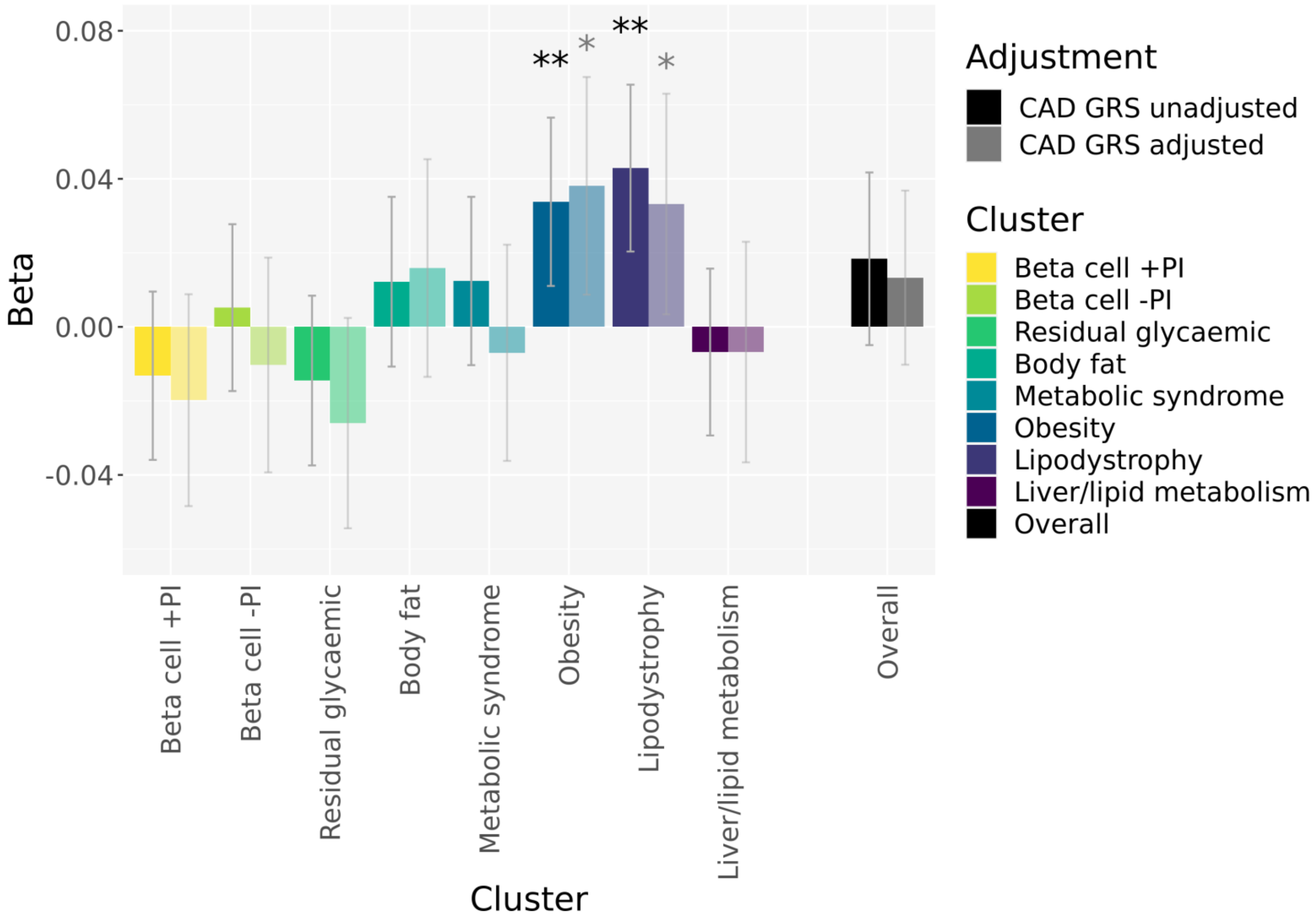


**Supplementary Figure 13. Associations of the overall GRS and cluster-specific partitioned GRS with coronary artery disease (CAD) in up to 137,559 individuals across diverse ancestry groups.** The panel summarizes the associations the overall GRS and each cluster-specific component of the partitioned GRS with CAD, with and without adjustment for a multi-ancestry CAD GRS. The height of each bar corresponds to the log-odds ratio (beta) per standard deviation of the GRS, and the grey bar shows the 95% confidence interval. Analyses were undertaken in all individuals, with adjustment for T2D status. *P<0.05, nominal association. **P<0.0063, Bonferroni correction for eight clusters.

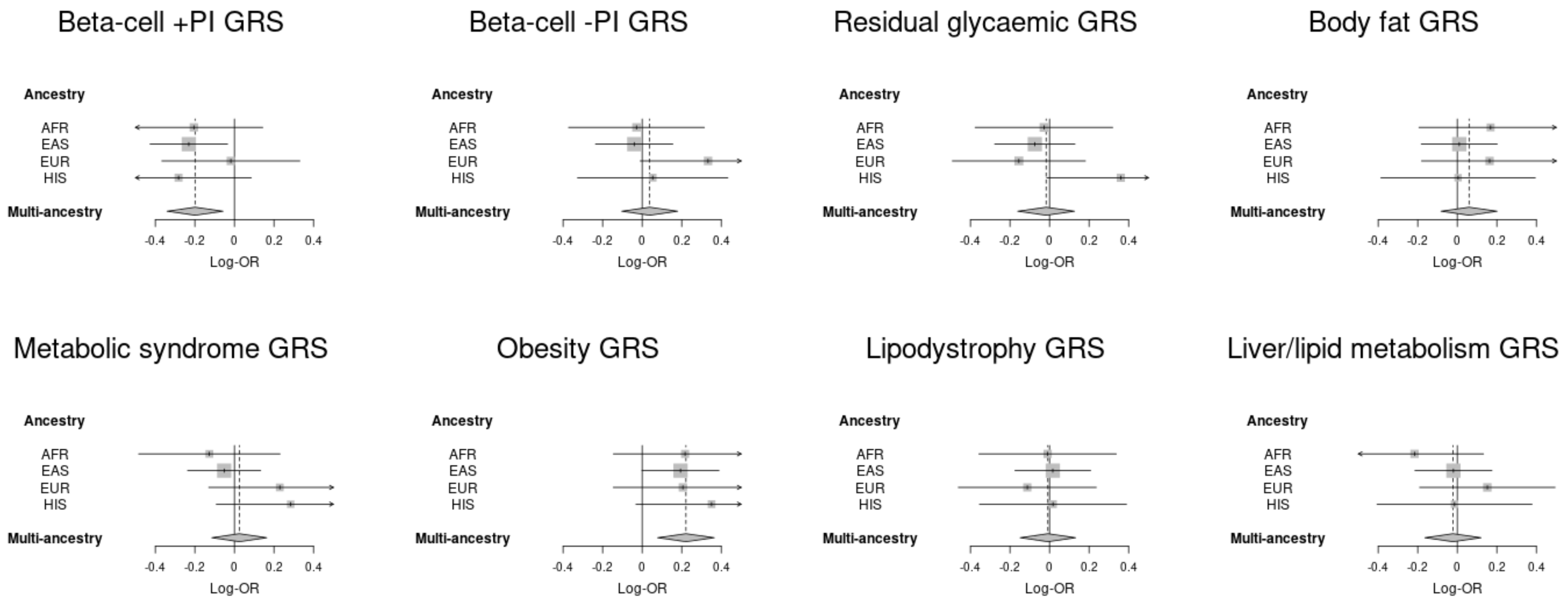


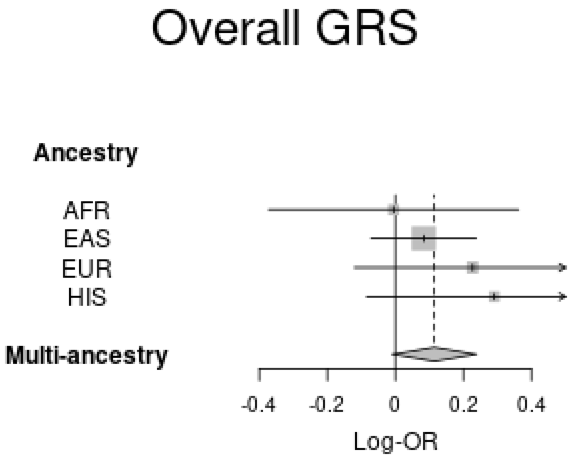


**Supplementary Figure 14. Association of overall T2D GRS and cluster-specific components of partitioned GRS with end-stage diabetic nephropathy across multiple ancestry groups.** In each forest plot, the log-odds ratio (log-OR) of the standardised GRS for each ancestry is presented, together with the 95% confidence interval (horizontal bar) and weight (inverse variance, size of grey box). The grey diamond corresponds to the fixed-effects estimate of the log-OR of the GRS across ancestry groups (upper/lower points of diamond) and corresponding 95% confidence interval (left/right points of diamond). Analyses were conducted in individuals with T2D only. AFR: African ancestry group. EAS: East Asian ancestry group. EUR: European ancestry group. HIS: Hispanic ancestry group.

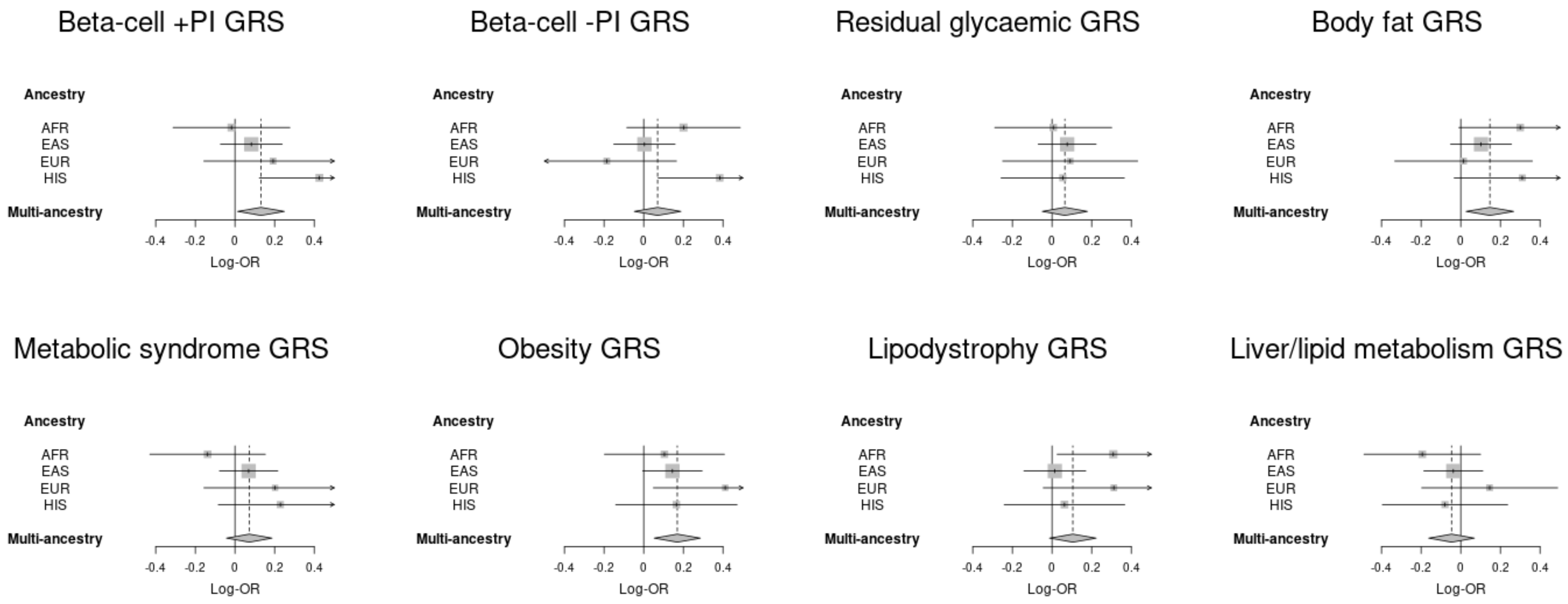


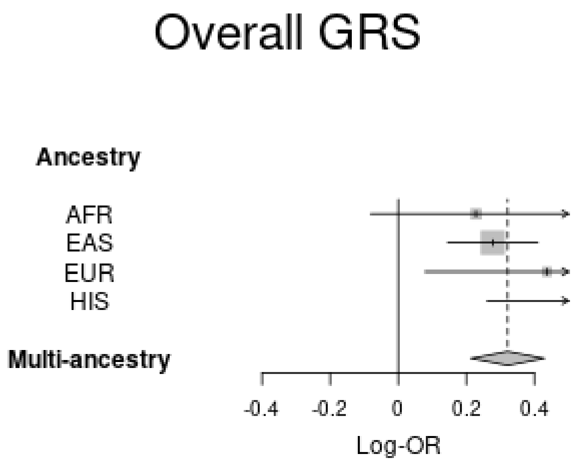


**Supplementary Figure 15. Association of overall T2D GRS and cluster-specific components of partitioned GRS with proliferative diabetic retinopathy across multiple ancestry groups.** In each forest plot, the log-odds ratio (log-OR) of the standardised GRS for each ancestry is presented, together with the 95% confidence interval (horizontal bar) and weight (inverse variance, size of grey box). The grey diamond corresponds to the fixed-effects estimate of the log-OR of the GRS across ancestry groups (upper/lower points of diamond) and corresponding 95% confidence interval (left/right points of diamond). Analyses were conducted in individuals with T2D only. AFR: African ancestry group. EAS: East Asian ancestry group. EUR: European ancestry group. HIS: Hispanic ancestry group.
